# Investigating Indirect Genetic Effects on Neurodevelopmental Traits Across Development and Reporters: Triangulating Twin-comparison and Trio-PGS Approaches

**DOI:** 10.64898/2026.11.09.26367604

**Authors:** Zhaotian Chi, Kaito Kawakami, Robert Plomin, Tom A. McAdams

## Abstract

Polygenic associations with neurodevelopmental traits may reflect both direct and indirect genetic effects. However, evidence separating these effects is limited and mixed, and rarely spans development or reporters, or triangulates across methods. We triangulated twin-comparison and trio-based analyses in the Twins Early Development Study, using eight polygenic scores (PGS) and repeated parent, teacher and self-reports of attention-deficit/hyperactivity disorder (ADHD) and autism spectrum disorder (ASD) traits from ages 4 to 26. Analyses included up to 7,259 offspring with observed or imputed parental genetic data. Both approaches consistently supported direct genetic effects of ADHD and ASD PGS on corresponding traits, whereas evidence for within-trait indirect effects was limited. Cross-trait indirect effects emerged for several PGS, particularly in parent reports, and some were unexpectedly negative. These findings suggest that within-trait polygenic prediction of ADHD and ASD traits mainly reflects direct rather than indirect genetic effects, whereas cross-trait associations reflect some indirect genetic effects.

---

Neurodevelopmental disorders (NDDs) are characterised by atypical brain development which causes impairments in cognitive, emotional, social, or motor functioning^1^. Common NDDs, such as attention-deficit/hyperactivity disorder (ADHD) and autism spectrum disorder (ASD), are highly heritable and show strong intergenerational transmission^2,3^. Genome-wide association studies (GWAS) have identified thousands of common variants contributing to NDDs. When aggregated into polygenic scores (PGS), these variants predict NDD symptoms across development^2,4^.

However, an important consideration of PGS-based prediction is that associations between polygenic scores and outcomes may not solely reflect direct genetic effects (DGE) of inherited alleles^5^. Instead, they can also capture environmentally mediated effects of relatives’ genotypes, known as indirect genetic effects (IGEs), as well as confounding by population stratification and assortative mating. Specifically, Kong and colleagues showed that parental genotypes, both transmitted and non-transmitted, can influence offspring indirectly via the rearing environment, or “genetic nurture”^6^. Distinguishing DGEs from IGEs is essential for determining the extent to which polygenic predictions reflect causal effects of inherited alleles, as well as understanding the mechanisms underlying the intergenerational transmission of neurodevelopmental risks.

Several complementary family genomic approaches have been developed to estimate direct and indirect genetic effects by exploiting the random segregation of alleles during meiosis. The sibling-comparison design uses genetic differences among genotyped siblings or dizygotic (DZ) twins to predict corresponding differences in phenotype. Because siblings inherit a random subset of parental alleles, this estimand is unconfounded by IGEs. The trio design uses samples comprising genotyped mother-father-child trios, regressing offspring phenotype on offspring genotype while conditioning on parental genotype. Because parental genotype is held constant, the resulting estimate reflects only the within-family segregation of alleles, meaning both the sibling-comparison and trio design have the same estimand.

It has been noted, however, that the sibling comparison approach is subject to potential bias and limitations. In particular, sibling-based PGS models do not necessarily uncover the estimates of direct genetic effects, even in the absence of indirect genetic effects^7^. Within-family estimates can be attenuated or inflated due to the correlation structure of environmental influences between siblings and the extent to which PGS captures a mixture of direct and indirect genetic components.

In comparison, the trio-based PGS model is less biased and more powerful, but relatively few cohorts have genotyped trios. Prior work has demonstrated that missing parental genotypes can partially be reconstructed using Mendelian imputation in parent-offspring and sibling pairs^8,9^, thereby increasing the power of trio-based PGS analysis.

Despite these methodological advances, few studies have tested for IGEs on neurodevelopmental traits and the evidence is mixed. Sibling-comparison studies have identified IGEs on educational and cognitive outcomes, but few on psychiatric traits^10,11^. Cross-trait IGEs on parent-reported adolescent ADHD symptoms have been identified using a self-rated general health PGS, although age-specific effects were not examined^10^. Another study examined externalising PGS associations across development and reporters, finding stable IGEs on conduct problems, but not on ADHD traits^12^. Evidence from sibling-comparison studies on other neurodevelopmental traits remains sparse.

Trio-based PGS analyses of parent-reported neurodevelopmental traits at age 3 identified cross-trait IGEs of maternal ASD PGS on hyperactivity and language difficulties, but no within-trait IGEs^13^. Similarly, associations between parental ADHD PGS and mother-reported ADHD traits at age 8 were largely explained by DGEs, although maternal neuroticism PGS showed a cross-trait IGE^14^. Another study found robust DGEs of child ADHD PGS on mother-reported hyperactivity and inattention, with little evidence for IGEs of parental ADHD PGS but modest cross-trait IGEs from other parental psychiatric PGS^15^.

Taken together, evidence for IGEs on neurodevelopmental traits remains limited. However, most studies have focused on early childhood, a single cross-sectional assessment and/or a single PGS. Existing studies have also relied on a single analytical approach. No previous study has triangulated across designs or made comparisons across developmental stages, reporters, or neurodevelopmental traits. Finally, current evidence is largely restricted to ADHD-related outcomes, leaving it unclear whether similar patterns extend to other neurodevelopmental outcomes.

In the present study, we estimated DGEs and IGEs on ADHD and ASD traits across ages and reporters using both twin-comparison and trio-based PGS analyses in the Twins Early Development Study (TEDS). We incorporated Mendelian imputation of parental genotypes to maximise power in trio analyses and examined reporter-specific cross-developmental latent factors to determine whether indirect genetic effects differ across developmental stages and informants on stable trait indices. A summary and comparison between the two methods for calculating IGE is shown in Supplementary Table S1.

Specifically, we asked (1) whether child PGS predict ADHD and ASD traits in DZ twin pairs within and between families across development; (2) whether trio-based PGS analyses provide evidence for direct and indirect genetic effects on the same outcomes; and (3) whether these findings differ on reporter-specific and cross-reporter latent factors capturing stable variation over time.

## Results

### Twin-comparison Analyses

We examined ADHD and ASD traits across development and reporters (Supplementary Table S2). Full estimates of all models, including FDR-adjusted q values, are reported in Supplementary Tables S3-S8.

We first examined DGEs and IGEs using twin-comparison models that contrasted population-level and within-family PGS associations (Figures 1 and 2). Twin-comparison analyses provided consistent evidence for within-trait DGEs on both ADHD and ASD across development. For ADHD, within-trait DGEs were observed from ages 4 to 21 across parent-, teacher- and self-reported measures (within-family β = 0.08-0.18), although there was no evidence for a DGE on self-reported symptoms at age 26. Evidence for within-trait IGEs was limited, with only small negative effects observed for parent-reported ADHD symptoms at ages 4 and 7 (β = −0.05), neither surviving FDR correction.

**Figure 1.**
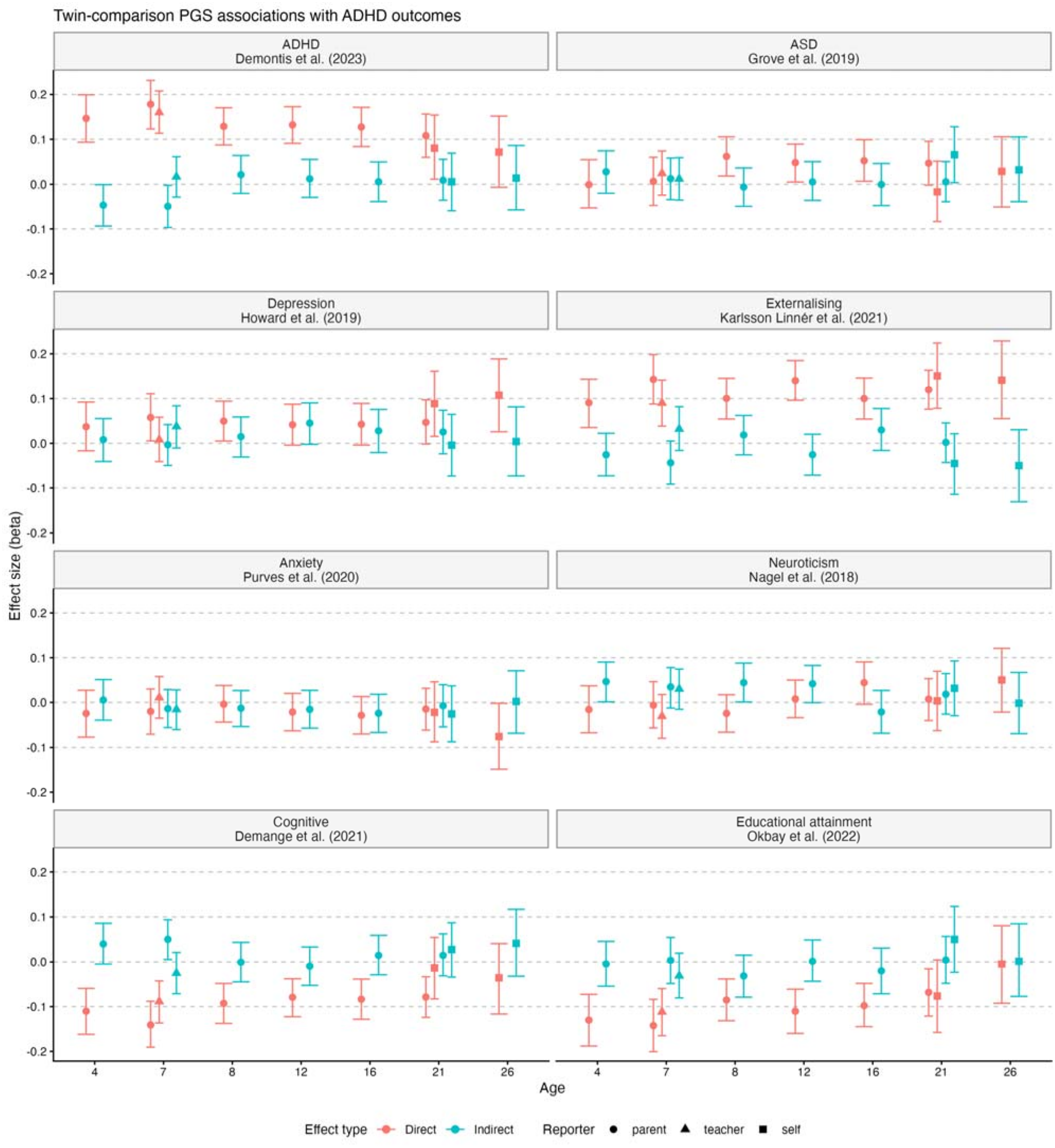
Twin-comparison PGS associations with ADHD outcomes for eight PGS

**Figure 2.**
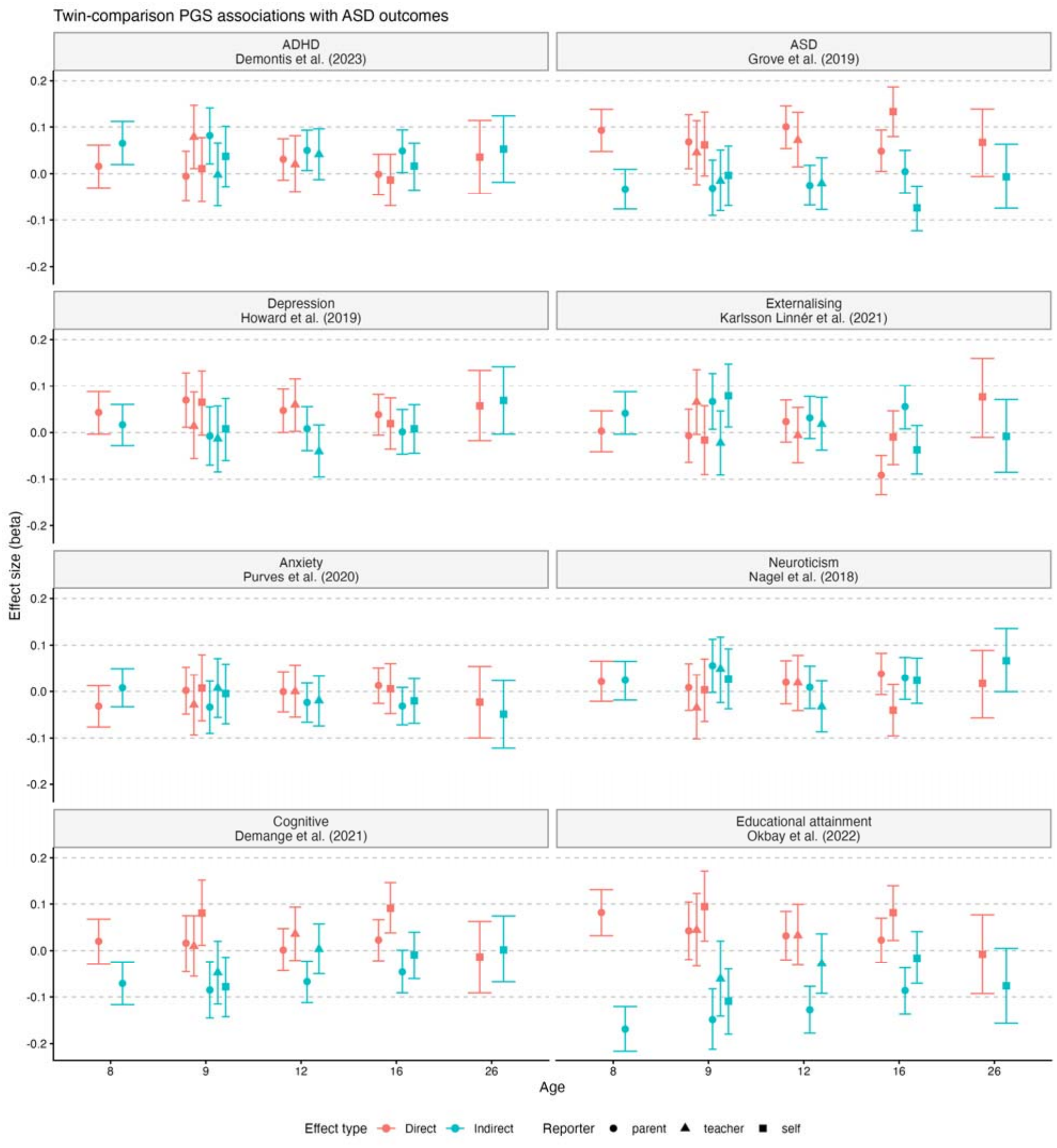
Twin-comparison PGS associations with ASD outcomes for eight PGS

A similar pattern was observed in within-trait analysis on ASD traits (Figure 2). Within-trait DGEs were observed across most measures from ages 8 to 16 (β = 0.05-0.13) but not for self-reported ASD traits at age 26. Evidence for within-trait IGEs was limited to self-reported ASD traits at age 16 (β = −0.07), which survived FDR correction.

Beyond within-trait associations, several cross-trait DGEs were also observed. Depression PGS predicted ADHD traits in childhood and early adulthood, whereas ASD, cognition, educational attainment (EA) and externalising PGS showed direct associations across multiple developmental stages. Cross-trait IGEs were limited to cognition and neuroticism PGS (parent-reported, childhood) and ASD PGS (self-reported, age 21), none surviving FDR correction. For ASD outcomes, cross-trait DGEs were identified for depression, cognition, ADHD, externalising and EA PGS at specific developmental stages. Cross-trait IGEs were evident for ADHD, externalising, cognition and EA PGS, primarily for parent-reported outcomes (strongest for EA PGS, β = −0.09 to −0.17); the cognition and EA effects largely survived FDR correction.

### Trio-PGS Analyses

We next examined DGEs and IGEs using trio-PGS models including child, maternal and paternal PGS (Figures 3 and 4). Consistent with the twin-comparison analyses, DGEs were considerably more consistent and stronger than IGEs, which were mainly observed in cross-trait associations. Within-trait associations provided evidence for consistent DGEs. Child ADHD PGS predicted ADHD traits across all developmental stages (β = 0.07-0.17), and child ASD PGS predicted ASD traits across most assessments (β = 0.06-0.11), with the exception of age 26. Evidence for within-trait IGEs was limited, and did not survive FDR correction.

**Figure 3.**
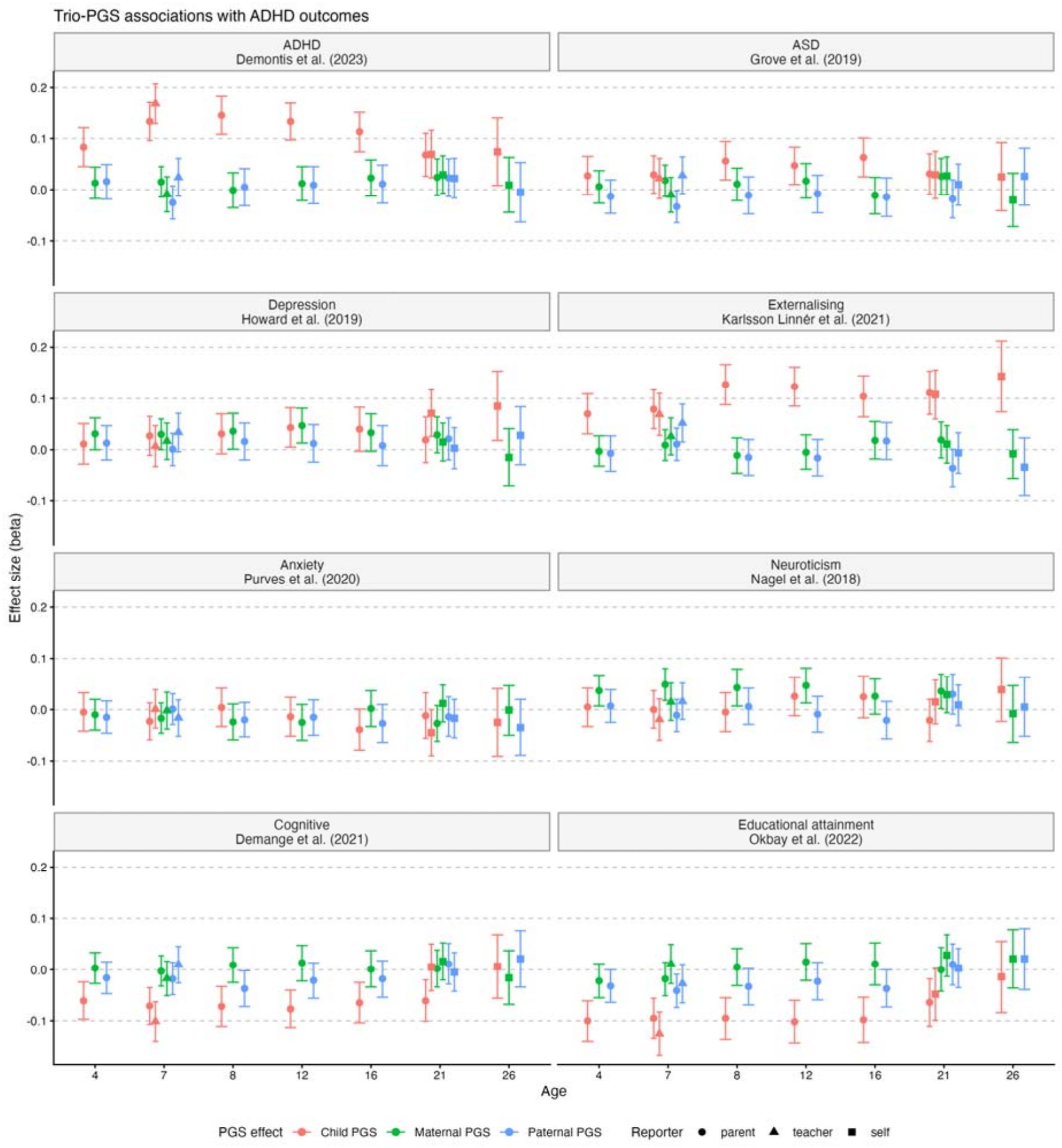
Trio-PGS associations between eight selected PGS and ADHD outcomes

**Figure 4.**
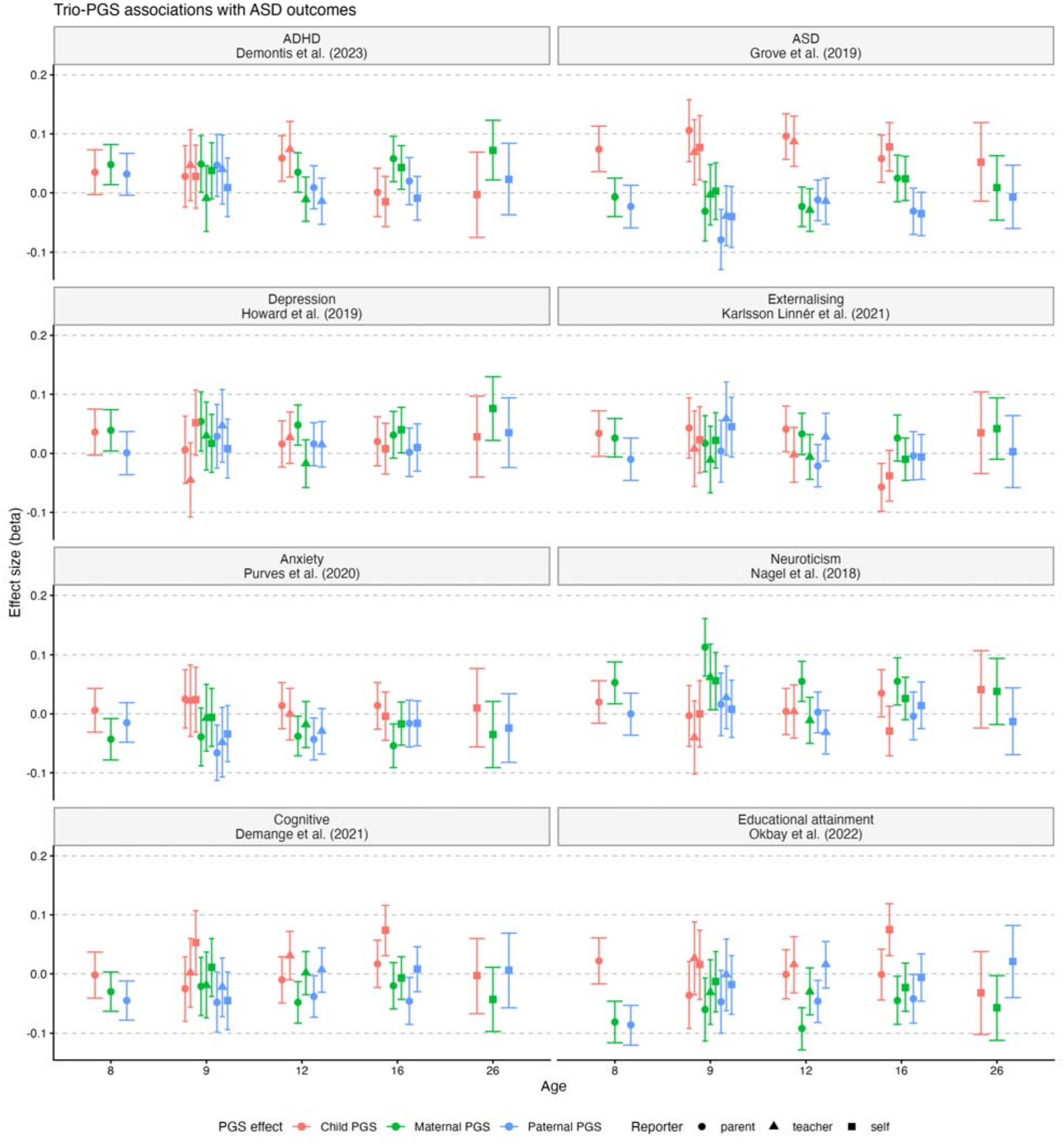
Trio-PGS associations between eight selected PGS and ASD outcomes

Beyond within-trait associations, several cross-trait DGEs were identified. For ADHD outcomes, child depression PGS was predictive at ages 12, 21 and 26. Child ASD, cognitive ability, EA and externalising PGS showed direct associations across multiple developmental stages. For ASD outcomes, child ADHD, cognition and EA PGS also showed positive direct associations. In contrast, externalising PGS was positively associated with parent-report ASD trait at age 12, but negatively associated with parent-report outcome at age 16.

Cross-trait IGEs were more heterogeneous. For ADHD outcomes, maternal depression and neuroticism PGS and paternal ASD, cognitive, EA and externalising PGS showed indirect associations at specific developmental stages, none of which survived FDR correction. For ASD outcomes, IGEs were observed mainly for parent-reported traits: positive for maternal ADHD, depression and neuroticism PGS, and negative for maternal and paternal anxiety, cognitive and EA PGS. After FDR correction, IGEs on ASD outcomes were retained mainly for maternal ADHD, depression, anxiety, neuroticism, cognitive and EA PGS, and for paternal EA PGS (|β| = 0.05-0.11).

### Reporter-specific and cross-developmental latent factors

To capture stable trait factors, we calculated three reporter-specific cross-development latent factors (parent-, teacher- and self-report) and cross-reporter, cross-development common factors for ADHD and ASD (Figures 5 and 6). Twin-comparison analyses again provided consistent evidence for within-trait DGEs on both ADHD and ASD latent factors (β = 0.07-0.16), with no evidence for IGEs. Cross-trait DGEs were observed for both ADHD and ASD latent factors across multiple psychiatric and cognition PGS. While no cross-trait IGEs were identified for ADHD latent factors, ASD factors showed some IGEs from ADHD, cognition, EA and externalising PGS, predominantly for the parent-reported and common factors; all except externalising survived FDR correction.

**Figure 5.**
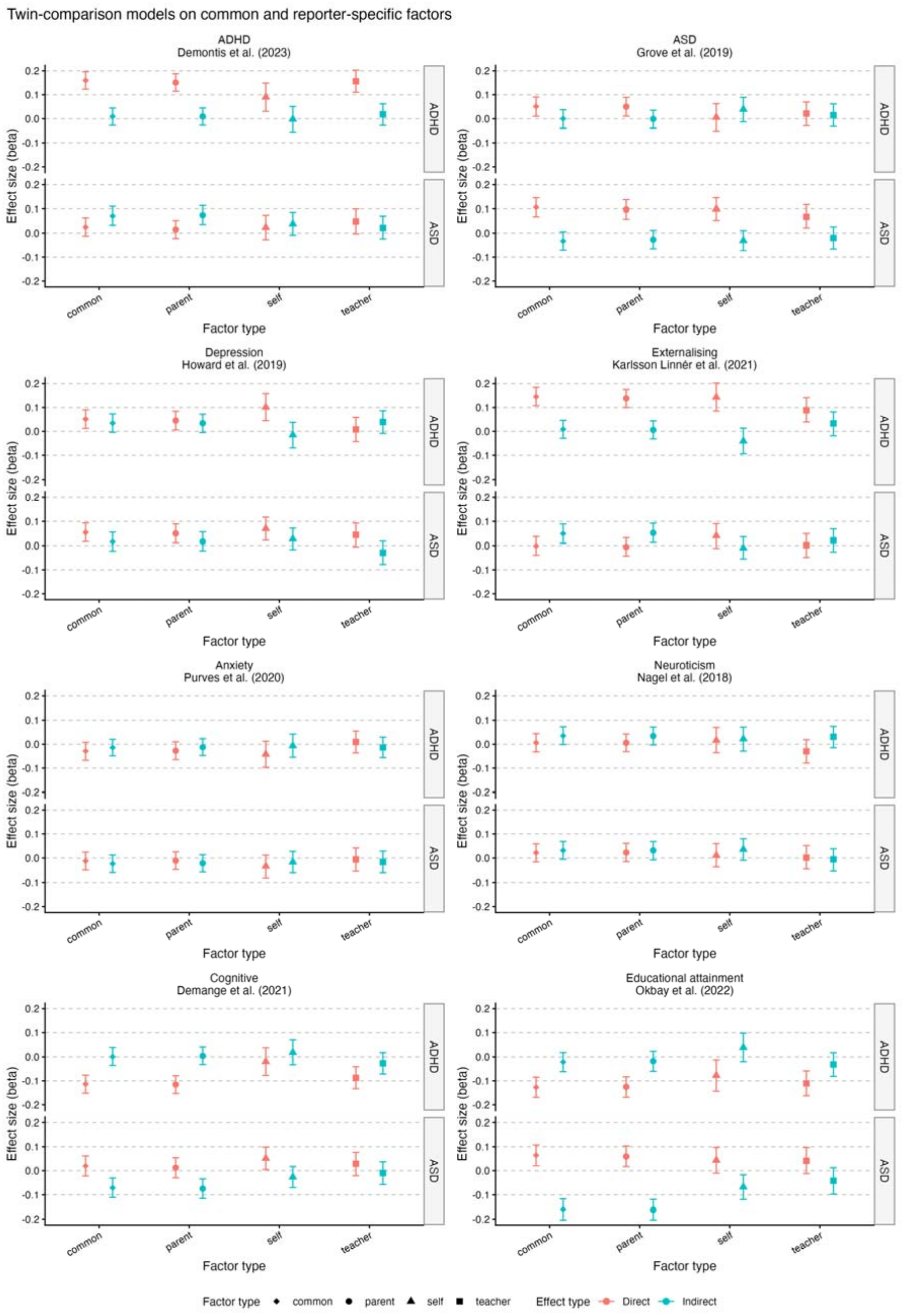
Twin-comparison analyses on reporter-specific and common latent factors for eight PGS

**Figure 6.**
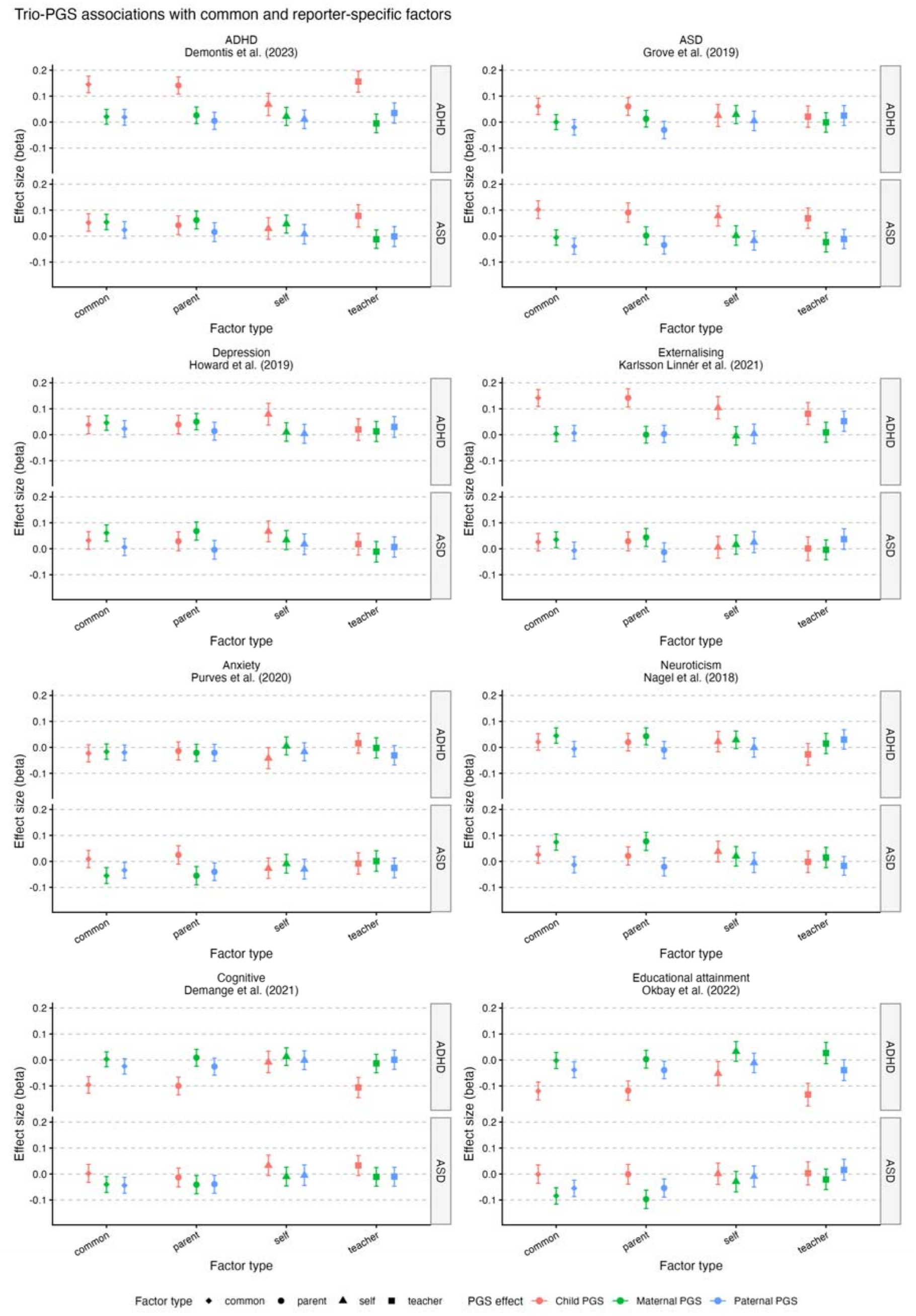
Trio-PGS associations with reporter-specific and common latent factors for eight PGS

Trio-PGS analyses similarly demonstrated significant within-trait DGEs (β = 0.07-0.16). Evidence for within-trait IGEs remained limited, with only paternal ASD PGS predicting the common ASD factor, which did not survive FDR correction. Several cross-trait DGEs were observed. ADHD latent factors were associated with ASD, cognition, depression, EA and externalising PGS, whereas ASD latent factors were associated with ADHD and depression PGS. Cross-trait IGEs were again limited and different between mothers and fathers. For ADHD, indirect effects were observed for maternal depression and neuroticism PGS, together with paternal EA and externalising PGS; only the maternal effects survived FDR correction. For ASD, cross-trait IGEs were identified across maternal ADHD, depression, cognition, EA, externalising and neuroticism PGS, as well as paternal EA and cognition PGS, although these associations were largely restricted to the parent-reported and common factors. Maternal and paternal anxiety PGS again showed negative IGEs (maternal only after FDR correction).

Together, these findings largely replicated those from individual developmental outcomes across age and reporter, indicating that robust DGEs remained after reducing measurement error, whereas IGEs remained comparatively small and inconsistent. Notably, several DGEs and IGEs were evident only for the parent-reported and common latent factors, with little evidence for teacher- or self-reported factors.

## Discussion

Across eight waves of data spanning childhood to adulthood, using data from three reporters, and applying two complementary family-genomic designs, we found consistent evidence that polygenic associations with ADHD and ASD traits were driven predominantly by direct genetic effects. These effects were evident across childhood, adolescence and early adulthood, across parent, teacher and self-report, and in latent factors capturing stability across reporters and development. In contrast, within-trait indirect genetic effects were rare and inconsistent. Only one within-trait IGE survived FDR correction. Cross-trait DGEs and IGEs were observed across multiple neurodevelopmental, psychiatric and cognitive PGS.

Our findings provide systematic evidence that within-trait polygenic associations on neurodevelopmental traits are predominantly attributable to DGEs, with little consistent evidence for IGEs. These align with some previous studies on childhood neurodevelopmental traits^13,14^, while extending this evidence across a substantially wider developmental period and multiple reporters. Our findings also add to a growing but mixed literature on IGEs across psychiatric traits. While IGEs have been relatively consistently observed for educational and cognitive outcomes, evidence for psychiatric traits has been more heterogeneous, with IGEs for some externalising and internalising phenotypes but not consistently replicated across studies^16,17^. Rather than indicating a categorical distinction between neurodevelopmental and psychiatric conditions, our findings suggest that the relative contribution of DGEs and IGEs varies across traits. ADHD and ASD symptoms may be more closely tied to offspring-intrinsic genetic risk, whereas outcomes such as EA and internalising problems may additionally reflect more environmentally mediated pathways correlated with parental genotype.

In line with previous work^12,15^, cross-trait findings further provided direct and indirect genetic evidence for the frequent comorbidity of neurodevelopmental, psychiatric, and cognitive conditions. Across both approaches, ADHD outcomes showed direct associations with ASD, depression, cognition, EA and externalising PGS, whereas ASD outcomes were associated with ADHD, cognition and EA PGS, suggesting that genetic liabilities related to psychiatric and cognitive traits also contribute directly to neurodevelopmental symptoms. Cross-trait IGEs of ADHD, cognition and EA PGS on ASD traits were also consistent across both approaches and robust to FDR correction, supporting the existence of environmentally mediated pathways beyond disorder-specific genetic transmission. Interestingly, these indirect effects were largely limited to parent-reported outcomes and the common latent factor, with little evidence in teacher- or self-reported measures. One possibility is that this pattern reflects reporter effects, whereby parental genotype influences parents’ perceptions or reporting of their child’s behaviour. Alternatively, these indirect effects may reflect environmental pathways that are specific to the home. Parental genotypes may shape aspects of family life, such as household routines, parent–child conflict or how homework and bedtimes are managed, and children’s behavioural responses to these may be most evident at home rather than at school. Moreover, robust DGEs were observed across parent-, teacher- and self-reported outcomes, suggesting that parent ratings captured meaningful child symptoms rather than simply reflecting parental reporting bias.

We also observed unexpected negative IGEs, whereby higher parental ASD and anxiety PGS were associated with lower offspring ASD traits. These associations were small and inconsistent, and several did not survive FDR correction; they should therefore be interpreted with caution. These patterns were not observed in previous studies examining IGEs of ASD liability on child social communication traits at age 3^13^, raising the possibility that negative IGEs may emerge later in development. Previous studies have reported negative covariance between offspring direct and parental indirect genetic effects for childhood externalising traits^18^ and depression^19^. Although different from negative PGS-based indirect associations, these findings also suggest that direct and indirect genetic pathways may act in opposing directions. One possibility is that parental ASD or anxiety-related behavioural tendencies contribute to structured routines, earlier recognition of difficulties or other parental behaviours that support children, although these mechanisms were not directly tested. Selection offers an alternative explanation: van den Berg et al. found that negative correlations between direct genetic effects and nontransmitted coefficients for fluid intelligence in UK Biobank weakened when ascertainment bias was reduced^20^. Recent work in TEDS found that attrition had little impact on genetic and environmental variance components, providing some reassurance, although this does not exclude selection bias in the present PGS analyses^21^. Replication and direct assessment of potential mediating behaviours are therefore needed.

Several limitations should be considered. Twin-comparison and trio-PGS approaches rely on different assumptions and capture partially distinct components of IGEs, particularly with respect to non-transmitted alleles. However, the consistency of main findings between approaches increases confidence in the overall conclusions. We used Mendelian imputation for many parental genotypes, which may have influenced the estimation despite previous validation of the imputation methodology. Overall, however, the convergence of results indicates that direct genetic effects dominate polygenic prediction of ADHD and ASD across development, whereas IGEs are rarer, smaller, and less consistent. Replication in independent cohorts with directly genotyped parents and increasingly predictive polygenic scores will be important for extending the robustness of these findings.

## Methods

### Sample

We analysed data from the Twins Early Development Study (TEDS), a longitudinal cohort of twins born in England and Wales between 1994 and 1996^22^. Standard TEDS exclusion criteria were applied, and twins with available genotype data were included. Recent work has shown that the genotyped TEDS sample is broadly representative of the full cohort, with minimal evidence of selection bias^21^. Twin-comparison analyses were restricted to dizygotic (DZ) twins (n=6614), whereas trio-based analyses included both monozygotic (MZ) and DZ twins (n=7259). Ethical approval for TEDS was obtained from King’s College London (HR/DP-20/21-22060).

### Phenotypic Measurements

ADHD and ASD related traits were assessed repeatedly from early childhood to young adulthood using age-appropriate, well-validated instruments (age 4, 7, 8, 9, 12, 16, 21, 26). ADHD traits were measured using the Strengths and Difficulties Questionnaire (SDQ)^23^ and the Conners’ Rating Scales^24^. ASD traits were assessed using the Childhood Autism Spectrum Test (CAST)^25^, Autism Spectrum Quotient (AQ)^26^, and Ritvo Autism Asperger Diagnostic Scale (RAADS)^27^. Depending on developmental stage, ratings were obtained from parents, teachers and participants themselves. Supplementary Table S2 provides an overview of the questionnaires and informants across ages.

### Covariates

We adjusted all models for child age, sex, genotyping chip and the first ten genetic principal components.

### Polygenic scores

Polygenic scores were calculated using LDpred2-auto^28^ for children and parents using available TEDS genotype data and imputed parental genotype data. Based on published genome-wide association studies, we included 8 PGS for ADHD^29^, ASD^30^, anxiety^31^, depression^32^, educational attainment^33^, cognition^34^, neuroticism^35^, and externalising problems^36^.

### Parental genotype imputation

Missing parental genotypes were imputed using SNIPAR v0.0.22,^9^ which applies Mendelian inheritance within families to infer the expected genotypes of unobserved parents. Briefly, offspring genotypes contain alleles transmitted by their parents. When multiple siblings are genotyped, identity-by-descent (IBD) sharing between siblings provides additional information about the parental haplotypes from which these alleles were inherited. Similarly, when one parent and their offspring are genotyped, the alleles transmitted by the ungenotyped parent are observed in the offspring, while the remaining untransmitted alleles remain unknown and are inferred using population allele frequencies. In the case where sibling pairs are genotyped, but not their parents, SNIPAR imputes the sum of the maternal and paternal genotypes. In genotyped parent-offspring pairs, half of the ungenotyped parent is observed in the offspring, while the remaining alleles are imputed using population frequency.

Following standard genotype quality-control procedures (autosomal, biallelic SNPs with imputation INFO > 0.99 and Hardy-Weinberg equilibrium mid-*P* > 1 × 10), one member of each monozygotic twin pair was randomly excluded, in line with SNIPAR recommendations. The remaining sample was then phased using SHAPEIT5 v5.1.1. Sibling IBD sharing was then inferred from the phased haplotypes using SNIPAR’s *ibd.py* function. Lastly, we used SNIPAR’s *impute.py* function to carry out parental genotype imputation, resulting in 7,259 trios. Both parental genotypes were directly measured for 1,240 participants (17.1%), one parental genotype was measured and the other imputed for 2,044 (28.2%), and neither parental genotype was directly measured for 3,975 (54.8%). Polygenic scores for offspring and observed or imputed parents were subsequently calculated from LDpred2-auto weights using SNIPAR’s *pgs.py*.

### Statistical analyses

#### Twin-comparison models

Direct genetic effects were estimated using within-family regression models following previous studies^12,37^. For each phenotype-PGS combination, we fitted (i) a population model including offspring PGS and (ii) a within-between model including the twin mean PGS (between-family effect) and the child-specific deviation from the mean (within-family effect). The within-family coefficient was interpreted as the DGE, and the IGE was calculated as the difference between the population and within-family estimates. Standard errors and percentile 95% confidence intervals for all estimates were obtained by non-parametric bootstrapping (5,000 resamples), and two-sided p values were derived from z = β/SE.

#### Trio-PGS models

We fitted trio-based regression models including child, maternal and paternal PGS. Child PGS effects were interpreted as estimates of DGEs conditional on parental genotype, whereas maternal and paternal PGS effects represented IGEs after adjustment for offspring genotype. Trio-PGS models included both members of MZ and DZ twin pairs; standard errors were clustered by family (HC1) to account for non-independence.

#### Common and reporter-specific latent factors

Frequently, researchers are interested in trait stability over development and/or across informants, as focussing on such stability can remove time-specific variation and error. To investigate IGE and DGE estimates on stability, confirmatory factor analyses were conducted.

For ADHD and ASD separately, we fitted single-factor CFAs to scale scores (i) across all ages and reporters (common factor) and (ii) across ages within each reporter (reporter-specific factors; loadings constrained equal for two-indicator factors), using robust maximum likelihood with FIML for missing data.^38^ Factor scores were extracted using lavPredict and used as outcomes. Twin-comparison and trio-based analyses were repeated using these latent factor scores.

#### Multiple testing

We controlled the false discovery rate (FDR) using the Benjamini-Hochberg procedure, separately for each estimate type within each set of analyses (Supplementary Tables S3-S8). We report nominally significant associations (p < .05) and indicate those surviving FDR correction (q < .05).

## Data availability

The TEDS data used in this study are available through an application to the TEDS team, subject to approval and applicable data access conditions.

## Code availability

The code used to conduct the analyses will be made available on the Open Science Framework upon publication.

## Notes

### Competing Interest Statement

The authors have declared no competing interest.

### Author Declarations

Ethical approval for TEDS was obtained from King's College London (HR/DP-20/21-22060). All TEDS datasets used in this study had been de-identified prior to being made available to us.

## References

1. Jensen M, Girirajan S. Mapping a shared genetic basis for neurodevelopmental disorders. Genome Medicine. 2017;9(1):1–3. doi:10.1186/S13073-017-0503-4

2. Faraone SV, Larsson H. Genetics of attention deficit hyperactivity disorder. Mol Psychiatry. 2019;24(4):562–575. doi:10.1038/s41380-018-0070-0

3. Farhat LC, Brentani H, de Toledo VHC, et al. ADHD and autism symptoms in youth: a network analysis. Journal of child psychology and psychiatry, and allied disciplines. 2022;63(2):143–151. doi:10.1111/JCPP.13436

4. Ronald A, de Bode N, Polderman TJC. Systematic Review: How the Attention-Deficit/Hyperactivity Disorder Polygenic Risk Score Adds to Our Understanding of ADHD and Associated Traits. J Am Acad Child Adolesc Psychiatry. 2021;60(10):1234–1277. doi:10.1016/j.jaac.2021.01.019

5. Pingault JB, Allegrini AG, Odigie T, et al. Research Review: How to interpret associations between polygenic scores, environmental risks, and phenotypes. Journal of Child Psychology and Psychiatry. 2022;63(10):1125–1139. doi:10.1111/jcpp.13607

6. Kong A, Thorleifsson G, Frigge ML, et al. The nature of nurture: Effects of parental genotypes. Science. 2018;359(6374):424–428. doi:10.1126/science.aan6877

7. Fletcher J, Wu Y, Li T, Lu Q. Interpreting polygenic score effects in sibling analysis. PLOS ONE. 2024;19(2):e0282212. doi:10.1371/journal.pone.0282212

8. Hwang LD, Tubbs JD, Luong J, et al. Estimating indirect parental genetic effects on offspring phenotypes using virtual parental genotypes derived from sibling and half sibling pairs. PLoS Genetics. 2020;16(10):e1009154. doi:10.1371/journal.pgen.1009154

9. Young AI, Nehzati SM, Benonisdottir S, et al. Mendelian imputation of parental genotypes improves estimates of direct genetic effects. Nature Genetics. 2022;54(6):897. doi:10.1038/s41588-022-01085-0

10. Selzam S, Ritchie SJ, Pingault JB, Reynolds CA, O’Reilly PF, Plomin R. Comparing Within- and Between-Family Polygenic Score Prediction. Am J Hum Genet. 2019;105(2):351–363. doi:10.1016/j.ajhg.2019.06.006

11. Van Der Laan CM, Van De Weijer SGA, Pool R, et al. Direct and Indirect Genetic Effects on Aggression. Biological Psychiatry Global Open Science. 2023;3(4):958–968. doi:10.1016/j.bpsgos.2023.04.006

12. Bright JK, Rayner C, Freeman Z, et al. Using twin-pairs to assess potential bias in polygenic prediction of externalising behaviours across development. Mol Psychiatry. Published online February 19, 2025. doi:10.1038/s41380-025-02920-6

13. Hegemann L, Eilertsen E, Hagen Pettersen J, et al. Direct and indirect genetic effects on early neurodevelopmental traits. Journal of Child Psychology and Psychiatry. 2025;66:1053–1064. doi:10.1111/jcpp.14122

14. Pingault JB, Barkhuizen W, Wang B, et al. Genetic nurture versus genetic transmission of risk for ADHD traits in the Norwegian Mother, Father and Child Cohort Study. Mol Psychiatry. 2023;28(4):1731–1738. doi:10.1038/s41380-022-01863-6

15. Allegrini AG, Hannigan LJ, Frach L, et al. Intergenerational transmission of polygenic predisposition for neuropsychiatric traits on emotional and behavioural difficulties in childhood. Nat Commun. 2025;16(1):2674. doi:10.1038/s41467-025-57694-w

16. Chegeni R, Nes RB, Cheesman R, Ayorech Z, Ystrøm E, Røysamb E. Direct and indirect parental genetic effects on offspring susceptibility to internalizing problems across development. Nat Mental Health. 2026;4(5):754–765. doi:10.1038/s44220-026-00619-3

17. Trindade Pons V, Oldehinkel AJ, van Loo HM. Genetic nurture effects in depressive and anxiety disorders and symptoms, and in related traits. Mol Psychiatry. 2025;30(12):5694–5700. doi:10.1038/s41380-025-03265-w

18. Eilertsen EM, Cheesman R, Ayorech Z, et al. On the importance of parenting in externalizing disorders: an evaluation of indirect genetic effects in families. Journal of Child Psychology and Psychiatry. 2022;63(10):1186–1195. doi:10.1111/jcpp.13654

19. Cheesman R, Eilertsen EM, Ahmadzadeh YI, et al. How important are parents in the development of child anxiety and depression? A genomic analysis of parent-offspring trios in the Norwegian Mother Father and Child Cohort Study (MoBa). BMC Med. 2020;18(1):284. doi:10.1186/s12916-020-01760-1

20. van den Berg DM, Malawsky DS, Huang W, et al. Imputation of fluid intelligence scores reduces ascertainment bias and increases power for analyses of common and rare variants. Nat Genet. Published online October 5, 2026:1–10. doi:10.1038/s41588-026-02787-5

21. Bignardi G, Eley TC, Lockhart CL, Rayner C, Tilling K, McAdams T. Attrition in the Twins Early Development Study: predictors, consequences, and corrections. PsyArXiv. Preprint posted online August 24, 2026:ex276_v1. Accessed October 24, 2026. https://osf.io/preprints/psyarxiv/ex276_v1/

22. Lockhart C, Bright J, Ahmadzadeh Y, et al. Twins Early Development Study (TEDS): A genetically sensitive investigation of mental health outcomes in the mid-twenties. JCPP Adv. 2023;3(2):e12154. doi:10.1002/jcv2.12154

23. Goodman. The Strengths and Difficulties Questionnaire_ A Research Note - Goodman - 1997 - Journal of Child Psychology and Psychiatry - Wiley Online Library. J Child Psychol Psychiat. 1997;38(5):581–586.

24. Conners CK, Pitkanen J, Rzepa SR. Conners 3rd Edition (Conners 3; Conners 2008). In: Encyclopedia of Clinical Neuropsychology. Springer, New York, NY; 2011:675–678. doi:10.1007/978-0-387-79948-3_1534

25. Scott FJ, Baron-Cohen S, Bolton P, Brayne C. The CAST (Childhood Asperger Syndrome Test): preliminary development of a UK screen for mainstream primary-school-age children. Autism. 2002;6(1):9–31. doi:10.1177/1362361302006001003

26. Baron-Cohen S, Wheelwright S, Skinner R, Martin J, Clubley E. The autism-spectrum quotient (AQ): evidence from Asperger syndrome/high-functioning autism, males and females, scientists and mathematicians. J Autism Dev Disord. 2001;31(1):5–17. doi:10.1023/a:1005653411471

27. Ritvo RA, Ritvo ER, Guthrie D, et al. The Ritvo Autism Asperger Diagnostic Scale-Revised (RAADS-R): a scale to assist the diagnosis of Autism Spectrum Disorder in adults: an international validation study. J Autism Dev Disord. 2011;41(8):1076–1089. doi:10.1007/s10803-010-1133-5

28. Privé F, Arbel J, Vilhjálmsson BJ. LDpred2: better, faster, stronger. Bioinformatics. 2020;36(22-23):5424–5431. doi:10.1093/bioinformatics/btaa1029

29. Demontis D, Walters GB, Athanasiadis G, et al. Genome-wide analyses of ADHD identify 27 risk loci, refine the genetic architecture and implicate several cognitive domains. Nat Genet. 2023;55(2):198–208. doi:10.1038/s41588-022-01285-8

30. Grove J, Ripke S, Als TD, et al. Identification of common genetic risk variants for autism spectrum disorder. Nat Genet. 2019;51(3):431–444. doi:10.1038/s41588-019-0344-8

31. Purves KL, Coleman JRI, Meier SM, et al. A major role for common genetic variation in anxiety disorders. Mol Psychiatry. 2020;25(12):3292–3303. doi:10.1038/s41380-019-0559-1

32. Howard DM, Adams MJ, Clarke TK, et al. Genome-wide meta-analysis of depression identifies 102 independent variants and highlights the importance of the prefrontal brain regions. Nat Neurosci. 2019;22(3):343–352. doi:10.1038/s41593-018-0326-7

33. Okbay A, Wu Y, Wang N, et al. Polygenic prediction of educational attainment within and between families from genome-wide association analyses in 3 million individuals. Nat Genet. 2022;54(4):437–449. doi:10.1038/s41588-022-01016-z

34. Demange PA, Malanchini M, Mallard TT, et al. Investigating the genetic architecture of noncognitive skills using GWAS-by-subtraction. Nat Genet. 2021;53(1):35–44. doi:10.1038/s41588-020-00754-2

35. Nagel M, Jansen PR, Stringer S, et al. Meta-analysis of genome-wide association studies for neuroticism in 449,484 individuals identifies novel genetic loci and pathways. Nat Genet. 2018;50(7):920–927. doi:10.1038/s41588-018-0151-7

36. Karlsson Linnér R, Mallard TT, Barr PB, et al. Multivariate analysis of 1.5 million people identifies genetic associations with traits related to self-regulation and addiction. Nat Neurosci. 2021;24(10):1367–1376. doi:10.1038/s41593-021-00908-3

37. Demange PA, Hottenga JJ, Abdellaoui A, et al. Estimating effects of parents’ cognitive and non-cognitive skills on offspring education using polygenic scores. Nat Commun. 2022;13(1):4801. doi:10.1038/s41467-022-32003-x

38. Rosseel Y. lavaan : An R Package for Structural Equation. Journal of Statistical Software. 2012;48(2).

