## Supplementary materials for "Investigating Indirect Genetic Effects on Neurodevelopmental Traits Across Development and Reporters: Triangulating Twin-comparison and Trio-PGS Approaches"

### Supplementary Table S1. Summary of statistical approaches and their key features

| Feature | Twin-comparison design | Trio-PGS design with imputed parental genotypes |
| --- | --- | --- |
| Core structure | DZ twin pairs with offspring PGS and phenotypes | Parent–offspring trios with offspring PGS and measured or imputed parental PGS |
| Direct genetic effect | Within-family effect of the child’s PGS | Effect of the child’s own PGS on their outcome, conditional on parental PGS |
| Indirect genetic effect | Population-level effect minus within-family effect | Effect of parental PGS on the child’s outcome, conditional on child PGS |
| Genetic variation captured | Transmitted alleles (shared between parent and child) | Transmitted and non-transmitted alleles (captured by parental PGS) |
| Source of indirect effects | Environmental pathways shaped by parental genotype, captured via between-family PGS effects | Environmental pathways from parental genotype, independent of child genotype |
| Required data | Genotype and phenotype of DZ twins (no parental genotype required) | Child genotype and phenotype, and measured or imputed parental genotype |
| Limitations | Cannot capture non-transmitted alleles; assumes that indirect genetic effects are shared within families; limited by PGS predictive power | Requires trio genotyping or high-quality imputation; limited by PGS predictive power |

**Notes:** DZ, dizygotic; PGS, polygenic score.

### Supplementary Table S2. Overview of longitudinal measures

| Table S2. Overview of longitudinal measures | | | |
| --- | --- | --- | --- |
| NDD trait | Questionnaire | age | reporter |
| ADHD | SDQ | 4 | parent |
|  | SDQ | 7 | parent |
|  | SDQ | 7 | teacher |
|  | Conners | 8 | parent |
|  | Conners | 12 | parent |
|  | Conners | 16 | parent |
|  | Conners | 21 | parent |
|  | Conners | 21 | self |
|  | Conners | 26 | self |
| ASD | CAST | 8 | parent |
|  | CAST | 9 | parent |
|  | CAST | 9 | self |
|  | CAST | 9 | teacher |
|  | CAST | 12 | parent |
|  | CAST | 12 | teacher |
|  | AQ | 16 | parent |
|  | AQ | 16 | self |
|  | RAADS | 26 | self |
| Note: sample sizes vary across assessments because of differences in phenotype availability. SDQ, Strengths and Difficulties Questionnaire; Conners, Conners’ Rating Scales; CAST, Childhood Autism Spectrum Test; AQ, Autism Spectrum Quotient; RAADS, Ritvo Autism and Asperger Diagnostic Scale. | | | |

### Supplementary Table S3. Twin-comparison analyses of ADHD traits

#### ADHD PGS · Demontis et al. (2023)

| Outcome (age, reporter) | N (offspring) | Families (clusters) | Population (β, 95% CI, SE, *p*, *q*) | Direct (β, 95% CI, SE, *p*, *q*) | Between (β, 95% CI, SE, *p*, *q*) | Indirect (β, 95% CI, SE, *p*, *q*) |
| --- | --- | --- | --- | --- | --- | --- |
| Age 4 Parent | 5,180 | 2,610 | 0.099 [0.073, 0.126] SE 0.014 *p* < .001 *q* < .001 | 0.146 [0.094, 0.199] SE 0.027 *p* < .001 *q* < .001 | 0.085 [0.053, 0.116] SE 0.016 *p* < .001 *q* < .001 | −0.047 [−0.094, −0.001] SE 0.024 *p* = .048 *q* = .469 |
| Age 7 Parent | 5,317 | 2,659 | 0.128 [0.101, 0.155] SE 0.014 *p* < .001 *q* < .001 | 0.178 [0.123, 0.231] SE 0.028 *p* < .001 *q* < .001 | 0.113 [0.082, 0.143] SE 0.015 *p* < .001 *q* < .001 | −0.050 [−0.097, −0.002] SE 0.024 *p* = .039 *q* = .469 |
| Age 7 Teacher | 4,453 | 2,287 | 0.177 [0.147, 0.206] SE 0.015 *p* < .001 *q* < .001 | 0.160 [0.113, 0.208] SE 0.024 *p* < .001 *q* < .001 | 0.182 [0.145, 0.217] SE 0.018 *p* < .001 *q* < .001 | 0.017 [−0.029, 0.061] SE 0.023 *p* = .468 *q* = .861 |
| Age 8 Parent | 4,794 | 2,397 | 0.150 [0.120, 0.179] SE 0.015 *p* < .001 *q* < .001 | 0.129 [0.087, 0.170] SE 0.021 *p* < .001 *q* < .001 | 0.157 [0.120, 0.195] SE 0.019 *p* < .001 *q* < .001 | 0.021 [−0.021, 0.064] SE 0.022 *p* = .325 *q* = .781 |
| Age 12 Parent | 4,532 | 2,266 | 0.144 [0.113, 0.175] SE 0.016 *p* < .001 *q* < .001 | 0.132 [0.091, 0.172] SE 0.021 *p* < .001 *q* < .001 | 0.148 [0.109, 0.186] SE 0.020 *p* < .001 *q* < .001 | 0.012 [−0.030, 0.055] SE 0.022 *p* = .579 *q* = .907 |
| Age 16 Parent | 4,015 | 2,008 | 0.133 [0.101, 0.166] SE 0.017 *p* < .001 *q* < .001 | 0.127 [0.084, 0.171] SE 0.023 *p* < .001 *q* < .001 | 0.135 [0.094, 0.175] SE 0.021 *p* < .001 *q* < .001 | 0.006 [−0.039, 0.050] SE 0.023 *p* = .804 *q* = .987 |
| Age 21 Parent | 4,226 | 2,117 | 0.117 [0.085, 0.149] SE 0.017 *p* < .001 *q* < .001 | 0.108 [0.060, 0.156] SE 0.024 *p* < .001 *q* < .001 | 0.120 [0.080, 0.160] SE 0.020 *p* < .001 *q* < .001 | 0.009 [−0.036, 0.056] SE 0.024 *p* = .705 *q* = .987 |
| Age 21 Self | 3,349 | 2,021 | 0.086 [0.052, 0.122] SE 0.018 *p* < .001 *q* < .001 | 0.080 [0.011, 0.154] SE 0.036 *p* = .027 *q* = .055 | 0.087 [0.049, 0.128] SE 0.020 *p* < .001 *q* < .001 | 0.006 [−0.059, 0.069] SE 0.033 *p* = .866 *q* = .987 |
| Age 26 Self | 2,981 | 1,946 | 0.085 [0.051, 0.121] SE 0.018 *p* < .001 *q* < .001 | 0.072 [−0.007, 0.152] SE 0.039 *p* = .070 *q* = .114 | 0.089 [0.050, 0.128] SE 0.020 *p* < .001 *q* < .001 | 0.014 [−0.057, 0.086] SE 0.036 *p* = .703 *q* = .987 |

**Notes:** Values are standardised regression coefficients (β), with 95% confidence intervals (CIs) in brackets, followed by standard errors (SE) and two-sided *p* values. N, number of offspring; Families, number of family clusters. Population, association of offspring PGS in the population model; Direct, within-family coefficient (child deviation from twin-pair mean PGS); Between, between-family coefficient (twin-pair mean PGS); Indirect, population minus within-family coefficient. SEs and percentile 95% CIs were obtained by bootstrapping with families as the resampling unit; *p* values were derived from z = β/SE using bootstrap SEs. False discovery rate (FDR)-adjusted *p* values (*q*; Benjamini–Hochberg) were computed separately for each estimate type across all PGS and outcomes in this table. Models adjust for sex, genotyping chip and the first ten genetic principal components.

**Supplementary Table S3. Twin-comparison analyses of ADHD traits** *(continued)*

#### ASD PGS · Grove et al. (2019)

| Outcome (age, reporter) | N (offspring) | Families (clusters) | Population (β, 95% CI, SE, *p*, *q*) | Direct (β, 95% CI, SE, *p*, *q*) | Between (β, 95% CI, SE, *p*, *q*) | Indirect (β, 95% CI, SE, *p*, *q*) |
| --- | --- | --- | --- | --- | --- | --- |
| Age 4 Parent | 5,180 | 2,610 | 0.027 [0.000, 0.055] SE 0.014 *p* = .054 *q* = .066 | −0.001 [−0.053, 0.055] SE 0.027 *p* = .979 *q* = .979 | 0.037 [0.004, 0.070] SE 0.017 *p* = .030 *q* = .037 | 0.028 [−0.020, 0.074] SE 0.024 *p* = .246 *q* = .775 |
| Age 7 Parent | 5,317 | 2,659 | 0.019 [−0.008, 0.046] SE 0.014 *p* = .175 *q* = .195 | 0.006 [−0.048, 0.060] SE 0.027 *p* = .814 *q* = .865 | 0.023 [−0.008, 0.055] SE 0.016 *p* = .151 *q* = .175 | 0.013 [−0.035, 0.058] SE 0.024 *p* = .598 *q* = .917 |
| Age 7 Teacher | 4,453 | 2,287 | 0.036 [0.006, 0.067] SE 0.015 *p* = .016 *q* = .022 | 0.024 [−0.024, 0.074] SE 0.025 *p* = .327 *q* = .445 | 0.040 [0.003, 0.078] SE 0.019 *p* = .035 *q* = .044 | 0.012 [−0.036, 0.059] SE 0.024 *p* = .615 *q* = .923 |
| Age 8 Parent | 4,794 | 2,397 | 0.056 [0.026, 0.087] SE 0.015 *p* < .001 *q* < .001 | 0.062 [0.018, 0.106] SE 0.022 *p* = .005 *q* = .011 | 0.054 [0.016, 0.093] SE 0.019 *p* = .004 *q* = .007 | −0.006 [−0.050, 0.036] SE 0.022 *p* = .784 *q* = .987 |
| Age 12 Parent | 4,532 | 2,266 | 0.053 [0.022, 0.086] SE 0.016 *p* < .001 *q* = .002 | 0.048 [0.005, 0.089] SE 0.022 *p* = .027 *q* = .055 | 0.055 [0.016, 0.097] SE 0.021 *p* = .009 *q* = .012 | 0.005 [−0.036, 0.050] SE 0.022 *p* = .809 *q* = .987 |
| Age 16 Parent | 4,015 | 2,008 | 0.052 [0.020, 0.085] SE 0.016 *p* = .001 *q* = .002 | 0.052 [0.007, 0.099] SE 0.024 *p* = .027 *q* = .055 | 0.052 [0.011, 0.094] SE 0.021 *p* = .013 *q* = .018 | 0.000 [−0.048, 0.046] SE 0.024 *p* = .987 *q* = .987 |
| Age 21 Parent | 4,226 | 2,117 | 0.052 [0.021, 0.084] SE 0.016 *p* = .001 *q* = .002 | 0.047 [−0.002, 0.095] SE 0.025 *p* = .058 *q* = .104 | 0.054 [0.016, 0.093] SE 0.020 *p* = .007 *q* = .010 | 0.005 [−0.039, 0.050] SE 0.023 *p* = .816 *q* = .987 |
| Age 21 Self | 3,349 | 2,021 | 0.049 [0.014, 0.083] SE 0.017 *p* = .004 *q* = .006 | −0.017 [−0.084, 0.051] SE 0.035 *p* = .626 *q* = .738 | 0.066 [0.027, 0.105] SE 0.020 *p* < .001 *q* = .002 | 0.066 [0.004, 0.128] SE 0.032 *p* = .038 *q* = .469 |
| Age 26 Self | 2,981 | 1,946 | 0.061 [0.026, 0.097] SE 0.018 *p* < .001 *q* = .001 | 0.029 [−0.051, 0.106] SE 0.040 *p* = .474 *q* = .598 | 0.067 [0.028, 0.108] SE 0.021 *p* = .001 *q* = .003 | 0.032 [−0.039, 0.105] SE 0.038 *p* = .399 *q* = .837 |

**Notes:** Values are standardised regression coefficients (β), with 95% confidence intervals (CIs) in brackets, followed by standard errors (SE) and two-sided *p* values. N, number of offspring; Families, number of family clusters. Population, association of offspring PGS in the population model; Direct, within-family coefficient (child deviation from twin-pair mean PGS); Between, between-family coefficient (twin-pair mean PGS); Indirect, population minus within-family coefficient. SEs and percentile 95% CIs were obtained by bootstrapping with families as the resampling unit; *p* values were derived from z = β/SE using bootstrap SEs. False discovery rate (FDR)-adjusted *p* values (*q*; Benjamini–Hochberg) were computed separately for each estimate type across all PGS and outcomes in this table. Models adjust for sex, genotyping chip and the first ten genetic principal components.

**Supplementary Table S3. Twin-comparison analyses of ADHD traits** *(continued)*

#### Depression PGS · Howard et al. (2019)

| Outcome (age, reporter) | N (offspring) | Families (clusters) | Population (β, 95% CI, SE, *p*, *q*) | Direct (β, 95% CI, SE, *p*, *q*) | Between (β, 95% CI, SE, *p*, *q*) | Indirect (β, 95% CI, SE, *p*, *q*) |
| --- | --- | --- | --- | --- | --- | --- |
| Age 4 Parent | 5,180 | 2,610 | 0.046 [0.018, 0.072] SE 0.014 *p* = .001 *q* = .002 | 0.037 [−0.017, 0.092] SE 0.028 *p* = .178 *q* = .262 | 0.048 [0.017, 0.078] SE 0.016 *p* = .003 *q* = .004 | 0.008 [−0.040, 0.055] SE 0.024 *p* = .732 *q* = .987 |
| Age 7 Parent | 5,317 | 2,659 | 0.054 [0.028, 0.081] SE 0.013 *p* < .001 *q* < .001 | 0.058 [0.005, 0.111] SE 0.027 *p* = .033 *q* = .063 | 0.053 [0.023, 0.084] SE 0.015 *p* < .001 *q* < .001 | −0.003 [−0.049, 0.042] SE 0.024 *p* = .891 *q* = .987 |
| Age 7 Teacher | 4,453 | 2,287 | 0.046 [0.016, 0.076] SE 0.016 *p* = .004 *q* = .006 | 0.008 [−0.041, 0.058] SE 0.025 *p* = .747 *q* = .826 | 0.057 [0.022, 0.094] SE 0.019 *p* = .003 *q* = .004 | 0.038 [−0.010, 0.084] SE 0.024 *p* = .119 *q* = .775 |
| Age 8 Parent | 4,794 | 2,397 | 0.065 [0.034, 0.096] SE 0.016 *p* < .001 *q* < .001 | 0.050 [0.005, 0.094] SE 0.023 *p* = .029 *q* = .057 | 0.069 [0.031, 0.108] SE 0.019 *p* < .001 *q* < .001 | 0.015 [−0.031, 0.059] SE 0.023 *p* = .514 *q* = .861 |
| Age 12 Parent | 4,532 | 2,266 | 0.087 [0.055, 0.118] SE 0.016 *p* < .001 *q* < .001 | 0.041 [−0.004, 0.087] SE 0.023 *p* = .073 *q* = .115 | 0.101 [0.063, 0.138] SE 0.020 *p* < .001 *q* < .001 | 0.045 [−0.002, 0.090] SE 0.023 *p* = .052 *q* = .469 |
| Age 16 Parent | 4,015 | 2,008 | 0.070 [0.039, 0.104] SE 0.017 *p* < .001 *q* < .001 | 0.043 [−0.004, 0.089] SE 0.024 *p* = .073 *q* = .115 | 0.079 [0.038, 0.120] SE 0.021 *p* < .001 *q* < .001 | 0.028 [−0.021, 0.076] SE 0.025 *p* = .256 *q* = .775 |
| Age 21 Parent | 4,226 | 2,117 | 0.072 [0.040, 0.105] SE 0.017 *p* < .001 *q* < .001 | 0.047 [−0.002, 0.097] SE 0.025 *p* = .066 *q* = .110 | 0.080 [0.040, 0.121] SE 0.020 *p* < .001 *q* < .001 | 0.025 [−0.023, 0.074] SE 0.025 *p* = .306 *q* = .776 |
| Age 21 Self | 3,349 | 2,021 | 0.085 [0.049, 0.120] SE 0.018 *p* < .001 *q* < .001 | 0.089 [0.016, 0.161] SE 0.037 *p* = .016 *q* = .036 | 0.084 [0.043, 0.125] SE 0.021 *p* < .001 *q* < .001 | −0.004 [−0.073, 0.065] SE 0.034 *p* = .906 *q* = .987 |
| Age 26 Self | 2,981 | 1,946 | 0.111 [0.073, 0.150] SE 0.020 *p* < .001 *q* < .001 | 0.107 [0.026, 0.189] SE 0.041 *p* = .009 *q* = .020 | 0.112 [0.070, 0.156] SE 0.022 *p* < .001 *q* < .001 | 0.004 [−0.073, 0.081] SE 0.039 *p* = .918 *q* = .987 |

**Notes:** Values are standardised regression coefficients (β), with 95% confidence intervals (CIs) in brackets, followed by standard errors (SE) and two-sided *p* values. N, number of offspring; Families, number of family clusters. Population, association of offspring PGS in the population model; Direct, within-family coefficient (child deviation from twin-pair mean PGS); Between, between-family coefficient (twin-pair mean PGS); Indirect, population minus within-family coefficient. SEs and percentile 95% CIs were obtained by bootstrapping with families as the resampling unit; *p* values were derived from z = β/SE using bootstrap SEs. False discovery rate (FDR)-adjusted *p* values (*q*; Benjamini–Hochberg) were computed separately for each estimate type across all PGS and outcomes in this table. Models adjust for sex, genotyping chip and the first ten genetic principal components.

**Supplementary Table S3. Twin-comparison analyses of ADHD traits** *(continued)*

#### Externalising PGS · Karlsson Linnér et al. (2021)

| Outcome (age, reporter) | N (offspring) | Families (clusters) | Population (β, 95% CI, SE, *p*, *q*) | Direct (β, 95% CI, SE, *p*, *q*) | Between (β, 95% CI, SE, *p*, *q*) | Indirect (β, 95% CI, SE, *p*, *q*) |
| --- | --- | --- | --- | --- | --- | --- |
| Age 4 Parent | 5,180 | 2,610 | 0.065 [0.038, 0.092] SE 0.014 *p* < .001 *q* < .001 | 0.091 [0.035, 0.143] SE 0.028 *p* = .001 *q* = .003 | 0.057 [0.026, 0.088] SE 0.016 *p* < .001 *q* < .001 | −0.026 [−0.072, 0.022] SE 0.025 *p* = .299 *q* = .776 |
| Age 7 Parent | 5,317 | 2,659 | 0.100 [0.072, 0.126] SE 0.014 *p* < .001 *q* < .001 | 0.143 [0.088, 0.198] SE 0.028 *p* < .001 *q* < .001 | 0.086 [0.055, 0.117] SE 0.016 *p* < .001 *q* < .001 | −0.043 [−0.092, 0.005] SE 0.025 *p* = .076 *q* = .612 |
| Age 7 Teacher | 4,453 | 2,287 | 0.122 [0.091, 0.153] SE 0.016 *p* < .001 *q* < .001 | 0.090 [0.038, 0.141] SE 0.026 *p* < .001 *q* = .002 | 0.131 [0.093, 0.168] SE 0.019 *p* < .001 *q* < .001 | 0.032 [−0.016, 0.082] SE 0.025 *p* = .200 *q* = .775 |
| Age 8 Parent | 4,794 | 2,397 | 0.119 [0.087, 0.150] SE 0.016 *p* < .001 *q* < .001 | 0.100 [0.054, 0.145] SE 0.023 *p* < .001 *q* < .001 | 0.125 [0.086, 0.162] SE 0.019 *p* < .001 *q* < .001 | 0.019 [−0.026, 0.062] SE 0.023 *p* = .413 *q* = .837 |
| Age 12 Parent | 4,532 | 2,266 | 0.115 [0.084, 0.146] SE 0.016 *p* < .001 *q* < .001 | 0.140 [0.096, 0.185] SE 0.022 *p* < .001 *q* < .001 | 0.107 [0.069, 0.147] SE 0.020 *p* < .001 *q* < .001 | −0.025 [−0.071, 0.020] SE 0.023 *p* = .280 *q* = .775 |
| Age 16 Parent | 4,015 | 2,008 | 0.130 [0.095, 0.165] SE 0.018 *p* < .001 *q* < .001 | 0.100 [0.054, 0.146] SE 0.024 *p* < .001 *q* < .001 | 0.139 [0.096, 0.182] SE 0.022 *p* < .001 *q* < .001 | 0.030 [−0.016, 0.078] SE 0.024 *p* = .212 *q* = .775 |
| Age 21 Parent | 4,226 | 2,117 | 0.122 [0.090, 0.154] SE 0.017 *p* < .001 *q* < .001 | 0.120 [0.076, 0.163] SE 0.022 *p* < .001 *q* < .001 | 0.123 [0.082, 0.163] SE 0.020 *p* < .001 *q* < .001 | 0.002 [−0.043, 0.046] SE 0.023 *p* = .927 *q* = .987 |
| Age 21 Self | 3,349 | 2,021 | 0.106 [0.070, 0.141] SE 0.018 *p* < .001 *q* < .001 | 0.151 [0.078, 0.224] SE 0.037 *p* < .001 *q* < .001 | 0.096 [0.055, 0.136] SE 0.021 *p* < .001 *q* < .001 | −0.045 [−0.114, 0.021] SE 0.035 *p* = .198 *q* = .775 |
| Age 26 Self | 2,981 | 1,946 | 0.092 [0.053, 0.130] SE 0.020 *p* < .001 *q* < .001 | 0.141 [0.055, 0.229] SE 0.044 *p* = .001 *q* = .003 | 0.082 [0.039, 0.124] SE 0.022 *p* < .001 *q* < .001 | −0.050 [−0.131, 0.030] SE 0.041 *p* = .224 *q* = .775 |

**Notes:** Values are standardised regression coefficients (β), with 95% confidence intervals (CIs) in brackets, followed by standard errors (SE) and two-sided *p* values. N, number of offspring; Families, number of family clusters. Population, association of offspring PGS in the population model; Direct, within-family coefficient (child deviation from twin-pair mean PGS); Between, between-family coefficient (twin-pair mean PGS); Indirect, population minus within-family coefficient. SEs and percentile 95% CIs were obtained by bootstrapping with families as the resampling unit; *p* values were derived from z = β/SE using bootstrap SEs. False discovery rate (FDR)-adjusted *p* values (*q*; Benjamini–Hochberg) were computed separately for each estimate type across all PGS and outcomes in this table. Models adjust for sex, genotyping chip and the first ten genetic principal components.

**Supplementary Table S3. Twin-comparison analyses of ADHD traits** *(continued)*

#### Anxiety PGS · Purves et al. (2020)

| Outcome (age, reporter) | N (offspring) | Families (clusters) | Population (β, 95% CI, SE, *p*, *q*) | Direct (β, 95% CI, SE, *p*, *q*) | Between (β, 95% CI, SE, *p*, *q*) | Indirect (β, 95% CI, SE, *p*, *q*) |
| --- | --- | --- | --- | --- | --- | --- |
| Age 4 Parent | 5,180 | 2,610 | −0.019 [−0.047, 0.008] SE 0.014 *p* = .175 *q* = .195 | −0.024 [−0.077, 0.027] SE 0.027 *p* = .364 *q* = .482 | −0.017 [−0.049, 0.014] SE 0.016 *p* = .288 *q* = .319 | 0.005 [−0.039, 0.051] SE 0.023 *p* = .811 *q* = .987 |
| Age 7 Parent | 5,317 | 2,659 | −0.034 [−0.060, −0.007] SE 0.014 *p* = .015 *q* = .021 | −0.020 [−0.071, 0.030] SE 0.026 *p* = .437 *q* = .562 | −0.039 [−0.069, −0.008] SE 0.016 *p* = .015 *q* = .020 | −0.014 [−0.056, 0.029] SE 0.022 *p* = .520 *q* = .861 |
| Age 7 Teacher | 4,453 | 2,287 | −0.005 [−0.034, 0.024] SE 0.015 *p* = .739 *q* = .771 | 0.011 [−0.035, 0.057] SE 0.024 *p* = .652 *q* = .758 | −0.010 [−0.045, 0.027] SE 0.019 *p* = .599 *q* = .607 | −0.015 [−0.060, 0.028] SE 0.022 *p* = .488 *q* = .861 |
| Age 8 Parent | 4,794 | 2,397 | −0.017 [−0.048, 0.014] SE 0.016 *p* = .288 *q* = .310 | −0.004 [−0.044, 0.038] SE 0.021 *p* = .847 *q* = .884 | −0.022 [−0.060, 0.018] SE 0.020 *p* = .271 *q* = .305 | −0.013 [−0.053, 0.026] SE 0.020 *p* = .526 *q* = .861 |
| Age 12 Parent | 4,532 | 2,266 | −0.037 [−0.068, −0.005] SE 0.016 *p* = .021 *q* = .027 | −0.021 [−0.063, 0.020] SE 0.021 *p* = .308 *q* = .427 | −0.042 [−0.082, 0.000] SE 0.021 *p* = .046 *q* = .056 | −0.015 [−0.057, 0.027] SE 0.022 *p* = .475 *q* = .861 |
| Age 16 Parent | 4,015 | 2,008 | −0.053 [−0.087, −0.020] SE 0.017 *p* = .002 *q* = .003 | −0.029 [−0.070, 0.013] SE 0.021 *p* = .172 *q* = .259 | −0.062 [−0.104, −0.019] SE 0.021 *p* = .003 *q* = .005 | −0.024 [−0.067, 0.018] SE 0.021 *p* = .262 *q* = .775 |
| Age 21 Parent | 4,226 | 2,117 | −0.022 [−0.054, 0.010] SE 0.016 *p* = .169 *q* = .195 | −0.015 [−0.062, 0.031] SE 0.024 *p* = .544 *q* = .664 | −0.025 [−0.066, 0.015] SE 0.021 *p* = .234 *q* = .267 | −0.007 [−0.054, 0.040] SE 0.024 *p* = .756 *q* = .987 |
| Age 21 Self | 3,349 | 2,021 | −0.048 [−0.081, −0.013] SE 0.017 *p* = .005 *q* = .007 | −0.022 [−0.088, 0.046] SE 0.034 *p* = .515 *q* = .639 | −0.054 [−0.094, −0.015] SE 0.020 *p* = .007 *q* = .010 | −0.026 [−0.087, 0.037] SE 0.031 *p* = .414 *q* = .837 |
| Age 26 Self | 2,981 | 1,946 | −0.073 [−0.110, −0.036] SE 0.019 *p* < .001 *q* < .001 | −0.076 [−0.148, −0.002] SE 0.037 *p* = .042 *q* = .077 | −0.073 [−0.115, −0.031] SE 0.021 *p* < .001 *q* < .001 | 0.002 [−0.068, 0.071] SE 0.035 *p* = .948 *q* = .987 |

**Notes:** Values are standardised regression coefficients (β), with 95% confidence intervals (CIs) in brackets, followed by standard errors (SE) and two-sided *p* values. N, number of offspring; Families, number of family clusters. Population, association of offspring PGS in the population model; Direct, within-family coefficient (child deviation from twin-pair mean PGS); Between, between-family coefficient (twin-pair mean PGS); Indirect, population minus within-family coefficient. SEs and percentile 95% CIs were obtained by bootstrapping with families as the resampling unit; *p* values were derived from z = β/SE using bootstrap SEs. False discovery rate (FDR)-adjusted *p* values (*q*; Benjamini–Hochberg) were computed separately for each estimate type across all PGS and outcomes in this table. Models adjust for sex, genotyping chip and the first ten genetic principal components.

**Supplementary Table S3. Twin-comparison analyses of ADHD traits** *(continued)*

#### Neuroticism PGS · Nagel et al. (2018)

| Outcome (age, reporter) | N (offspring) | Families (clusters) | Population (β, 95% CI, SE, *p*, *q*) | Direct (β, 95% CI, SE, *p*, *q*) | Between (β, 95% CI, SE, *p*, *q*) | Indirect (β, 95% CI, SE, *p*, *q*) |
| --- | --- | --- | --- | --- | --- | --- |
| Age 4 Parent | 5,180 | 2,610 | 0.031 [0.003, 0.057] SE 0.014 *p* = .027 *q* = .034 | −0.016 [−0.067, 0.037] SE 0.027 *p* = .556 *q* = .667 | 0.046 [0.015, 0.077] SE 0.016 *p* = .004 *q* = .006 | 0.046 [0.001, 0.090] SE 0.023 *p* = .045 *q* = .469 |
| Age 7 Parent | 5,317 | 2,659 | 0.028 [0.001, 0.054] SE 0.013 *p* = .031 *q* = .039 | −0.006 [−0.057, 0.046] SE 0.026 *p* = .817 *q* = .865 | 0.040 [0.009, 0.071] SE 0.016 *p* = .012 *q* = .017 | 0.034 [−0.012, 0.078] SE 0.023 *p* = .135 *q* = .775 |
| Age 7 Teacher | 4,453 | 2,287 | 0.000 [−0.031, 0.030] SE 0.015 *p* = 1.000 *q* = 1.000 | −0.031 [−0.080, 0.017] SE 0.025 *p* = .223 *q* = .321 | 0.010 [−0.028, 0.046] SE 0.018 *p* = .579 *q* = .595 | 0.030 [−0.015, 0.075] SE 0.023 *p* = .192 *q* = .775 |
| Age 8 Parent | 4,794 | 2,397 | 0.020 [−0.012, 0.048] SE 0.015 *p* = .182 *q* = .199 | −0.024 [−0.066, 0.017] SE 0.022 *p* = .260 *q* = .367 | 0.034 [−0.005, 0.071] SE 0.019 *p* = .074 *q* = .088 | 0.044 [0.001, 0.088] SE 0.022 *p* = .046 *q* = .469 |
| Age 12 Parent | 4,532 | 2,266 | 0.049 [0.018, 0.080] SE 0.016 *p* = .002 *q* = .003 | 0.008 [−0.034, 0.050] SE 0.021 *p* = .712 *q* = .801 | 0.063 [0.025, 0.101] SE 0.020 *p* = .002 *q* = .003 | 0.042 [0.000, 0.083] SE 0.021 *p* = .052 *q* = .469 |
| Age 16 Parent | 4,015 | 2,008 | 0.023 [−0.012, 0.056] SE 0.017 *p* = .176 *q* = .195 | 0.044 [−0.004, 0.091] SE 0.024 *p* = .066 *q* = .110 | 0.015 [−0.028, 0.058] SE 0.022 *p* = .495 *q* = .524 | −0.021 [−0.068, 0.027] SE 0.024 *p* = .376 *q* = .837 |
| Age 21 Parent | 4,226 | 2,117 | 0.026 [−0.006, 0.057] SE 0.016 *p* = .104 *q* = .125 | 0.007 [−0.040, 0.053] SE 0.024 *p* = .757 *q* = .826 | 0.032 [−0.007, 0.071] SE 0.020 *p* = .110 *q* = .129 | 0.018 [−0.026, 0.065] SE 0.023 *p* = .424 *q* = .837 |
| Age 21 Self | 3,349 | 2,021 | 0.035 [0.002, 0.067] SE 0.017 *p* = .040 *q* = .049 | 0.003 [−0.063, 0.070] SE 0.034 *p* = .920 *q* = .933 | 0.043 [0.006, 0.081] SE 0.019 *p* = .024 *q* = .031 | 0.031 [−0.029, 0.093] SE 0.031 *p* = .312 *q* = .776 |
| Age 26 Self | 2,981 | 1,946 | 0.048 [0.011, 0.087] SE 0.019 *p* = .012 *q* = .016 | 0.050 [−0.021, 0.121] SE 0.037 *p* = .173 *q* = .259 | 0.048 [0.005, 0.092] SE 0.022 *p* = .029 *q* = .037 | −0.002 [−0.069, 0.067] SE 0.035 *p* = .960 *q* = .987 |

**Notes:** Values are standardised regression coefficients (β), with 95% confidence intervals (CIs) in brackets, followed by standard errors (SE) and two-sided *p* values. N, number of offspring; Families, number of family clusters. Population, association of offspring PGS in the population model; Direct, within-family coefficient (child deviation from twin-pair mean PGS); Between, between-family coefficient (twin-pair mean PGS); Indirect, population minus within-family coefficient. SEs and percentile 95% CIs were obtained by bootstrapping with families as the resampling unit; *p* values were derived from z = β/SE using bootstrap SEs. False discovery rate (FDR)-adjusted *p* values (*q*; Benjamini–Hochberg) were computed separately for each estimate type across all PGS and outcomes in this table. Models adjust for sex, genotyping chip and the first ten genetic principal components.

**Supplementary Table S3. Twin-comparison analyses of ADHD traits** *(continued)*

#### Cognitive PGS · Demange et al. (2021)

| Outcome (age, reporter) | N (offspring) | Families (clusters) | Population (β, 95% CI, SE, *p*, *q*) | Direct (β, 95% CI, SE, *p*, *q*) | Between (β, 95% CI, SE, *p*, *q*) | Indirect (β, 95% CI, SE, *p*, *q*) |
| --- | --- | --- | --- | --- | --- | --- |
| Age 4 Parent | 5,180 | 2,610 | −0.070 [−0.096, −0.043] SE 0.014 *p* < .001 *q* < .001 | −0.110 [−0.162, −0.059] SE 0.026 *p* < .001 *q* < .001 | −0.058 [−0.089, −0.025] SE 0.016 *p* < .001 *q* < .001 | 0.040 [−0.005, 0.086] SE 0.023 *p* = .087 *q* = .626 |
| Age 7 Parent | 5,317 | 2,659 | −0.091 [−0.117, −0.064] SE 0.013 *p* < .001 *q* < .001 | −0.141 [−0.191, −0.088] SE 0.026 *p* < .001 *q* < .001 | −0.075 [−0.104, −0.044] SE 0.015 *p* < .001 *q* < .001 | 0.050 [0.005, 0.094] SE 0.023 *p* = .026 *q* = .469 |
| Age 7 Teacher | 4,453 | 2,287 | −0.114 [−0.144, −0.084] SE 0.016 *p* < .001 *q* < .001 | −0.089 [−0.137, −0.043] SE 0.024 *p* < .001 *q* < .001 | −0.122 [−0.159, −0.084] SE 0.019 *p* < .001 *q* < .001 | −0.025 [−0.071, 0.021] SE 0.023 *p* = .280 *q* = .775 |
| Age 8 Parent | 4,794 | 2,397 | −0.094 [−0.124, −0.062] SE 0.016 *p* < .001 *q* < .001 | −0.093 [−0.138, −0.048] SE 0.023 *p* < .001 *q* < .001 | −0.094 [−0.130, −0.055] SE 0.019 *p* < .001 *q* < .001 | −0.001 [−0.045, 0.044] SE 0.023 *p* = .970 *q* = .987 |
| Age 12 Parent | 4,532 | 2,266 | −0.089 [−0.121, −0.057] SE 0.016 *p* < .001 *q* < .001 | −0.079 [−0.123, −0.038] SE 0.022 *p* < .001 *q* < .001 | −0.092 [−0.131, −0.054] SE 0.020 *p* < .001 *q* < .001 | −0.010 [−0.053, 0.033] SE 0.022 *p* = .657 *q* = .966 |
| Age 16 Parent | 4,015 | 2,008 | −0.069 [−0.103, −0.035] SE 0.017 *p* < .001 *q* < .001 | −0.084 [−0.128, −0.039] SE 0.023 *p* < .001 *q* < .001 | −0.064 [−0.106, −0.022] SE 0.021 *p* = .002 *q* = .004 | 0.015 [−0.029, 0.059] SE 0.023 *p* = .524 *q* = .861 |
| Age 21 Parent | 4,226 | 2,117 | −0.064 [−0.096, −0.033] SE 0.016 *p* < .001 *q* < .001 | −0.079 [−0.124, −0.033] SE 0.023 *p* < .001 *q* = .002 | −0.059 [−0.100, −0.020] SE 0.020 *p* = .003 *q* = .005 | 0.015 [−0.031, 0.062] SE 0.024 *p* = .540 *q* = .864 |
| Age 21 Self | 3,349 | 2,021 | 0.014 [−0.023, 0.051] SE 0.019 *p* = .461 *q* = .488 | −0.014 [−0.083, 0.054] SE 0.035 *p* = .695 *q* = .795 | 0.021 [−0.019, 0.062] SE 0.021 *p* = .317 *q* = .346 | 0.028 [−0.034, 0.087] SE 0.031 *p* = .377 *q* = .837 |
| Age 26 Self | 2,981 | 1,946 | 0.006 [−0.033, 0.043] SE 0.019 *p* = .752 *q* = .774 | −0.036 [−0.117, 0.041] SE 0.040 *p* = .368 *q* = .482 | 0.014 [−0.029, 0.057] SE 0.022 *p* = .525 *q* = .547 | 0.041 [−0.032, 0.117] SE 0.038 *p* = .273 *q* = .775 |

**Notes:** Values are standardised regression coefficients (β), with 95% confidence intervals (CIs) in brackets, followed by standard errors (SE) and two-sided *p* values. N, number of offspring; Families, number of family clusters. Population, association of offspring PGS in the population model; Direct, within-family coefficient (child deviation from twin-pair mean PGS); Between, between-family coefficient (twin-pair mean PGS); Indirect, population minus within-family coefficient. SEs and percentile 95% CIs were obtained by bootstrapping with families as the resampling unit; *p* values were derived from z = β/SE using bootstrap SEs. False discovery rate (FDR)-adjusted *p* values (*q*; Benjamini–Hochberg) were computed separately for each estimate type across all PGS and outcomes in this table. Models adjust for sex, genotyping chip and the first ten genetic principal components.

**Supplementary Table S3. Twin-comparison analyses of ADHD traits** *(continued)*

#### Educational attainment PGS · Okbay et al. (2022)

| Outcome (age, reporter) | N (offspring) | Families (clusters) | Population (β, 95% CI, SE, *p*, *q*) | Direct (β, 95% CI, SE, *p*, *q*) | Between (β, 95% CI, SE, *p*, *q*) | Indirect (β, 95% CI, SE, *p*, *q*) |
| --- | --- | --- | --- | --- | --- | --- |
| Age 4 Parent | 5,180 | 2,610 | −0.135 [−0.162, −0.108] SE 0.014 *p* < .001 *q* < .001 | −0.130 [−0.188, −0.073] SE 0.030 *p* < .001 *q* < .001 | −0.136 [−0.167, −0.106] SE 0.015 *p* < .001 *q* < .001 | −0.005 [−0.054, 0.046] SE 0.026 *p* = .858 *q* = .987 |
| Age 7 Parent | 5,317 | 2,659 | −0.139 [−0.166, −0.112] SE 0.014 *p* < .001 *q* < .001 | −0.142 [−0.201, −0.084] SE 0.030 *p* < .001 *q* < .001 | −0.138 [−0.167, −0.109] SE 0.015 *p* < .001 *q* < .001 | 0.003 [−0.048, 0.054] SE 0.026 *p* = .895 *q* = .987 |
| Age 7 Teacher | 4,453 | 2,287 | −0.143 [−0.173, −0.113] SE 0.015 *p* < .001 *q* < .001 | −0.112 [−0.165, −0.060] SE 0.027 *p* < .001 *q* < .001 | −0.151 [−0.185, −0.116] SE 0.018 *p* < .001 *q* < .001 | −0.031 [−0.081, 0.019] SE 0.026 *p* = .223 *q* = .775 |
| Age 8 Parent | 4,794 | 2,397 | −0.117 [−0.147, −0.085] SE 0.016 *p* < .001 *q* < .001 | −0.085 [−0.132, −0.038] SE 0.024 *p* < .001 *q* < .001 | −0.125 [−0.160, −0.087] SE 0.019 *p* < .001 *q* < .001 | −0.032 [−0.079, 0.015] SE 0.024 *p* = .192 *q* = .775 |
| Age 12 Parent | 4,532 | 2,266 | −0.109 [−0.142, −0.077] SE 0.016 *p* < .001 *q* < .001 | −0.110 [−0.160, −0.061] SE 0.025 *p* < .001 *q* < .001 | −0.109 [−0.145, −0.072] SE 0.019 *p* < .001 *q* < .001 | 0.001 [−0.043, 0.049] SE 0.024 *p* = .961 *q* = .987 |
| Age 16 Parent | 4,015 | 2,008 | −0.118 [−0.152, −0.083] SE 0.017 *p* < .001 *q* < .001 | −0.098 [−0.145, −0.048] SE 0.025 *p* < .001 *q* < .001 | −0.123 [−0.164, −0.083] SE 0.021 *p* < .001 *q* < .001 | −0.020 [−0.071, 0.031] SE 0.026 *p* = .430 *q* = .837 |
| Age 21 Parent | 4,226 | 2,117 | −0.064 [−0.097, −0.032] SE 0.016 *p* < .001 *q* < .001 | −0.068 [−0.121, −0.016] SE 0.027 *p* = .012 *q* = .026 | −0.063 [−0.102, −0.024] SE 0.020 *p* = .002 *q* = .003 | 0.004 [−0.048, 0.057] SE 0.027 *p* = .875 *q* = .987 |
| Age 21 Self | 3,349 | 2,021 | −0.026 [−0.063, 0.008] SE 0.019 *p* = .171 *q* = .195 | −0.076 [−0.158, 0.004] SE 0.041 *p* = .063 *q* = .110 | −0.016 [−0.056, 0.023] SE 0.020 *p* = .424 *q* = .455 | 0.050 [−0.023, 0.123] SE 0.038 *p* = .184 *q* = .775 |
| Age 26 Self | 2,981 | 1,946 | −0.004 [−0.040, 0.035] SE 0.019 *p* = .833 *q* = .845 | −0.005 [−0.093, 0.081] SE 0.044 *p* = .912 *q* = .933 | −0.003 [−0.045, 0.038] SE 0.021 *p* = .886 *q* = .886 | 0.001 [−0.077, 0.085] SE 0.041 *p* = .977 *q* = .987 |

**Notes:** Values are standardised regression coefficients (β), with 95% confidence intervals (CIs) in brackets, followed by standard errors (SE) and two-sided *p* values. N, number of offspring; Families, number of family clusters. Population, association of offspring PGS in the population model; Direct, within-family coefficient (child deviation from twin-pair mean PGS); Between, between-family coefficient (twin-pair mean PGS); Indirect, population minus within-family coefficient. SEs and percentile 95% CIs were obtained by bootstrapping with families as the resampling unit; *p* values were derived from z = β/SE using bootstrap SEs. False discovery rate (FDR)-adjusted *p* values (*q*; Benjamini–Hochberg) were computed separately for each estimate type across all PGS and outcomes in this table. Models adjust for sex, genotyping chip and the first ten genetic principal components.

### Supplementary Table S4. Twin-comparison analyses of ASD traits

#### ADHD PGS · Demontis et al. (2023)

| Outcome (age, reporter) | N (offspring) | Families (clusters) | Population (β, 95% CI, SE, *p*, *q*) | Direct (β, 95% CI, SE, *p*, *q*) | Between (β, 95% CI, SE, *p*, *q*) | Indirect (β, 95% CI, SE, *p*, *q*) |
| --- | --- | --- | --- | --- | --- | --- |
| Age 8 Parent | 4,802 | 2,402 | 0.081 [0.053, 0.110] SE 0.015 *p* < .001 *q* < .001 | 0.016 [−0.031, 0.061] SE 0.023 *p* = .495 *q* = .793 | 0.101 [0.067, 0.138] SE 0.019 *p* < .001 *q* < .001 | 0.065 [0.020, 0.112] SE 0.023 *p* = .005 *q* = .043 |
| Age 9 Parent | 2,474 | 1,237 | 0.076 [0.032, 0.121] SE 0.023 *p* < .001 *q* = .002 | −0.005 [−0.059, 0.048] SE 0.027 *p* = .842 *q* = .951 | 0.102 [0.046, 0.159] SE 0.029 *p* < .001 *q* = .002 | 0.082 [0.021, 0.141] SE 0.031 *p* = .008 *q* = .054 |
| Age 9 Teacher | 2,084 | 1,108 | 0.076 [0.031, 0.121] SE 0.023 *p* < .001 *q* = .002 | 0.079 [0.011, 0.147] SE 0.034 *p* = .021 *q* = .129 | 0.076 [0.020, 0.131] SE 0.029 *p* = .009 *q* = .019 | −0.002 [−0.069, 0.066] SE 0.034 *p* = .947 *q* = .962 |
| Age 9 Self | 2,373 | 1,218 | 0.048 [0.005, 0.090] SE 0.022 *p* = .029 *q* = .049 | 0.011 [−0.060, 0.077] SE 0.035 *p* = .758 *q* = .941 | 0.059 [0.007, 0.109] SE 0.026 *p* = .023 *q* = .045 | 0.037 [−0.028, 0.101] SE 0.033 *p* = .260 *q* = .467 |
| Age 12 Parent | 4,624 | 2,312 | 0.081 [0.050, 0.113] SE 0.016 *p* < .001 *q* < .001 | 0.031 [−0.014, 0.075] SE 0.023 *p* = .170 *q* = .434 | 0.097 [0.058, 0.135] SE 0.020 *p* < .001 *q* < .001 | 0.050 [0.007, 0.093] SE 0.022 *p* = .025 *q* = .119 |
| Age 12 Teacher | 3,699 | 2,064 | 0.061 [0.027, 0.098] SE 0.018 *p* < .001 *q* = .002 | 0.020 [−0.039, 0.081] SE 0.030 *p* = .513 *q* = .793 | 0.072 [0.034, 0.113] SE 0.021 *p* < .001 *q* = .002 | 0.042 [−0.013, 0.096] SE 0.028 *p* = .133 *q* = .367 |
| Age 16 Parent | 4,013 | 2,008 | 0.048 [0.013, 0.081] SE 0.018 *p* = .008 *q* = .014 | −0.001 [−0.046, 0.042] SE 0.022 *p* = .954 *q* = .981 | 0.064 [0.021, 0.107] SE 0.022 *p* = .004 *q* = .009 | 0.049 [0.002, 0.094] SE 0.023 *p* = .037 *q* = .155 |
| Age 16 Self | 4,004 | 2,018 | 0.003 [−0.030, 0.034] SE 0.016 *p* = .851 *q* = .888 | −0.014 [−0.069, 0.042] SE 0.028 *p* = .632 *q* = .887 | 0.008 [−0.031, 0.046] SE 0.019 *p* = .674 *q* = .713 | 0.016 [−0.036, 0.065] SE 0.026 *p* = .527 *q* = .731 |
| Age 26 Self | 2,981 | 1,944 | 0.088 [0.051, 0.124] SE 0.019 *p* < .001 *q* < .001 | 0.035 [−0.043, 0.114] SE 0.040 *p* = .376 *q* = .676 | 0.100 [0.059, 0.140] SE 0.021 *p* < .001 *q* < .001 | 0.053 [−0.018, 0.124] SE 0.037 *p* = .150 *q* = .376 |

**Notes:** Values are standardised regression coefficients (β), with 95% confidence intervals (CIs) in brackets, followed by standard errors (SE) and two-sided *p* values. N, number of offspring; Families, number of family clusters. Population, association of offspring PGS in the population model; Direct, within-family coefficient (child deviation from twin-pair mean PGS); Between, between-family coefficient (twin-pair mean PGS); Indirect, population minus within-family coefficient. SEs and percentile 95% CIs were obtained by bootstrapping with families as the resampling unit; *p* values were derived from z = β/SE using bootstrap SEs. False discovery rate (FDR)-adjusted *p* values (*q*; Benjamini–Hochberg) were computed separately for each estimate type across all PGS and outcomes in this table. Models adjust for sex, genotyping chip and the first ten genetic principal components.

**Supplementary Table S4. Twin-comparison analyses of ASD traits** *(continued)*

#### ASD PGS · Grove et al. (2019)

| Outcome (age, reporter) | N (offspring) | Families (clusters) | Population (β, 95% CI, SE, *p*, *q*) | Direct (β, 95% CI, SE, *p*, *q*) | Between (β, 95% CI, SE, *p*, *q*) | Indirect (β, 95% CI, SE, *p*, *q*) |
| --- | --- | --- | --- | --- | --- | --- |
| Age 8 Parent | 4,802 | 2,402 | 0.060 [0.029, 0.090] SE 0.015 *p* < .001 *q* < .001 | 0.093 [0.048, 0.138] SE 0.023 *p* < .001 *q* = .001 | 0.048 [0.012, 0.086] SE 0.019 *p* = .012 *q* = .024 | −0.033 [−0.076, 0.009] SE 0.022 *p* = .129 *q* = .367 |
| Age 9 Parent | 2,474 | 1,237 | 0.036 [−0.006, 0.079] SE 0.022 *p* = .102 *q* = .150 | 0.068 [0.011, 0.126] SE 0.029 *p* = .020 *q* = .129 | 0.026 [−0.027, 0.080] SE 0.028 *p* = .353 *q* = .418 | −0.032 [−0.090, 0.029] SE 0.031 *p* = .303 *q* = .519 |
| Age 9 Teacher | 2,084 | 1,108 | 0.030 [−0.014, 0.073] SE 0.022 *p* = .173 *q* = .230 | 0.046 [−0.024, 0.113] SE 0.035 *p* = .189 *q* = .454 | 0.025 [−0.029, 0.078] SE 0.027 *p* = .354 *q* = .418 | −0.015 [−0.080, 0.051] SE 0.033 *p* = .643 *q* = .857 |
| Age 9 Self | 2,373 | 1,218 | 0.058 [0.014, 0.101] SE 0.022 *p* = .008 *q* = .015 | 0.062 [−0.005, 0.132] SE 0.035 *p* = .081 *q* = .265 | 0.057 [0.005, 0.108] SE 0.026 *p* = .028 *q* = .054 | −0.004 [−0.069, 0.059] SE 0.033 *p* = .911 *q* = .951 |
| Age 12 Parent | 4,624 | 2,312 | 0.075 [0.045, 0.106] SE 0.016 *p* < .001 *q* < .001 | 0.101 [0.054, 0.145] SE 0.023 *p* < .001 *q* < .001 | 0.066 [0.029, 0.104] SE 0.019 *p* < .001 *q* = .002 | −0.025 [−0.068, 0.018] SE 0.022 *p* = .245 *q* = .465 |
| Age 12 Teacher | 3,699 | 2,064 | 0.051 [0.017, 0.086] SE 0.018 *p* = .005 *q* = .010 | 0.072 [0.015, 0.131] SE 0.030 *p* = .015 *q* = .123 | 0.045 [0.004, 0.086] SE 0.021 *p* = .032 *q* = .059 | −0.021 [−0.077, 0.034] SE 0.028 *p* = .466 *q* = .697 |
| Age 16 Parent | 4,013 | 2,008 | 0.053 [0.020, 0.087] SE 0.017 *p* = .002 *q* = .004 | 0.048 [0.005, 0.094] SE 0.023 *p* = .033 *q* = .172 | 0.054 [0.013, 0.098] SE 0.022 *p* = .014 *q* = .028 | 0.004 [−0.041, 0.050] SE 0.023 *p* = .849 *q* = .929 |
| Age 16 Self | 4,004 | 2,018 | 0.059 [0.028, 0.091] SE 0.016 *p* < .001 *q* < .001 | 0.133 [0.079, 0.187] SE 0.027 *p* < .001 *q* < .001 | 0.033 [−0.005, 0.071] SE 0.019 *p* = .082 *q* = .132 | −0.074 [−0.123, −0.027] SE 0.025 *p* = .003 *q* = .028 |
| Age 26 Self | 2,981 | 1,944 | 0.061 [0.025, 0.097] SE 0.018 *p* < .001 *q* = .002 | 0.067 [−0.006, 0.138] SE 0.037 *p* = .072 *q* = .260 | 0.059 [0.020, 0.100] SE 0.021 *p* = .005 *q* = .011 | −0.007 [−0.075, 0.063] SE 0.035 *p* = .851 *q* = .929 |

**Notes:** Values are standardised regression coefficients (β), with 95% confidence intervals (CIs) in brackets, followed by standard errors (SE) and two-sided *p* values. N, number of offspring; Families, number of family clusters. Population, association of offspring PGS in the population model; Direct, within-family coefficient (child deviation from twin-pair mean PGS); Between, between-family coefficient (twin-pair mean PGS); Indirect, population minus within-family coefficient. SEs and percentile 95% CIs were obtained by bootstrapping with families as the resampling unit; *p* values were derived from z = β/SE using bootstrap SEs. False discovery rate (FDR)-adjusted *p* values (*q*; Benjamini–Hochberg) were computed separately for each estimate type across all PGS and outcomes in this table. Models adjust for sex, genotyping chip and the first ten genetic principal components.

**Supplementary Table S4. Twin-comparison analyses of ASD traits** *(continued)*

#### Depression PGS · Howard et al. (2019)

| Outcome (age, reporter) | N (offspring) | Families (clusters) | Population (β, 95% CI, SE, *p*, *q*) | Direct (β, 95% CI, SE, *p*, *q*) | Between (β, 95% CI, SE, *p*, *q*) | Indirect (β, 95% CI, SE, *p*, *q*) |
| --- | --- | --- | --- | --- | --- | --- |
| Age 8 Parent | 4,802 | 2,402 | 0.060 [0.030, 0.089] SE 0.015 *p* < .001 *q* < .001 | 0.043 [−0.003, 0.088] SE 0.023 *p* = .064 *q* = .248 | 0.065 [0.029, 0.101] SE 0.019 *p* < .001 *q* = .002 | 0.017 [−0.028, 0.060] SE 0.023 *p* = .466 *q* = .697 |
| Age 9 Parent | 2,474 | 1,237 | 0.062 [0.018, 0.108] SE 0.023 *p* = .007 *q* = .013 | 0.070 [0.011, 0.129] SE 0.030 *p* = .019 *q* = .129 | 0.060 [0.004, 0.117] SE 0.029 *p* = .039 *q* = .069 | −0.007 [−0.070, 0.055] SE 0.032 *p* = .817 *q* = .929 |
| Age 9 Teacher | 2,084 | 1,108 | 0.001 [−0.043, 0.047] SE 0.023 *p* = .965 *q* = .965 | 0.014 [−0.056, 0.087] SE 0.036 *p* = .703 *q* = .938 | −0.003 [−0.056, 0.053] SE 0.028 *p* = .915 *q* = .941 | −0.013 [−0.084, 0.057] SE 0.036 *p* = .718 *q* = .892 |
| Age 9 Self | 2,373 | 1,218 | 0.073 [0.030, 0.115] SE 0.021 *p* < .001 *q* = .002 | 0.065 [−0.005, 0.133] SE 0.035 *p* = .062 *q* = .248 | 0.075 [0.023, 0.126] SE 0.026 *p* = .004 *q* = .009 | 0.008 [−0.060, 0.073] SE 0.034 *p* = .812 *q* = .929 |
| Age 12 Parent | 4,624 | 2,312 | 0.055 [0.025, 0.086] SE 0.015 *p* < .001 *q* < .001 | 0.047 [0.000, 0.093] SE 0.024 *p* = .048 *q* = .217 | 0.058 [0.021, 0.096] SE 0.019 *p* = .002 *q* = .007 | 0.008 [−0.039, 0.055] SE 0.024 *p* = .731 *q* = .892 |
| Age 12 Teacher | 3,699 | 2,064 | 0.019 [−0.014, 0.052] SE 0.017 *p* = .264 *q* = .333 | 0.060 [0.003, 0.116] SE 0.029 *p* = .042 *q* = .203 | 0.008 [−0.032, 0.047] SE 0.020 *p* = .689 *q* = .719 | −0.040 [−0.095, 0.016] SE 0.028 *p* = .152 *q* = .376 |
| Age 16 Parent | 4,013 | 2,008 | 0.040 [0.007, 0.072] SE 0.017 *p* = .019 *q* = .033 | 0.038 [−0.006, 0.082] SE 0.022 *p* = .086 *q* = .270 | 0.040 [−0.002, 0.082] SE 0.021 *p* = .057 *q* = .097 | 0.002 [−0.046, 0.049] SE 0.024 *p* = .949 *q* = .962 |
| Age 16 Self | 4,004 | 2,018 | 0.027 [−0.004, 0.060] SE 0.016 *p* = .092 *q* = .140 | 0.019 [−0.035, 0.074] SE 0.029 *p* = .503 *q* = .793 | 0.030 [−0.007, 0.069] SE 0.020 *p* = .134 *q* = .192 | 0.008 [−0.044, 0.060] SE 0.027 *p* = .762 *q* = .915 |
| Age 26 Self | 2,981 | 1,944 | 0.126 [0.087, 0.165] SE 0.020 *p* < .001 *q* < .001 | 0.057 [−0.017, 0.134] SE 0.039 *p* = .141 *q* = .406 | 0.139 [0.096, 0.182] SE 0.022 *p* < .001 *q* < .001 | 0.069 [−0.003, 0.142] SE 0.037 *p* = .063 *q* = .217 |

**Notes:** Values are standardised regression coefficients (β), with 95% confidence intervals (CIs) in brackets, followed by standard errors (SE) and two-sided *p* values. N, number of offspring; Families, number of family clusters. Population, association of offspring PGS in the population model; Direct, within-family coefficient (child deviation from twin-pair mean PGS); Between, between-family coefficient (twin-pair mean PGS); Indirect, population minus within-family coefficient. SEs and percentile 95% CIs were obtained by bootstrapping with families as the resampling unit; *p* values were derived from z = β/SE using bootstrap SEs. False discovery rate (FDR)-adjusted *p* values (*q*; Benjamini–Hochberg) were computed separately for each estimate type across all PGS and outcomes in this table. Models adjust for sex, genotyping chip and the first ten genetic principal components.

**Supplementary Table S4. Twin-comparison analyses of ASD traits** *(continued)*

#### Externalising PGS · Karlsson Linnér et al. (2021)

| Outcome (age, reporter) | N (offspring) | Families (clusters) | Population (β, 95% CI, SE, *p*, *q*) | Direct (β, 95% CI, SE, *p*, *q*) | Between (β, 95% CI, SE, *p*, *q*) | Indirect (β, 95% CI, SE, *p*, *q*) |
| --- | --- | --- | --- | --- | --- | --- |
| Age 8 Parent | 4,802 | 2,402 | 0.044 [0.014, 0.075] SE 0.016 *p* = .006 *q* = .012 | 0.003 [−0.041, 0.046] SE 0.023 *p* = .888 *q* = .969 | 0.057 [0.020, 0.095] SE 0.019 *p* = .003 *q* = .007 | 0.041 [−0.003, 0.088] SE 0.023 *p* = .075 *q* = .236 |
| Age 9 Parent | 2,474 | 1,237 | 0.060 [0.015, 0.103] SE 0.022 *p* = .006 *q* = .012 | −0.007 [−0.064, 0.050] SE 0.029 *p* = .819 *q* = .951 | 0.080 [0.025, 0.134] SE 0.028 *p* = .004 *q* = .010 | 0.066 [0.007, 0.127] SE 0.030 *p* = .029 *q* = .130 |
| Age 9 Teacher | 2,084 | 1,108 | 0.043 [−0.003, 0.091] SE 0.024 *p* = .073 *q* = .115 | 0.065 [−0.004, 0.136] SE 0.036 *p* = .065 *q* = .248 | 0.037 [−0.019, 0.093] SE 0.029 *p* = .202 *q* = .274 | −0.022 [−0.091, 0.046] SE 0.035 *p* = .527 *q* = .731 |
| Age 9 Self | 2,373 | 1,218 | 0.063 [0.019, 0.104] SE 0.022 *p* = .004 *q* = .009 | −0.016 [−0.090, 0.058] SE 0.037 *p* = .667 *q* = .906 | 0.087 [0.036, 0.135] SE 0.026 *p* < .001 *q* = .003 | 0.079 [0.012, 0.148] SE 0.034 *p* = .022 *q* = .115 |
| Age 12 Parent | 4,624 | 2,312 | 0.055 [0.022, 0.086] SE 0.016 *p* < .001 *q* = .002 | 0.023 [−0.020, 0.070] SE 0.023 *p* = .311 *q* = .639 | 0.064 [0.025, 0.102] SE 0.020 *p* = .001 *q* = .004 | 0.032 [−0.013, 0.078] SE 0.023 *p* = .172 *q* = .376 |
| Age 12 Teacher | 3,699 | 2,064 | 0.012 [−0.023, 0.049] SE 0.018 *p* = .505 *q* = .559 | −0.006 [−0.065, 0.054] SE 0.030 *p* = .845 *q* = .951 | 0.017 [−0.023, 0.059] SE 0.021 *p* = .418 *q* = .470 | 0.018 [−0.038, 0.076] SE 0.029 *p* = .528 *q* = .731 |
| Age 16 Parent | 4,013 | 2,008 | −0.036 [−0.069, −0.002] SE 0.017 *p* = .034 *q* = .056 | −0.091 [−0.134, −0.049] SE 0.022 *p* < .001 *q* < .001 | −0.019 [−0.061, 0.023] SE 0.021 *p* = .366 *q* = .425 | 0.056 [0.008, 0.101] SE 0.024 *p* = .019 *q* = .105 |
| Age 16 Self | 4,004 | 2,018 | −0.047 [−0.079, −0.013] SE 0.017 *p* = .006 *q* = .012 | −0.010 [−0.069, 0.046] SE 0.029 *p* = .744 *q* = .940 | −0.058 [−0.096, −0.018] SE 0.020 *p* = .004 *q* = .009 | −0.037 [−0.089, 0.015] SE 0.027 *p* = .163 *q* = .376 |
| Age 26 Self | 2,981 | 1,944 | 0.068 [0.031, 0.106] SE 0.019 *p* < .001 *q* = .001 | 0.076 [−0.010, 0.160] SE 0.044 *p* = .079 *q* = .265 | 0.067 [0.026, 0.109] SE 0.021 *p* = .001 *q* = .004 | −0.008 [−0.085, 0.071] SE 0.040 *p* = .837 *q* = .929 |

**Notes:** Values are standardised regression coefficients (β), with 95% confidence intervals (CIs) in brackets, followed by standard errors (SE) and two-sided *p* values. N, number of offspring; Families, number of family clusters. Population, association of offspring PGS in the population model; Direct, within-family coefficient (child deviation from twin-pair mean PGS); Between, between-family coefficient (twin-pair mean PGS); Indirect, population minus within-family coefficient. SEs and percentile 95% CIs were obtained by bootstrapping with families as the resampling unit; *p* values were derived from z = β/SE using bootstrap SEs. False discovery rate (FDR)-adjusted *p* values (*q*; Benjamini–Hochberg) were computed separately for each estimate type across all PGS and outcomes in this table. Models adjust for sex, genotyping chip and the first ten genetic principal components.

**Supplementary Table S4. Twin-comparison analyses of ASD traits** *(continued)*

#### Anxiety PGS · Purves et al. (2020)

| Outcome (age, reporter) | N (offspring) | Families (clusters) | Population (β, 95% CI, SE, *p*, *q*) | Direct (β, 95% CI, SE, *p*, *q*) | Between (β, 95% CI, SE, *p*, *q*) | Indirect (β, 95% CI, SE, *p*, *q*) |
| --- | --- | --- | --- | --- | --- | --- |
| Age 8 Parent | 4,802 | 2,402 | −0.024 [−0.054, 0.007] SE 0.015 *p* = .110 *q* = .158 | −0.032 [−0.076, 0.013] SE 0.023 *p* = .160 *q* = .427 | −0.021 [−0.058, 0.016] SE 0.019 *p* = .269 *q* = .343 | 0.008 [−0.033, 0.049] SE 0.021 *p* = .711 *q* = .892 |
| Age 9 Parent | 2,474 | 1,237 | −0.032 [−0.076, 0.011] SE 0.022 *p* = .146 *q* = .202 | 0.002 [−0.049, 0.053] SE 0.026 *p* = .939 *q* = .981 | −0.044 [−0.103, 0.011] SE 0.029 *p* = .129 *q* = .190 | −0.034 [−0.090, 0.022] SE 0.029 *p* = .238 *q* = .465 |
| Age 9 Teacher | 2,084 | 1,108 | −0.021 [−0.067, 0.026] SE 0.024 *p* = .382 *q* = .450 | −0.029 [−0.093, 0.036] SE 0.034 *p* = .395 *q* = .694 | −0.019 [−0.076, 0.040] SE 0.029 *p* = .512 *q* = .568 | 0.007 [−0.056, 0.071] SE 0.032 *p* = .816 *q* = .929 |
| Age 9 Self | 2,373 | 1,218 | 0.003 [−0.042, 0.049] SE 0.023 *p* = .896 *q* = .909 | 0.007 [−0.063, 0.079] SE 0.036 *p* = .841 *q* = .951 | 0.001 [−0.053, 0.056] SE 0.027 *p* = .970 *q* = .970 | −0.005 [−0.069, 0.059] SE 0.033 *p* = .887 *q* = .951 |
| Age 12 Parent | 4,624 | 2,312 | −0.024 [−0.056, 0.007] SE 0.016 *p* = .134 *q* = .189 | 0.000 [−0.044, 0.043] SE 0.022 *p* = .987 *q* = .997 | −0.033 [−0.073, 0.006] SE 0.020 *p* = .099 *q* = .155 | −0.024 [−0.066, 0.018] SE 0.021 *p* = .270 *q* = .474 |
| Age 12 Teacher | 3,699 | 2,064 | −0.019 [−0.053, 0.016] SE 0.018 *p* = .291 *q* = .361 | 0.000 [−0.055, 0.057] SE 0.029 *p* = .997 *q* = .997 | −0.026 [−0.067, 0.016] SE 0.021 *p* = .216 *q* = .288 | −0.020 [−0.074, 0.033] SE 0.027 *p* = .475 *q* = .697 |
| Age 16 Parent | 4,013 | 2,008 | −0.019 [−0.053, 0.014] SE 0.017 *p* = .264 *q* = .333 | 0.013 [−0.026, 0.051] SE 0.020 *p* = .518 *q* = .793 | −0.030 [−0.074, 0.012] SE 0.022 *p* = .173 *q* = .239 | −0.031 [−0.072, 0.009] SE 0.021 *p* = .130 *q* = .367 |
| Age 16 Self | 4,004 | 2,018 | −0.015 [−0.047, 0.018] SE 0.016 *p* = .349 *q* = .418 | 0.006 [−0.047, 0.060] SE 0.027 *p* = .836 *q* = .951 | −0.022 [−0.061, 0.016] SE 0.020 *p* = .271 *q* = .343 | −0.020 [−0.068, 0.028] SE 0.024 *p* = .408 *q* = .653 |
| Age 26 Self | 2,981 | 1,944 | −0.072 [−0.110, −0.034] SE 0.019 *p* < .001 *q* < .001 | −0.023 [−0.100, 0.054] SE 0.039 *p* = .562 *q* = .825 | −0.082 [−0.125, −0.040] SE 0.022 *p* < .001 *q* < .001 | −0.049 [−0.121, 0.024] SE 0.037 *p* = .188 *q* = .397 |

**Notes:** Values are standardised regression coefficients (β), with 95% confidence intervals (CIs) in brackets, followed by standard errors (SE) and two-sided *p* values. N, number of offspring; Families, number of family clusters. Population, association of offspring PGS in the population model; Direct, within-family coefficient (child deviation from twin-pair mean PGS); Between, between-family coefficient (twin-pair mean PGS); Indirect, population minus within-family coefficient. SEs and percentile 95% CIs were obtained by bootstrapping with families as the resampling unit; *p* values were derived from z = β/SE using bootstrap SEs. False discovery rate (FDR)-adjusted *p* values (*q*; Benjamini–Hochberg) were computed separately for each estimate type across all PGS and outcomes in this table. Models adjust for sex, genotyping chip and the first ten genetic principal components.

**Supplementary Table S4. Twin-comparison analyses of ASD traits** *(continued)*

#### Neuroticism PGS · Nagel et al. (2018)

| Outcome (age, reporter) | N (offspring) | Families (clusters) | Population (β, 95% CI, SE, *p*, *q*) | Direct (β, 95% CI, SE, *p*, *q*) | Between (β, 95% CI, SE, *p*, *q*) | Indirect (β, 95% CI, SE, *p*, *q*) |
| --- | --- | --- | --- | --- | --- | --- |
| Age 8 Parent | 4,802 | 2,402 | 0.046 [0.016, 0.074] SE 0.015 *p* = .002 *q* = .005 | 0.021 [−0.021, 0.066] SE 0.022 *p* = .337 *q* = .639 | 0.054 [0.018, 0.089] SE 0.018 *p* = .003 *q* = .007 | 0.024 [−0.018, 0.065] SE 0.021 *p* = .252 *q* = .466 |
| Age 9 Parent | 2,474 | 1,237 | 0.064 [0.018, 0.110] SE 0.023 *p* = .005 *q* = .011 | 0.009 [−0.041, 0.060] SE 0.026 *p* = .740 *q* = .940 | 0.083 [0.024, 0.140] SE 0.030 *p* = .006 *q* = .012 | 0.056 [−0.002, 0.112] SE 0.030 *p* = .060 *q* = .216 |
| Age 9 Teacher | 2,084 | 1,108 | 0.014 [−0.030, 0.059] SE 0.023 *p* = .543 *q* = .583 | −0.035 [−0.102, 0.036] SE 0.035 *p* = .323 *q* = .639 | 0.029 [−0.025, 0.085] SE 0.028 *p* = .300 *q* = .367 | 0.049 [−0.024, 0.117] SE 0.036 *p* = .170 *q* = .376 |
| Age 9 Self | 2,373 | 1,218 | 0.030 [−0.014, 0.072] SE 0.022 *p* = .173 *q* = .230 | 0.003 [−0.064, 0.070] SE 0.034 *p* = .920 *q* = .981 | 0.038 [−0.014, 0.091] SE 0.027 *p* = .159 *q* = .225 | 0.026 [−0.037, 0.092] SE 0.033 *p* = .421 *q* = .659 |
| Age 12 Parent | 4,624 | 2,312 | 0.029 [−0.003, 0.059] SE 0.016 *p* = .070 *q* = .112 | 0.020 [−0.027, 0.066] SE 0.024 *p* = .405 *q* = .694 | 0.032 [−0.009, 0.071] SE 0.020 *p* = .110 *q* = .164 | 0.009 [−0.037, 0.055] SE 0.023 *p* = .698 *q* = .892 |
| Age 12 Teacher | 3,699 | 2,064 | −0.014 [−0.048, 0.021] SE 0.018 *p* = .437 *q* = .507 | 0.019 [−0.041, 0.078] SE 0.030 *p* = .529 *q* = .794 | −0.024 [−0.065, 0.017] SE 0.021 *p* = .253 *q* = .331 | −0.033 [−0.087, 0.023] SE 0.028 *p* = .243 *q* = .465 |
| Age 16 Parent | 4,013 | 2,008 | 0.067 [0.034, 0.100] SE 0.017 *p* < .001 *q* < .001 | 0.038 [−0.007, 0.082] SE 0.023 *p* = .092 *q* = .276 | 0.078 [0.036, 0.118] SE 0.021 *p* < .001 *q* < .001 | 0.029 [−0.017, 0.074] SE 0.023 *p* = .204 *q* = .419 |
| Age 16 Self | 4,004 | 2,018 | −0.016 [−0.048, 0.016] SE 0.016 *p* = .317 *q* = .387 | −0.040 [−0.095, 0.015] SE 0.028 *p* = .151 *q* = .418 | −0.008 [−0.046, 0.031] SE 0.019 *p* = .674 *q* = .713 | 0.024 [−0.026, 0.072] SE 0.025 *p* = .343 *q* = .575 |
| Age 26 Self | 2,981 | 1,944 | 0.084 [0.049, 0.120] SE 0.018 *p* < .001 *q* < .001 | 0.017 [−0.057, 0.089] SE 0.037 *p* = .641 *q* = .887 | 0.099 [0.058, 0.141] SE 0.021 *p* < .001 *q* < .001 | 0.067 [−0.001, 0.136] SE 0.035 *p* = .055 *q* = .208 |

**Notes:** Values are standardised regression coefficients (β), with 95% confidence intervals (CIs) in brackets, followed by standard errors (SE) and two-sided *p* values. N, number of offspring; Families, number of family clusters. Population, association of offspring PGS in the population model; Direct, within-family coefficient (child deviation from twin-pair mean PGS); Between, between-family coefficient (twin-pair mean PGS); Indirect, population minus within-family coefficient. SEs and percentile 95% CIs were obtained by bootstrapping with families as the resampling unit; *p* values were derived from z = β/SE using bootstrap SEs. False discovery rate (FDR)-adjusted *p* values (*q*; Benjamini–Hochberg) were computed separately for each estimate type across all PGS and outcomes in this table. Models adjust for sex, genotyping chip and the first ten genetic principal components.

**Supplementary Table S4. Twin-comparison analyses of ASD traits** *(continued)*

#### Cognitive PGS · Demange et al. (2021)

| Outcome (age, reporter) | N (offspring) | Families (clusters) | Population (β, 95% CI, SE, *p*, *q*) | Direct (β, 95% CI, SE, *p*, *q*) | Between (β, 95% CI, SE, *p*, *q*) | Indirect (β, 95% CI, SE, *p*, *q*) |
| --- | --- | --- | --- | --- | --- | --- |
| Age 8 Parent | 4,802 | 2,402 | −0.051 [−0.080, −0.022] SE 0.015 *p* < .001 *q* = .002 | 0.020 [−0.029, 0.067] SE 0.025 *p* = .420 *q* = .703 | −0.074 [−0.110, −0.038] SE 0.019 *p* < .001 *q* < .001 | −0.071 [−0.116, −0.025] SE 0.024 *p* = .003 *q* = .028 |
| Age 9 Parent | 2,474 | 1,237 | −0.069 [−0.111, −0.027] SE 0.022 *p* = .002 *q* = .004 | 0.016 [−0.046, 0.075] SE 0.030 *p* = .598 *q* = .862 | −0.098 [−0.151, −0.046] SE 0.027 *p* < .001 *q* = .001 | −0.085 [−0.145, −0.024] SE 0.031 *p* = .006 *q* = .045 |
| Age 9 Teacher | 2,084 | 1,108 | −0.038 [−0.084, 0.007] SE 0.023 *p* = .098 *q* = .148 | 0.010 [−0.055, 0.075] SE 0.034 *p* = .776 *q* = .946 | −0.052 [−0.109, 0.003] SE 0.028 *p* = .063 *q* = .106 | −0.048 [−0.115, 0.020] SE 0.034 *p* = .165 *q* = .376 |
| Age 9 Self | 2,373 | 1,218 | 0.003 [−0.039, 0.046] SE 0.021 *p* = .886 *q* = .909 | 0.081 [0.012, 0.151] SE 0.036 *p* = .025 *q* = .138 | −0.023 [−0.073, 0.026] SE 0.026 *p* = .376 *q* = .430 | −0.078 [−0.142, −0.014] SE 0.033 *p* = .016 *q* = .099 |
| Age 12 Parent | 4,624 | 2,312 | −0.066 [−0.096, −0.036] SE 0.016 *p* < .001 *q* < .001 | 0.001 [−0.043, 0.047] SE 0.023 *p* = .954 *q* = .981 | −0.088 [−0.127, −0.050] SE 0.020 *p* < .001 *q* < .001 | −0.067 [−0.112, −0.023] SE 0.023 *p* = .004 *q* = .034 |
| Age 12 Teacher | 3,699 | 2,064 | 0.039 [0.007, 0.072] SE 0.017 *p* = .022 *q* = .037 | 0.036 [−0.021, 0.093] SE 0.029 *p* = .221 *q* = .511 | 0.040 [0.001, 0.079] SE 0.020 *p* = .046 *q* = .080 | 0.003 [−0.050, 0.057] SE 0.028 *p* = .904 *q* = .951 |
| Age 16 Parent | 4,013 | 2,008 | −0.023 [−0.057, 0.010] SE 0.017 *p* = .176 *q* = .230 | 0.023 [−0.022, 0.066] SE 0.022 *p* = .302 *q* = .639 | −0.039 [−0.082, 0.004] SE 0.022 *p* = .076 *q* = .125 | −0.046 [−0.091, 0.001] SE 0.023 *p* = .047 *q* = .189 |
| Age 16 Self | 4,004 | 2,018 | 0.082 [0.050, 0.114] SE 0.016 *p* < .001 *q* < .001 | 0.091 [0.038, 0.146] SE 0.028 *p* = .001 *q* = .013 | 0.079 [0.040, 0.116] SE 0.019 *p* < .001 *q* < .001 | −0.009 [−0.060, 0.039] SE 0.025 *p* = .719 *q* = .892 |
| Age 26 Self | 2,981 | 1,944 | −0.012 [−0.050, 0.026] SE 0.019 *p* = .528 *q* = .576 | −0.014 [−0.091, 0.063] SE 0.039 *p* = .726 *q* = .940 | −0.012 [−0.054, 0.030] SE 0.022 *p* = .585 *q* = .639 | 0.002 [−0.067, 0.074] SE 0.036 *p* = .963 *q* = .963 |

**Notes:** Values are standardised regression coefficients (β), with 95% confidence intervals (CIs) in brackets, followed by standard errors (SE) and two-sided *p* values. N, number of offspring; Families, number of family clusters. Population, association of offspring PGS in the population model; Direct, within-family coefficient (child deviation from twin-pair mean PGS); Between, between-family coefficient (twin-pair mean PGS); Indirect, population minus within-family coefficient. SEs and percentile 95% CIs were obtained by bootstrapping with families as the resampling unit; *p* values were derived from z = β/SE using bootstrap SEs. False discovery rate (FDR)-adjusted *p* values (*q*; Benjamini–Hochberg) were computed separately for each estimate type across all PGS and outcomes in this table. Models adjust for sex, genotyping chip and the first ten genetic principal components.

**Supplementary Table S4. Twin-comparison analyses of ASD traits** *(continued)*

#### Educational attainment PGS · Okbay et al. (2022)

| Outcome (age, reporter) | N (offspring) | Families (clusters) | Population (β, 95% CI, SE, *p*, *q*) | Direct (β, 95% CI, SE, *p*, *q*) | Between (β, 95% CI, SE, *p*, *q*) | Indirect (β, 95% CI, SE, *p*, *q*) |
| --- | --- | --- | --- | --- | --- | --- |
| Age 8 Parent | 4,802 | 2,402 | −0.087 [−0.118, −0.056] SE 0.016 *p* < .001 *q* < .001 | 0.082 [0.032, 0.131] SE 0.025 *p* = .001 *q* = .013 | −0.130 [−0.165, −0.094] SE 0.018 *p* < .001 *q* < .001 | −0.169 [−0.216, −0.121] SE 0.025 *p* < .001 *q* < .001 |
| Age 9 Parent | 2,474 | 1,237 | −0.106 [−0.152, −0.059] SE 0.023 *p* < .001 *q* < .001 | 0.043 [−0.019, 0.104] SE 0.031 *p* = .175 *q* = .434 | −0.143 [−0.198, −0.088] SE 0.028 *p* < .001 *q* < .001 | −0.149 [−0.212, −0.083] SE 0.033 *p* < .001 *q* < .001 |
| Age 9 Teacher | 2,084 | 1,108 | −0.017 [−0.065, 0.032] SE 0.025 *p* = .497 *q* = .559 | 0.044 [−0.033, 0.122] SE 0.040 *p* = .264 *q* = .577 | −0.031 [−0.087, 0.028] SE 0.029 *p* = .285 *q* = .354 | −0.061 [−0.141, 0.020] SE 0.041 *p* = .137 *q* = .367 |
| Age 9 Self | 2,373 | 1,218 | −0.015 [−0.057, 0.027] SE 0.021 *p* = .475 *q* = .543 | 0.094 [0.020, 0.170] SE 0.038 *p* = .013 *q* = .118 | −0.041 [−0.089, 0.007] SE 0.025 *p* = .101 *q* = .155 | −0.109 [−0.179, −0.040] SE 0.036 *p* = .002 *q* = .028 |
| Age 12 Parent | 4,624 | 2,312 | −0.096 [−0.128, −0.064] SE 0.016 *p* < .001 *q* < .001 | 0.032 [−0.020, 0.084] SE 0.026 *p* = .227 *q* = .511 | −0.128 [−0.165, −0.090] SE 0.019 *p* < .001 *q* < .001 | −0.128 [−0.177, −0.077] SE 0.025 *p* < .001 *q* < .001 |
| Age 12 Teacher | 3,699 | 2,064 | 0.005 [−0.031, 0.041] SE 0.018 *p* = .781 *q* = .827 | 0.033 [−0.031, 0.099] SE 0.034 *p* = .331 *q* = .639 | −0.001 [−0.041, 0.041] SE 0.021 *p* = .962 *q* = .970 | −0.028 [−0.092, 0.036] SE 0.033 *p* = .393 *q* = .642 |
| Age 16 Parent | 4,013 | 2,008 | −0.064 [−0.096, −0.030] SE 0.017 *p* < .001 *q* < .001 | 0.023 [−0.026, 0.070] SE 0.024 *p* = .354 *q* = .654 | −0.086 [−0.126, −0.046] SE 0.020 *p* < .001 *q* < .001 | −0.086 [−0.137, −0.037] SE 0.025 *p* < .001 *q* = .012 |
| Age 16 Self | 4,004 | 2,018 | 0.065 [0.033, 0.097] SE 0.016 *p* < .001 *q* < .001 | 0.081 [0.022, 0.139] SE 0.030 *p* = .007 *q* = .073 | 0.061 [0.025, 0.099] SE 0.019 *p* = .001 *q* = .004 | −0.016 [−0.071, 0.041] SE 0.029 *p* = .569 *q* = .773 |
| Age 26 Self | 2,981 | 1,944 | −0.084 [−0.122, −0.046] SE 0.020 *p* < .001 *q* < .001 | −0.008 [−0.093, 0.077] SE 0.044 *p* = .858 *q* = .951 | −0.096 [−0.139, −0.054] SE 0.022 *p* < .001 *q* < .001 | −0.076 [−0.156, 0.005] SE 0.042 *p* = .068 *q* = .223 |

**Notes:** Values are standardised regression coefficients (β), with 95% confidence intervals (CIs) in brackets, followed by standard errors (SE) and two-sided *p* values. N, number of offspring; Families, number of family clusters. Population, association of offspring PGS in the population model; Direct, within-family coefficient (child deviation from twin-pair mean PGS); Between, between-family coefficient (twin-pair mean PGS); Indirect, population minus within-family coefficient. SEs and percentile 95% CIs were obtained by bootstrapping with families as the resampling unit; *p* values were derived from z = β/SE using bootstrap SEs. False discovery rate (FDR)-adjusted *p* values (*q*; Benjamini–Hochberg) were computed separately for each estimate type across all PGS and outcomes in this table. Models adjust for sex, genotyping chip and the first ten genetic principal components.

### Supplementary Table S5. Trio-PGS analyses of ADHD traits

#### ADHD PGS · Demontis et al. (2023)

| Outcome (age, reporter) | N (offspring) | Families (clusters) | Child PGS (β, 95% CI, SE, *p*, *q*) | Maternal PGS (β, 95% CI, SE, *p*, *q*) | Paternal PGS (β, 95% CI, SE, *p*, *q*) |
| --- | --- | --- | --- | --- | --- |
| Age 4 Parent | 5,816 | 3,251 | 0.083 [0.045, 0.121] SE 0.020 *p* < .001 *q* < .001 | 0.013 [−0.017, 0.044] SE 0.016 *p* = .395 *q* = .782 | 0.016 [−0.018, 0.049] SE 0.017 *p* = .353 *q* = .727 |
| Age 7 Parent | 5,993 | 3,340 | 0.133 [0.096, 0.170] SE 0.019 *p* < .001 *q* < .001 | 0.015 [−0.014, 0.045] SE 0.015 *p* = .318 *q* = .731 | −0.025 [−0.057, 0.007] SE 0.016 *p* = .128 *q* = .727 |
| Age 7 Teacher | 4,846 | 2,762 | 0.168 [0.129, 0.207] SE 0.020 *p* < .001 *q* < .001 | −0.009 [−0.043, 0.025] SE 0.017 *p* = .593 *q* = .782 | 0.024 [−0.012, 0.061] SE 0.019 *p* = .193 *q* = .727 |
| Age 8 Parent | 5,437 | 3,044 | 0.145 [0.108, 0.183] SE 0.019 *p* < .001 *q* < .001 | −0.001 [−0.035, 0.033] SE 0.017 *p* = .970 *q* = .984 | 0.005 [−0.031, 0.041] SE 0.018 *p* = .793 *q* = .877 |
| Age 12 Parent | 5,173 | 2,917 | 0.133 [0.097, 0.169] SE 0.018 *p* < .001 *q* < .001 | 0.012 [−0.021, 0.045] SE 0.017 *p* = .486 *q* = .782 | 0.009 [−0.027, 0.045] SE 0.018 *p* = .608 *q* = .777 |
| Age 16 Parent | 4,589 | 2,611 | 0.113 [0.074, 0.151] SE 0.020 *p* < .001 *q* < .001 | 0.023 [−0.012, 0.058] SE 0.018 *p* = .206 *q* = .618 | 0.011 [−0.026, 0.048] SE 0.019 *p* = .552 *q* = .777 |
| Age 21 Parent | 3,990 | 2,447 | 0.068 [0.026, 0.110] SE 0.021 *p* = .001 *q* = .002 | 0.024 [−0.011, 0.060] SE 0.018 *p* = .179 *q* = .596 | 0.023 [−0.013, 0.060] SE 0.019 *p* = .211 *q* = .727 |
| Age 21 Self | 3,747 | 2,469 | 0.069 [0.023, 0.116] SE 0.024 *p* = .003 *q* = .007 | 0.029 [−0.007, 0.066] SE 0.019 *p* = .118 *q* = .596 | 0.022 [−0.016, 0.061] SE 0.020 *p* = .254 *q* = .727 |
| Age 26 Self | 1,736 | 1,334 | 0.074 [0.008, 0.140] SE 0.034 *p* = .028 *q* = .054 | 0.009 [−0.044, 0.063] SE 0.027 *p* = .729 *q* = .890 | −0.005 [−0.063, 0.053] SE 0.030 *p* = .867 *q* = .916 |

**Notes:** Values are standardised regression coefficients (β), with 95% confidence intervals (CIs) in brackets, followed by standard errors (SE) and two-sided *p* values. N, number of offspring; Families, number of family clusters. Child, maternal and paternal PGS were entered simultaneously; child PGS coefficients index direct genetic effects and parental PGS coefficients index indirect genetic effects. SEs are family-clustered robust estimates; 95% CIs and *p* values use family-cluster degrees of freedom. False discovery rate (FDR)-adjusted *p* values (*q*; Benjamini–Hochberg) were computed separately for each estimate type across all PGS and outcomes in this table. Models adjust for sex, genotyping chip and the first ten genetic principal components; age-specific models also adjust for age at assessment.

**Supplementary Table S5. Trio-PGS analyses of ADHD traits** *(continued)*

#### ASD PGS · Grove et al. (2019)

| Outcome (age, reporter) | N (offspring) | Families (clusters) | Child PGS (β, 95% CI, SE, *p*, *q*) | Maternal PGS (β, 95% CI, SE, *p*, *q*) | Paternal PGS (β, 95% CI, SE, *p*, *q*) |
| --- | --- | --- | --- | --- | --- |
| Age 4 Parent | 5,816 | 3,251 | 0.027 [−0.010, 0.065] SE 0.019 *p* = .156 *q* = .242 | 0.006 [−0.026, 0.037] SE 0.016 *p* = .729 *q* = .890 | −0.013 [−0.046, 0.019] SE 0.017 *p* = .422 *q* = .741 |
| Age 7 Parent | 5,993 | 3,340 | 0.029 [−0.007, 0.066] SE 0.019 *p* = .117 *q* = .195 | 0.018 [−0.013, 0.048] SE 0.016 *p* = .261 *q* = .707 | −0.033 [−0.064, −0.002] SE 0.016 *p* = .040 *q* = .594 |
| Age 7 Teacher | 4,846 | 2,762 | 0.022 [−0.017, 0.061] SE 0.020 *p* = .278 *q* = .378 | −0.010 [−0.044, 0.024] SE 0.017 *p* = .554 *q* = .782 | 0.028 [−0.008, 0.064] SE 0.018 *p* = .129 *q* = .727 |
| Age 8 Parent | 5,437 | 3,044 | 0.056 [0.019, 0.094] SE 0.019 *p* = .003 *q* = .007 | 0.011 [−0.021, 0.042] SE 0.016 *p* = .503 *q* = .782 | −0.011 [−0.047, 0.025] SE 0.018 *p* = .540 *q* = .777 |
| Age 12 Parent | 5,173 | 2,917 | 0.047 [0.010, 0.083] SE 0.018 *p* = .012 *q* = .025 | 0.017 [−0.016, 0.051] SE 0.017 *p* = .317 *q* = .731 | −0.008 [−0.045, 0.028] SE 0.019 *p* = .656 *q* = .791 |
| Age 16 Parent | 4,589 | 2,611 | 0.063 [0.025, 0.101] SE 0.019 *p* = .001 *q* = .002 | −0.011 [−0.047, 0.024] SE 0.018 *p* = .530 *q* = .782 | −0.014 [−0.052, 0.023] SE 0.019 *p* = .460 *q* = .752 |
| Age 21 Parent | 3,990 | 2,447 | 0.031 [−0.009, 0.070] SE 0.020 *p* = .126 *q* = .202 | 0.026 [−0.010, 0.061] SE 0.018 *p* = .162 *q* = .596 | −0.018 [−0.055, 0.019] SE 0.019 *p* = .341 *q* = .727 |
| Age 21 Self | 3,747 | 2,469 | 0.029 [−0.017, 0.075] SE 0.023 *p* = .217 *q* = .312 | 0.027 [−0.010, 0.064] SE 0.019 *p* = .147 *q* = .596 | 0.010 [−0.030, 0.050] SE 0.020 *p* = .615 *q* = .777 |
| Age 26 Self | 1,736 | 1,334 | 0.025 [−0.041, 0.092] SE 0.034 *p* = .453 *q* = .566 | −0.020 [−0.072, 0.032] SE 0.026 *p* = .457 *q* = .782 | 0.026 [−0.030, 0.081] SE 0.028 *p* = .363 *q* = .727 |

**Notes:** Values are standardised regression coefficients (β), with 95% confidence intervals (CIs) in brackets, followed by standard errors (SE) and two-sided *p* values. N, number of offspring; Families, number of family clusters. Child, maternal and paternal PGS were entered simultaneously; child PGS coefficients index direct genetic effects and parental PGS coefficients index indirect genetic effects. SEs are family-clustered robust estimates; 95% CIs and *p* values use family-cluster degrees of freedom. False discovery rate (FDR)-adjusted *p* values (*q*; Benjamini–Hochberg) were computed separately for each estimate type across all PGS and outcomes in this table. Models adjust for sex, genotyping chip and the first ten genetic principal components; age-specific models also adjust for age at assessment.

**Supplementary Table S5. Trio-PGS analyses of ADHD traits** *(continued)*

#### Depression PGS · Howard et al. (2019)

| Outcome (age, reporter) | N (offspring) | Families (clusters) | Child PGS (β, 95% CI, SE, *p*, *q*) | Maternal PGS (β, 95% CI, SE, *p*, *q*) | Paternal PGS (β, 95% CI, SE, *p*, *q*) |
| --- | --- | --- | --- | --- | --- |
| Age 4 Parent | 5,816 | 3,251 | 0.011 [−0.028, 0.051] SE 0.020 *p* = .573 *q* = .676 | 0.031 [0.000, 0.062] SE 0.016 *p* = .050 *q* = .416 | 0.013 [−0.020, 0.047] SE 0.017 *p* = .437 *q* = .749 |
| Age 7 Parent | 5,993 | 3,340 | 0.027 [−0.011, 0.065] SE 0.019 *p* = .158 *q* = .242 | 0.030 [0.000, 0.060] SE 0.015 *p* = .052 *q* = .416 | 0.001 [−0.031, 0.032] SE 0.016 *p* = .963 *q* = .963 |
| Age 7 Teacher | 4,846 | 2,762 | 0.007 [−0.033, 0.047] SE 0.020 *p* = .738 *q* = .830 | 0.017 [−0.019, 0.052] SE 0.018 *p* = .366 *q* = .782 | 0.034 [−0.004, 0.071] SE 0.019 *p* = .079 *q* = .632 |
| Age 8 Parent | 5,437 | 3,044 | 0.031 [−0.008, 0.070] SE 0.020 *p* = .119 *q* = .195 | 0.036 [0.001, 0.071] SE 0.018 *p* = .043 *q* = .416 | 0.016 [−0.020, 0.052] SE 0.018 *p* = .394 *q* = .727 |
| Age 12 Parent | 5,173 | 2,917 | 0.043 [0.005, 0.082] SE 0.020 *p* = .029 *q* = .055 | 0.047 [0.013, 0.081] SE 0.017 *p* = .006 *q* = .168 | 0.012 [−0.024, 0.049] SE 0.019 *p* = .512 *q* = .752 |
| Age 16 Parent | 4,589 | 2,611 | 0.040 [−0.003, 0.083] SE 0.022 *p* = .066 *q* = .115 | 0.033 [−0.003, 0.070] SE 0.019 *p* = .074 *q* = .533 | 0.008 [−0.031, 0.047] SE 0.020 *p* = .699 *q* = .812 |
| Age 21 Parent | 3,990 | 2,447 | 0.019 [−0.025, 0.064] SE 0.023 *p* = .395 *q* = .508 | 0.029 [−0.006, 0.064] SE 0.018 *p* = .101 *q* = .596 | 0.021 [−0.020, 0.062] SE 0.021 *p* = .322 *q* = .727 |
| Age 21 Self | 3,747 | 2,469 | 0.071 [0.026, 0.117] SE 0.023 *p* = .002 *q* = .005 | 0.015 [−0.022, 0.052] SE 0.019 *p* = .427 *q* = .782 | 0.003 [−0.037, 0.043] SE 0.020 *p* = .891 *q* = .916 |
| Age 26 Self | 1,736 | 1,334 | 0.085 [0.018, 0.153] SE 0.034 *p* = .013 *q* = .026 | −0.015 [−0.071, 0.041] SE 0.029 *p* = .595 *q* = .782 | 0.028 [−0.029, 0.084] SE 0.029 *p* = .335 *q* = .727 |

**Notes:** Values are standardised regression coefficients (β), with 95% confidence intervals (CIs) in brackets, followed by standard errors (SE) and two-sided *p* values. N, number of offspring; Families, number of family clusters. Child, maternal and paternal PGS were entered simultaneously; child PGS coefficients index direct genetic effects and parental PGS coefficients index indirect genetic effects. SEs are family-clustered robust estimates; 95% CIs and *p* values use family-cluster degrees of freedom. False discovery rate (FDR)-adjusted *p* values (*q*; Benjamini–Hochberg) were computed separately for each estimate type across all PGS and outcomes in this table. Models adjust for sex, genotyping chip and the first ten genetic principal components; age-specific models also adjust for age at assessment.

**Supplementary Table S5. Trio-PGS analyses of ADHD traits** *(continued)*

#### Externalising PGS · Karlsson Linnér et al. (2021)

| Outcome (age, reporter) | N (offspring) | Families (clusters) | Child PGS (β, 95% CI, SE, *p*, *q*) | Maternal PGS (β, 95% CI, SE, *p*, *q*) | Paternal PGS (β, 95% CI, SE, *p*, *q*) |
| --- | --- | --- | --- | --- | --- |
| Age 4 Parent | 5,816 | 3,251 | 0.070 [0.031, 0.109] SE 0.020 *p* < .001 *q* = .001 | −0.003 [−0.032, 0.027] SE 0.015 *p* = .867 *q* = .961 | −0.007 [−0.042, 0.027] SE 0.017 *p* = .669 *q* = .791 |
| Age 7 Parent | 5,993 | 3,340 | 0.079 [0.041, 0.117] SE 0.019 *p* < .001 *q* < .001 | 0.009 [−0.021, 0.039] SE 0.015 *p* = .539 *q* = .782 | 0.011 [−0.021, 0.042] SE 0.016 *p* = .511 *q* = .752 |
| Age 7 Teacher | 4,846 | 2,762 | 0.069 [0.028, 0.110] SE 0.021 *p* < .001 *q* = .002 | 0.026 [−0.010, 0.062] SE 0.018 *p* = .152 *q* = .596 | 0.052 [0.015, 0.089] SE 0.019 *p* = .005 *q* = .360 |
| Age 8 Parent | 5,437 | 3,044 | 0.127 [0.088, 0.166] SE 0.020 *p* < .001 *q* < .001 | −0.011 [−0.046, 0.023] SE 0.018 *p* = .515 *q* = .782 | −0.015 [−0.050, 0.020] SE 0.018 *p* = .405 *q* = .729 |
| Age 12 Parent | 5,173 | 2,917 | 0.123 [0.085, 0.161] SE 0.019 *p* < .001 *q* < .001 | −0.005 [−0.038, 0.029] SE 0.017 *p* = .787 *q* = .929 | −0.016 [−0.051, 0.020] SE 0.018 *p* = .384 *q* = .727 |
| Age 16 Parent | 4,589 | 2,611 | 0.104 [0.064, 0.144] SE 0.020 *p* < .001 *q* < .001 | 0.018 [−0.018, 0.055] SE 0.019 *p* = .325 *q* = .731 | 0.017 [−0.019, 0.053] SE 0.018 *p* = .357 *q* = .727 |
| Age 21 Parent | 3,990 | 2,447 | 0.111 [0.069, 0.153] SE 0.021 *p* < .001 *q* < .001 | 0.019 [−0.016, 0.054] SE 0.018 *p* = .295 *q* = .731 | −0.036 [−0.073, 0.001] SE 0.019 *p* = .059 *q* = .594 |
| Age 21 Self | 3,747 | 2,469 | 0.108 [0.060, 0.155] SE 0.024 *p* < .001 *q* < .001 | 0.011 [−0.026, 0.047] SE 0.019 *p* = .571 *q* = .782 | −0.006 [−0.046, 0.033] SE 0.020 *p* = .758 *q* = .853 |
| Age 26 Self | 1,736 | 1,334 | 0.143 [0.074, 0.212] SE 0.035 *p* < .001 *q* < .001 | −0.008 [−0.056, 0.039] SE 0.024 *p* = .728 *q* = .890 | −0.034 [−0.090, 0.023] SE 0.029 *p* = .241 *q* = .727 |

**Notes:** Values are standardised regression coefficients (β), with 95% confidence intervals (CIs) in brackets, followed by standard errors (SE) and two-sided *p* values. N, number of offspring; Families, number of family clusters. Child, maternal and paternal PGS were entered simultaneously; child PGS coefficients index direct genetic effects and parental PGS coefficients index indirect genetic effects. SEs are family-clustered robust estimates; 95% CIs and *p* values use family-cluster degrees of freedom. False discovery rate (FDR)-adjusted *p* values (*q*; Benjamini–Hochberg) were computed separately for each estimate type across all PGS and outcomes in this table. Models adjust for sex, genotyping chip and the first ten genetic principal components; age-specific models also adjust for age at assessment.

**Supplementary Table S5. Trio-PGS analyses of ADHD traits** *(continued)*

#### Anxiety PGS · Purves et al. (2020)

| Outcome (age, reporter) | N (offspring) | Families (clusters) | Child PGS (β, 95% CI, SE, *p*, *q*) | Maternal PGS (β, 95% CI, SE, *p*, *q*) | Paternal PGS (β, 95% CI, SE, *p*, *q*) |
| --- | --- | --- | --- | --- | --- |
| Age 4 Parent | 5,816 | 3,251 | −0.005 [−0.042, 0.033] SE 0.019 *p* = .813 *q* = .869 | −0.010 [−0.040, 0.020] SE 0.015 *p* = .516 *q* = .782 | −0.015 [−0.046, 0.017] SE 0.016 *p* = .358 *q* = .727 |
| Age 7 Parent | 5,993 | 3,340 | −0.023 [−0.059, 0.013] SE 0.018 *p* = .211 *q* = .310 | −0.017 [−0.046, 0.013] SE 0.015 *p* = .265 *q* = .707 | 0.001 [−0.029, 0.031] SE 0.015 *p* = .944 *q* = .957 |
| Age 7 Teacher | 4,846 | 2,762 | 0.001 [−0.036, 0.039] SE 0.019 *p* = .947 *q* = .960 | −0.002 [−0.038, 0.034] SE 0.018 *p* = .913 *q* = .975 | −0.016 [−0.052, 0.019] SE 0.018 *p* = .366 *q* = .727 |
| Age 8 Parent | 5,437 | 3,044 | 0.004 [−0.033, 0.042] SE 0.019 *p* = .821 *q* = .869 | −0.024 [−0.059, 0.011] SE 0.018 *p* = .177 *q* = .596 | −0.020 [−0.053, 0.014] SE 0.017 *p* = .255 *q* = .727 |
| Age 12 Parent | 5,173 | 2,917 | −0.014 [−0.052, 0.024] SE 0.019 *p* = .464 *q* = .566 | −0.025 [−0.060, 0.010] SE 0.018 *p* = .157 *q* = .596 | −0.015 [−0.050, 0.019] SE 0.018 *p* = .393 *q* = .727 |
| Age 16 Parent | 4,589 | 2,611 | −0.039 [−0.079, 0.001] SE 0.020 *p* = .054 *q* = .097 | 0.002 [−0.033, 0.037] SE 0.018 *p* = .911 *q* = .975 | −0.027 [−0.064, 0.010] SE 0.019 *p* = .160 *q* = .727 |
| Age 21 Parent | 3,990 | 2,447 | −0.012 [−0.056, 0.033] SE 0.023 *p* = .608 *q* = .706 | −0.027 [−0.062, 0.008] SE 0.018 *p* = .126 *q* = .596 | −0.014 [−0.052, 0.025] SE 0.020 *p* = .487 *q* = .752 |
| Age 21 Self | 3,747 | 2,469 | −0.045 [−0.090, 0.000] SE 0.023 *p* = .051 *q* = .094 | 0.012 [−0.024, 0.048] SE 0.018 *p* = .524 *q* = .782 | −0.017 [−0.055, 0.020] SE 0.019 *p* = .369 *q* = .727 |
| Age 26 Self | 1,736 | 1,334 | −0.025 [−0.091, 0.041] SE 0.034 *p* = .456 *q* = .566 | −0.001 [−0.050, 0.047] SE 0.025 *p* = .960 *q* = .984 | −0.035 [−0.089, 0.020] SE 0.028 *p* = .211 *q* = .727 |

**Notes:** Values are standardised regression coefficients (β), with 95% confidence intervals (CIs) in brackets, followed by standard errors (SE) and two-sided *p* values. N, number of offspring; Families, number of family clusters. Child, maternal and paternal PGS were entered simultaneously; child PGS coefficients index direct genetic effects and parental PGS coefficients index indirect genetic effects. SEs are family-clustered robust estimates; 95% CIs and *p* values use family-cluster degrees of freedom. False discovery rate (FDR)-adjusted *p* values (*q*; Benjamini–Hochberg) were computed separately for each estimate type across all PGS and outcomes in this table. Models adjust for sex, genotyping chip and the first ten genetic principal components; age-specific models also adjust for age at assessment.

**Supplementary Table S5. Trio-PGS analyses of ADHD traits** *(continued)*

#### Neuroticism PGS · Nagel et al. (2018)

| Outcome (age, reporter) | N (offspring) | Families (clusters) | Child PGS (β, 95% CI, SE, *p*, *q*) | Maternal PGS (β, 95% CI, SE, *p*, *q*) | Paternal PGS (β, 95% CI, SE, *p*, *q*) |
| --- | --- | --- | --- | --- | --- |
| Age 4 Parent | 5,816 | 3,251 | 0.005 [−0.033, 0.042] SE 0.019 *p* = .806 *q* = .869 | 0.037 [0.007, 0.067] SE 0.015 *p* = .017 *q* = .274 | 0.007 [−0.025, 0.039] SE 0.016 *p* = .670 *q* = .791 |
| Age 7 Parent | 5,993 | 3,340 | 0.000 [−0.036, 0.037] SE 0.019 *p* = .982 *q* = .982 | 0.049 [0.018, 0.080] SE 0.016 *p* = .002 *q* = .144 | −0.011 [−0.043, 0.020] SE 0.016 *p* = .470 *q* = .752 |
| Age 7 Teacher | 4,846 | 2,762 | −0.019 [−0.060, 0.021] SE 0.021 *p* = .355 *q* = .465 | 0.015 [−0.021, 0.052] SE 0.019 *p* = .413 *q* = .782 | 0.016 [−0.019, 0.052] SE 0.018 *p* = .368 *q* = .727 |
| Age 8 Parent | 5,437 | 3,044 | −0.005 [−0.043, 0.033] SE 0.019 *p* = .793 *q* = .869 | 0.043 [0.007, 0.079] SE 0.018 *p* = .019 *q* = .274 | 0.006 [−0.029, 0.042] SE 0.018 *p* = .732 *q* = .837 |
| Age 12 Parent | 5,173 | 2,917 | 0.026 [−0.012, 0.063] SE 0.019 *p* = .181 *q* = .272 | 0.047 [0.013, 0.081] SE 0.017 *p* = .007 *q* = .168 | −0.009 [−0.044, 0.026] SE 0.018 *p* = .614 *q* = .777 |
| Age 16 Parent | 4,589 | 2,611 | 0.025 [−0.016, 0.065] SE 0.021 *p* = .229 *q* = .317 | 0.026 [−0.009, 0.060] SE 0.018 *p* = .148 *q* = .596 | −0.021 [−0.057, 0.016] SE 0.019 *p* = .275 *q* = .727 |
| Age 21 Parent | 3,990 | 2,447 | −0.021 [−0.062, 0.020] SE 0.021 *p* = .322 *q* = .429 | 0.036 [0.002, 0.069] SE 0.017 *p* = .037 *q* = .416 | 0.030 [−0.009, 0.069] SE 0.020 *p* = .132 *q* = .727 |
| Age 21 Self | 3,747 | 2,469 | 0.015 [−0.028, 0.058] SE 0.022 *p* = .506 *q* = .607 | 0.029 [−0.006, 0.064] SE 0.018 *p* = .105 *q* = .596 | 0.009 [−0.031, 0.048] SE 0.020 *p* = .666 *q* = .791 |
| Age 26 Self | 1,736 | 1,334 | 0.039 [−0.023, 0.101] SE 0.032 *p* = .221 *q* = .312 | −0.008 [−0.064, 0.047] SE 0.028 *p* = .766 *q* = .919 | 0.005 [−0.052, 0.063] SE 0.029 *p* = .855 *q* = .916 |

**Notes:** Values are standardised regression coefficients (β), with 95% confidence intervals (CIs) in brackets, followed by standard errors (SE) and two-sided *p* values. N, number of offspring; Families, number of family clusters. Child, maternal and paternal PGS were entered simultaneously; child PGS coefficients index direct genetic effects and parental PGS coefficients index indirect genetic effects. SEs are family-clustered robust estimates; 95% CIs and *p* values use family-cluster degrees of freedom. False discovery rate (FDR)-adjusted *p* values (*q*; Benjamini–Hochberg) were computed separately for each estimate type across all PGS and outcomes in this table. Models adjust for sex, genotyping chip and the first ten genetic principal components; age-specific models also adjust for age at assessment.

**Supplementary Table S5. Trio-PGS analyses of ADHD traits** *(continued)*

#### Cognitive PGS · Demange et al. (2021)

| Outcome (age, reporter) | N (offspring) | Families (clusters) | Child PGS (β, 95% CI, SE, *p*, *q*) | Maternal PGS (β, 95% CI, SE, *p*, *q*) | Paternal PGS (β, 95% CI, SE, *p*, *q*) |
| --- | --- | --- | --- | --- | --- |
| Age 4 Parent | 5,816 | 3,251 | −0.061 [−0.097, −0.024] SE 0.019 *p* = .001 *q* = .002 | 0.003 [−0.027, 0.033] SE 0.015 *p* = .841 *q* = .961 | −0.016 [−0.047, 0.015] SE 0.016 *p* = .322 *q* = .727 |
| Age 7 Parent | 5,993 | 3,340 | −0.071 [−0.107, −0.035] SE 0.018 *p* < .001 *q* < .001 | −0.003 [−0.032, 0.027] SE 0.015 *p* = .868 *q* = .961 | −0.018 [−0.049, 0.014] SE 0.016 *p* = .271 *q* = .727 |
| Age 7 Teacher | 4,846 | 2,762 | −0.101 [−0.140, −0.063] SE 0.020 *p* < .001 *q* < .001 | −0.017 [−0.051, 0.016] SE 0.017 *p* = .314 *q* = .731 | 0.010 [−0.026, 0.045] SE 0.018 *p* = .592 *q* = .777 |
| Age 8 Parent | 5,437 | 3,044 | −0.072 [−0.111, −0.033] SE 0.020 *p* < .001 *q* < .001 | 0.009 [−0.025, 0.043] SE 0.017 *p* = .604 *q* = .782 | −0.037 [−0.072, −0.002] SE 0.018 *p* = .037 *q* = .594 |
| Age 12 Parent | 5,173 | 2,917 | −0.077 [−0.113, −0.040] SE 0.019 *p* < .001 *q* < .001 | 0.013 [−0.022, 0.047] SE 0.018 *p* = .465 *q* = .782 | −0.021 [−0.056, 0.013] SE 0.018 *p* = .229 *q* = .727 |
| Age 16 Parent | 4,589 | 2,611 | −0.065 [−0.104, −0.025] SE 0.020 *p* = .001 *q* = .002 | 0.001 [−0.034, 0.037] SE 0.018 *p* = .942 *q* = .983 | −0.018 [−0.054, 0.017] SE 0.018 *p* = .317 *q* = .727 |
| Age 21 Parent | 3,990 | 2,447 | −0.061 [−0.101, −0.020] SE 0.021 *p* = .003 *q* = .007 | 0.002 [−0.034, 0.038] SE 0.018 *p* = .921 *q* = .975 | 0.011 [−0.028, 0.051] SE 0.020 *p* = .575 *q* = .777 |
| Age 21 Self | 3,747 | 2,469 | 0.005 [−0.041, 0.050] SE 0.023 *p* = .845 *q* = .876 | 0.016 [−0.020, 0.052] SE 0.018 *p* = .391 *q* = .782 | −0.005 [−0.042, 0.033] SE 0.019 *p* = .804 *q* = .877 |
| Age 26 Self | 1,736 | 1,334 | 0.006 [−0.056, 0.068] SE 0.032 *p* = .852 *q* = .876 | −0.016 [−0.068, 0.037] SE 0.027 *p* = .556 *q* = .782 | 0.021 [−0.034, 0.076] SE 0.028 *p* = .457 *q* = .752 |

**Notes:** Values are standardised regression coefficients (β), with 95% confidence intervals (CIs) in brackets, followed by standard errors (SE) and two-sided *p* values. N, number of offspring; Families, number of family clusters. Child, maternal and paternal PGS were entered simultaneously; child PGS coefficients index direct genetic effects and parental PGS coefficients index indirect genetic effects. SEs are family-clustered robust estimates; 95% CIs and *p* values use family-cluster degrees of freedom. False discovery rate (FDR)-adjusted *p* values (*q*; Benjamini–Hochberg) were computed separately for each estimate type across all PGS and outcomes in this table. Models adjust for sex, genotyping chip and the first ten genetic principal components; age-specific models also adjust for age at assessment.

**Supplementary Table S5. Trio-PGS analyses of ADHD traits** *(continued)*

#### Educational attainment PGS · Okbay et al. (2022)

| Outcome (age, reporter) | N (offspring) | Families (clusters) | Child PGS (β, 95% CI, SE, *p*, *q*) | Maternal PGS (β, 95% CI, SE, *p*, *q*) | Paternal PGS (β, 95% CI, SE, *p*, *q*) |
| --- | --- | --- | --- | --- | --- |
| Age 4 Parent | 5,816 | 3,251 | −0.100 [−0.140, −0.061] SE 0.020 *p* < .001 *q* < .001 | −0.022 [−0.055, 0.011] SE 0.017 *p* = .199 *q* = .618 | −0.032 [−0.064, 0.000] SE 0.016 *p* = .052 *q* = .594 |
| Age 7 Parent | 5,993 | 3,340 | −0.095 [−0.134, −0.056] SE 0.020 *p* < .001 *q* < .001 | −0.018 [−0.051, 0.014] SE 0.016 *p* = .262 *q* = .707 | −0.041 [−0.074, −0.009] SE 0.017 *p* = .012 *q* = .432 |
| Age 7 Teacher | 4,846 | 2,762 | −0.125 [−0.167, −0.083] SE 0.021 *p* < .001 *q* < .001 | 0.011 [−0.027, 0.049] SE 0.019 *p* = .569 *q* = .782 | −0.027 [−0.065, 0.010] SE 0.019 *p* = .150 *q* = .727 |
| Age 8 Parent | 5,437 | 3,044 | −0.095 [−0.136, −0.055] SE 0.021 *p* < .001 *q* < .001 | 0.005 [−0.031, 0.041] SE 0.018 *p* = .801 *q* = .930 | −0.033 [−0.069, 0.002] SE 0.018 *p* = .066 *q* = .594 |
| Age 12 Parent | 5,173 | 2,917 | −0.102 [−0.143, −0.060] SE 0.021 *p* < .001 *q* < .001 | 0.015 [−0.021, 0.051] SE 0.018 *p* = .428 *q* = .782 | −0.023 [−0.059, 0.014] SE 0.018 *p* = .220 *q* = .727 |
| Age 16 Parent | 4,589 | 2,611 | −0.098 [−0.142, −0.054] SE 0.022 *p* < .001 *q* < .001 | 0.011 [−0.030, 0.052] SE 0.021 *p* = .608 *q* = .782 | −0.037 [−0.073, 0.000] SE 0.019 *p* = .048 *q* = .594 |
| Age 21 Parent | 3,990 | 2,447 | −0.064 [−0.111, −0.018] SE 0.024 *p* = .007 *q* = .015 | 0.000 [−0.042, 0.043] SE 0.022 *p* = .991 *q* = .991 | 0.010 [−0.030, 0.050] SE 0.020 *p* = .612 *q* = .777 |
| Age 21 Self | 3,747 | 2,469 | −0.048 [−0.099, 0.003] SE 0.026 *p* = .067 *q* = .115 | 0.028 [−0.013, 0.068] SE 0.021 *p* = .182 *q* = .596 | 0.003 [−0.035, 0.041] SE 0.019 *p* = .889 *q* = .916 |
| Age 26 Self | 1,736 | 1,334 | −0.014 [−0.084, 0.055] SE 0.036 *p* = .686 *q* = .784 | 0.021 [−0.036, 0.078] SE 0.029 *p* = .467 *q* = .782 | 0.021 [−0.039, 0.080] SE 0.030 *p* = .495 *q* = .752 |

**Notes:** Values are standardised regression coefficients (β), with 95% confidence intervals (CIs) in brackets, followed by standard errors (SE) and two-sided *p* values. N, number of offspring; Families, number of family clusters. Child, maternal and paternal PGS were entered simultaneously; child PGS coefficients index direct genetic effects and parental PGS coefficients index indirect genetic effects. SEs are family-clustered robust estimates; 95% CIs and *p* values use family-cluster degrees of freedom. False discovery rate (FDR)-adjusted *p* values (*q*; Benjamini–Hochberg) were computed separately for each estimate type across all PGS and outcomes in this table. Models adjust for sex, genotyping chip and the first ten genetic principal components; age-specific models also adjust for age at assessment.

### Supplementary Table S6. Trio-PGS analyses of ASD traits

#### ADHD PGS · Demontis et al. (2023)

| Outcome (age, reporter) | N (offspring) | Families (clusters) | Child PGS (β, 95% CI, SE, *p*, *q*) | Maternal PGS (β, 95% CI, SE, *p*, *q*) | Paternal PGS (β, 95% CI, SE, *p*, *q*) |
| --- | --- | --- | --- | --- | --- |
| Age 8 Parent | 5,444 | 3,048 | 0.035 [−0.003, 0.073] SE 0.019 *p* = .067 *q* = .272 | 0.048 [0.014, 0.082] SE 0.017 *p* = .005 *q* = .036 | 0.032 [−0.004, 0.067] SE 0.018 *p* = .079 *q* = .356 |
| Age 9 Parent | 2,838 | 1,602 | 0.028 [−0.024, 0.080] SE 0.026 *p* = .289 *q* = .612 | 0.049 [0.001, 0.097] SE 0.024 *p* = .046 *q* = .127 | 0.047 [−0.006, 0.099] SE 0.027 *p* = .080 *q* = .356 |
| Age 9 Teacher | 2,132 | 1,265 | 0.047 [−0.013, 0.107] SE 0.030 *p* = .122 *q* = .366 | −0.009 [−0.065, 0.046] SE 0.028 *p* = .742 *q* = .812 | 0.040 [−0.019, 0.098] SE 0.030 *p* = .182 *q* = .485 |
| Age 9 Self | 2,730 | 1,576 | 0.028 [−0.026, 0.081] SE 0.027 *p* = .305 *q* = .620 | 0.038 [−0.010, 0.085] SE 0.024 *p* = .121 *q* = .251 | 0.009 [−0.040, 0.059] SE 0.025 *p* = .717 *q* = .915 |
| Age 12 Parent | 5,174 | 2,918 | 0.059 [0.020, 0.097] SE 0.020 *p* = .003 *q* = .024 | 0.035 [0.001, 0.068] SE 0.017 *p* = .043 *q* = .124 | 0.009 [−0.027, 0.046] SE 0.019 *p* = .616 *q* = .852 |
| Age 12 Teacher | 3,779 | 2,325 | 0.074 [0.027, 0.121] SE 0.024 *p* = .002 *q* = .018 | −0.011 [−0.048, 0.027] SE 0.019 *p* = .573 *q* = .709 | −0.014 [−0.053, 0.025] SE 0.020 *p* = .482 *q* = .752 |
| Age 16 Parent | 4,588 | 2,612 | 0.001 [−0.040, 0.042] SE 0.021 *p* = .963 *q* = .993 | 0.058 [0.019, 0.096] SE 0.020 *p* = .004 *q* = .036 | 0.020 [−0.020, 0.060] SE 0.020 *p* = .337 *q* = .735 |
| Age 16 Self | 4,608 | 2,627 | −0.015 [−0.057, 0.028] SE 0.022 *p* = .502 *q* = .723 | 0.043 [0.006, 0.080] SE 0.019 *p* = .023 *q* = .106 | −0.009 [−0.046, 0.028] SE 0.019 *p* = .627 *q* = .852 |
| Age 26 Self | 1,737 | 1,335 | −0.003 [−0.075, 0.069] SE 0.037 *p* = .932 *q* = .993 | 0.072 [0.022, 0.123] SE 0.026 *p* = .005 *q* = .036 | 0.023 [−0.037, 0.084] SE 0.031 *p* = .451 *q* = .752 |

**Notes:** Values are standardised regression coefficients (β), with 95% confidence intervals (CIs) in brackets, followed by standard errors (SE) and two-sided *p* values. N, number of offspring; Families, number of family clusters. Child, maternal and paternal PGS were entered simultaneously; child PGS coefficients index direct genetic effects and parental PGS coefficients index indirect genetic effects. SEs are family-clustered robust estimates; 95% CIs and *p* values use family-cluster degrees of freedom. False discovery rate (FDR)-adjusted *p* values (*q*; Benjamini–Hochberg) were computed separately for each estimate type across all PGS and outcomes in this table. Models adjust for sex, genotyping chip and the first ten genetic principal components; age-specific models also adjust for age at assessment.

**Supplementary Table S6. Trio-PGS analyses of ASD traits** *(continued)*

#### ASD PGS · Grove et al. (2019)

| Outcome (age, reporter) | N (offspring) | Families (clusters) | Child PGS (β, 95% CI, SE, *p*, *q*) | Maternal PGS (β, 95% CI, SE, *p*, *q*) | Paternal PGS (β, 95% CI, SE, *p*, *q*) |
| --- | --- | --- | --- | --- | --- |
| Age 8 Parent | 5,444 | 3,048 | 0.074 [0.036, 0.113] SE 0.020 *p* < .001 *q* = .003 | −0.007 [−0.040, 0.025] SE 0.017 *p* = .663 *q* = .772 | −0.023 [−0.059, 0.013] SE 0.018 *p* = .204 *q* = .525 |
| Age 9 Parent | 2,838 | 1,602 | 0.106 [0.053, 0.158] SE 0.027 *p* < .001 *q* = .002 | −0.031 [−0.081, 0.019] SE 0.026 *p* = .231 *q* = .378 | −0.079 [−0.129, −0.028] SE 0.026 *p* = .002 *q* = .072 |
| Age 9 Teacher | 2,132 | 1,265 | 0.069 [0.014, 0.124] SE 0.028 *p* = .015 *q* = .083 | −0.003 [−0.054, 0.048] SE 0.026 *p* = .913 *q* = .926 | −0.039 [−0.089, 0.012] SE 0.026 *p* = .131 *q* = .414 |
| Age 9 Self | 2,730 | 1,576 | 0.077 [0.022, 0.131] SE 0.028 *p* = .006 *q* = .036 | 0.003 [−0.045, 0.051] SE 0.025 *p* = .901 *q* = .926 | −0.040 [−0.092, 0.011] SE 0.026 *p* = .121 *q* = .414 |
| Age 12 Parent | 5,174 | 2,918 | 0.096 [0.057, 0.134] SE 0.019 *p* < .001 *q* < .001 | −0.023 [−0.057, 0.010] SE 0.017 *p* = .177 *q* = .335 | −0.012 [−0.047, 0.022] SE 0.018 *p* = .489 *q* = .752 |
| Age 12 Teacher | 3,779 | 2,325 | 0.087 [0.045, 0.130] SE 0.022 *p* < .001 *q* = .002 | −0.029 [−0.065, 0.007] SE 0.018 *p* = .118 *q* = .251 | −0.014 [−0.053, 0.025] SE 0.020 *p* = .494 *q* = .752 |
| Age 16 Parent | 4,588 | 2,612 | 0.058 [0.018, 0.098] SE 0.020 *p* = .005 *q* = .036 | 0.025 [−0.015, 0.064] SE 0.020 *p* = .220 *q* = .368 | −0.031 [−0.070, 0.008] SE 0.020 *p* = .121 *q* = .414 |
| Age 16 Self | 4,608 | 2,627 | 0.078 [0.037, 0.119] SE 0.021 *p* < .001 *q* = .003 | 0.024 [−0.013, 0.062] SE 0.019 *p* = .200 *q* = .351 | −0.035 [−0.072, 0.001] SE 0.018 *p* = .055 *q* = .356 |
| Age 26 Self | 1,737 | 1,335 | 0.052 [−0.014, 0.119] SE 0.034 *p* = .121 *q* = .366 | 0.009 [−0.046, 0.063] SE 0.028 *p* = .759 *q* = .816 | −0.007 [−0.060, 0.047] SE 0.027 *p* = .811 *q* = .942 |

**Notes:** Values are standardised regression coefficients (β), with 95% confidence intervals (CIs) in brackets, followed by standard errors (SE) and two-sided *p* values. N, number of offspring; Families, number of family clusters. Child, maternal and paternal PGS were entered simultaneously; child PGS coefficients index direct genetic effects and parental PGS coefficients index indirect genetic effects. SEs are family-clustered robust estimates; 95% CIs and *p* values use family-cluster degrees of freedom. False discovery rate (FDR)-adjusted *p* values (*q*; Benjamini–Hochberg) were computed separately for each estimate type across all PGS and outcomes in this table. Models adjust for sex, genotyping chip and the first ten genetic principal components; age-specific models also adjust for age at assessment.

**Supplementary Table S6. Trio-PGS analyses of ASD traits** *(continued)*

#### Depression PGS · Howard et al. (2019)

| Outcome (age, reporter) | N (offspring) | Families (clusters) | Child PGS (β, 95% CI, SE, *p*, *q*) | Maternal PGS (β, 95% CI, SE, *p*, *q*) | Paternal PGS (β, 95% CI, SE, *p*, *q*) |
| --- | --- | --- | --- | --- | --- |
| Age 8 Parent | 5,444 | 3,048 | 0.036 [−0.003, 0.075] SE 0.020 *p* = .068 *q* = .272 | 0.039 [0.004, 0.074] SE 0.018 *p* = .030 *q* = .106 | 0.001 [−0.036, 0.037] SE 0.018 *p* = .976 *q* = .990 |
| Age 9 Parent | 2,838 | 1,602 | 0.006 [−0.051, 0.063] SE 0.029 *p* = .836 *q* = .993 | 0.054 [0.004, 0.104] SE 0.025 *p* = .033 *q* = .108 | 0.029 [−0.025, 0.083] SE 0.028 *p* = .292 *q* = .673 |
| Age 9 Teacher | 2,132 | 1,265 | −0.045 [−0.108, 0.018] SE 0.032 *p* = .161 *q* = .446 | 0.030 [−0.028, 0.087] SE 0.029 *p* = .315 *q* = .472 | 0.047 [−0.015, 0.108] SE 0.031 *p* = .137 *q* = .414 |
| Age 9 Self | 2,730 | 1,576 | 0.052 [−0.003, 0.107] SE 0.028 *p* = .064 *q* = .272 | 0.017 [−0.032, 0.066] SE 0.025 *p* = .496 *q* = .649 | 0.008 [−0.042, 0.058] SE 0.026 *p* = .757 *q* = .922 |
| Age 12 Parent | 5,174 | 2,918 | 0.016 [−0.023, 0.055] SE 0.020 *p* = .416 *q* = .657 | 0.048 [0.014, 0.082] SE 0.017 *p* = .005 *q* = .036 | 0.016 [−0.021, 0.052] SE 0.019 *p* = .401 *q* = .752 |
| Age 12 Teacher | 3,779 | 2,325 | 0.027 [−0.017, 0.070] SE 0.022 *p* = .233 *q* = .541 | −0.017 [−0.058, 0.023] SE 0.020 *p* = .393 *q* = .541 | 0.015 [−0.023, 0.054] SE 0.020 *p* = .435 *q* = .752 |
| Age 16 Parent | 4,588 | 2,612 | 0.020 [−0.021, 0.062] SE 0.021 *p* = .341 *q* = .646 | 0.031 [−0.008, 0.071] SE 0.020 *p* = .119 *q* = .251 | 0.002 [−0.039, 0.043] SE 0.021 *p* = .926 *q* = .966 |
| Age 16 Self | 4,608 | 2,627 | 0.008 [−0.035, 0.051] SE 0.022 *p* = .722 *q* = .963 | 0.040 [0.001, 0.078] SE 0.019 *p* = .042 *q* = .124 | 0.010 [−0.030, 0.050] SE 0.020 *p* = .617 *q* = .852 |
| Age 26 Self | 1,737 | 1,335 | 0.028 [−0.040, 0.097] SE 0.035 *p* = .419 *q* = .657 | 0.076 [0.022, 0.130] SE 0.028 *p* = .006 *q* = .039 | 0.035 [−0.024, 0.094] SE 0.030 *p* = .243 *q* = .603 |

**Notes:** Values are standardised regression coefficients (β), with 95% confidence intervals (CIs) in brackets, followed by standard errors (SE) and two-sided *p* values. N, number of offspring; Families, number of family clusters. Child, maternal and paternal PGS were entered simultaneously; child PGS coefficients index direct genetic effects and parental PGS coefficients index indirect genetic effects. SEs are family-clustered robust estimates; 95% CIs and *p* values use family-cluster degrees of freedom. False discovery rate (FDR)-adjusted *p* values (*q*; Benjamini–Hochberg) were computed separately for each estimate type across all PGS and outcomes in this table. Models adjust for sex, genotyping chip and the first ten genetic principal components; age-specific models also adjust for age at assessment.

**Supplementary Table S6. Trio-PGS analyses of ASD traits** *(continued)*

#### Externalising PGS · Karlsson Linnér et al. (2021)

| Outcome (age, reporter) | N (offspring) | Families (clusters) | Child PGS (β, 95% CI, SE, *p*, *q*) | Maternal PGS (β, 95% CI, SE, *p*, *q*) | Paternal PGS (β, 95% CI, SE, *p*, *q*) |
| --- | --- | --- | --- | --- | --- |
| Age 8 Parent | 5,444 | 3,048 | 0.034 [−0.005, 0.072] SE 0.020 *p* = .087 *q* = .298 | 0.026 [−0.006, 0.059] SE 0.016 *p* = .110 *q* = .251 | −0.010 [−0.046, 0.026] SE 0.018 *p* = .575 *q* = .828 |
| Age 9 Parent | 2,838 | 1,602 | 0.043 [−0.008, 0.094] SE 0.026 *p* = .099 *q* = .324 | 0.017 [−0.031, 0.064] SE 0.024 *p* = .484 *q* = .645 | 0.004 [−0.049, 0.056] SE 0.027 *p* = .886 *q* = .952 |
| Age 9 Teacher | 2,132 | 1,265 | 0.008 [−0.056, 0.072] SE 0.032 *p* = .808 *q* = .993 | −0.011 [−0.067, 0.046] SE 0.029 *p* = .712 *q* = .801 | 0.059 [−0.003, 0.121] SE 0.032 *p* = .061 *q* = .356 |
| Age 9 Self | 2,730 | 1,576 | 0.023 [−0.033, 0.079] SE 0.029 *p* = .420 *q* = .657 | 0.022 [−0.025, 0.069] SE 0.024 *p* = .363 *q* = .520 | 0.045 [−0.006, 0.095] SE 0.026 *p* = .084 *q* = .356 |
| Age 12 Parent | 5,174 | 2,918 | 0.041 [0.003, 0.080] SE 0.020 *p* = .037 *q* = .190 | 0.033 [−0.002, 0.068] SE 0.018 *p* = .061 *q* = .163 | −0.021 [−0.057, 0.015] SE 0.018 *p* = .262 *q* = .629 |
| Age 12 Teacher | 3,779 | 2,325 | −0.002 [−0.049, 0.044] SE 0.024 *p* = .923 *q* = .993 | −0.006 [−0.044, 0.032] SE 0.019 *p* = .744 *q* = .812 | 0.028 [−0.013, 0.068] SE 0.021 *p* = .181 *q* = .485 |
| Age 16 Parent | 4,588 | 2,612 | −0.057 [−0.098, −0.017] SE 0.021 *p* = .006 *q* = .036 | 0.026 [−0.013, 0.065] SE 0.020 *p* = .194 *q* = .349 | −0.004 [−0.045, 0.037] SE 0.021 *p* = .840 *q* = .952 |
| Age 16 Self | 4,608 | 2,627 | −0.038 [−0.081, 0.005] SE 0.022 *p* = .082 *q* = .298 | −0.010 [−0.046, 0.026] SE 0.018 *p* = .575 *q* = .709 | −0.006 [−0.044, 0.032] SE 0.019 *p* = .768 *q* = .922 |
| Age 26 Self | 1,737 | 1,335 | 0.035 [−0.034, 0.104] SE 0.035 *p* = .320 *q* = .623 | 0.042 [−0.010, 0.094] SE 0.027 *p* = .112 *q* = .251 | 0.003 [−0.058, 0.064] SE 0.031 *p* = .923 *q* = .966 |

**Notes:** Values are standardised regression coefficients (β), with 95% confidence intervals (CIs) in brackets, followed by standard errors (SE) and two-sided *p* values. N, number of offspring; Families, number of family clusters. Child, maternal and paternal PGS were entered simultaneously; child PGS coefficients index direct genetic effects and parental PGS coefficients index indirect genetic effects. SEs are family-clustered robust estimates; 95% CIs and *p* values use family-cluster degrees of freedom. False discovery rate (FDR)-adjusted *p* values (*q*; Benjamini–Hochberg) were computed separately for each estimate type across all PGS and outcomes in this table. Models adjust for sex, genotyping chip and the first ten genetic principal components; age-specific models also adjust for age at assessment.

**Supplementary Table S6. Trio-PGS analyses of ASD traits** *(continued)*

#### Anxiety PGS · Purves et al. (2020)

| Outcome (age, reporter) | N (offspring) | Families (clusters) | Child PGS (β, 95% CI, SE, *p*, *q*) | Maternal PGS (β, 95% CI, SE, *p*, *q*) | Paternal PGS (β, 95% CI, SE, *p*, *q*) |
| --- | --- | --- | --- | --- | --- |
| Age 8 Parent | 5,444 | 3,048 | 0.006 [−0.031, 0.043] SE 0.019 *p* = .756 *q* = .980 | −0.043 [−0.078, −0.008] SE 0.018 *p* = .017 *q* = .087 | −0.015 [−0.048, 0.019] SE 0.017 *p* = .386 *q* = .752 |
| Age 9 Parent | 2,838 | 1,602 | 0.025 [−0.024, 0.075] SE 0.025 *p* = .310 *q* = .620 | −0.039 [−0.088, 0.010] SE 0.025 *p* = .119 *q* = .251 | −0.066 [−0.113, −0.019] SE 0.024 *p* = .006 *q* = .144 |
| Age 9 Teacher | 2,132 | 1,265 | 0.022 [−0.038, 0.083] SE 0.031 *p* = .471 *q* = .721 | −0.007 [−0.063, 0.050] SE 0.029 *p* = .820 *q* = .856 | −0.048 [−0.107, 0.011] SE 0.030 *p* = .113 *q* = .414 |
| Age 9 Self | 2,730 | 1,576 | 0.024 [−0.031, 0.079] SE 0.028 *p* = .396 *q* = .657 | −0.006 [−0.055, 0.043] SE 0.025 *p* = .809 *q* = .856 | −0.034 [−0.081, 0.014] SE 0.024 *p* = .165 *q* = .475 |
| Age 12 Parent | 5,174 | 2,918 | 0.014 [−0.025, 0.053] SE 0.020 *p* = .481 *q* = .721 | −0.038 [−0.071, −0.004] SE 0.017 *p* = .029 *q* = .106 | −0.043 [−0.078, −0.007] SE 0.018 *p* = .019 *q* = .228 |
| Age 12 Teacher | 3,779 | 2,325 | 0.000 [−0.044, 0.043] SE 0.022 *p* = .990 *q* = .993 | −0.018 [−0.057, 0.021] SE 0.020 *p* = .360 *q* = .520 | −0.029 [−0.068, 0.009] SE 0.020 *p* = .138 *q* = .414 |
| Age 16 Parent | 4,588 | 2,612 | 0.014 [−0.026, 0.053] SE 0.020 *p* = .492 *q* = .723 | −0.054 [−0.091, −0.017] SE 0.019 *p* = .004 *q* = .036 | −0.016 [−0.056, 0.023] SE 0.020 *p* = .421 *q* = .752 |
| Age 16 Self | 4,608 | 2,627 | −0.004 [−0.045, 0.037] SE 0.021 *p* = .838 *q* = .993 | −0.017 [−0.053, 0.020] SE 0.019 *p* = .368 *q* = .520 | −0.016 [−0.054, 0.022] SE 0.019 *p* = .400 *q* = .752 |
| Age 26 Self | 1,737 | 1,335 | 0.010 [−0.056, 0.077] SE 0.034 *p* = .762 *q* = .980 | −0.035 [−0.091, 0.021] SE 0.028 *p* = .219 *q* = .368 | −0.024 [−0.082, 0.034] SE 0.029 *p* = .415 *q* = .752 |

**Notes:** Values are standardised regression coefficients (β), with 95% confidence intervals (CIs) in brackets, followed by standard errors (SE) and two-sided *p* values. N, number of offspring; Families, number of family clusters. Child, maternal and paternal PGS were entered simultaneously; child PGS coefficients index direct genetic effects and parental PGS coefficients index indirect genetic effects. SEs are family-clustered robust estimates; 95% CIs and *p* values use family-cluster degrees of freedom. False discovery rate (FDR)-adjusted *p* values (*q*; Benjamini–Hochberg) were computed separately for each estimate type across all PGS and outcomes in this table. Models adjust for sex, genotyping chip and the first ten genetic principal components; age-specific models also adjust for age at assessment.

**Supplementary Table S6. Trio-PGS analyses of ASD traits** *(continued)*

#### Neuroticism PGS · Nagel et al. (2018)

| Outcome (age, reporter) | N (offspring) | Families (clusters) | Child PGS (β, 95% CI, SE, *p*, *q*) | Maternal PGS (β, 95% CI, SE, *p*, *q*) | Paternal PGS (β, 95% CI, SE, *p*, *q*) |
| --- | --- | --- | --- | --- | --- |
| Age 8 Parent | 5,444 | 3,048 | 0.020 [−0.016, 0.056] SE 0.018 *p* = .288 *q* = .612 | 0.053 [0.017, 0.088] SE 0.018 *p* = .004 *q* = .036 | 0.000 [−0.036, 0.035] SE 0.018 *p* = .997 *q* = .997 |
| Age 9 Parent | 2,838 | 1,602 | −0.003 [−0.055, 0.048] SE 0.026 *p* = .899 *q* = .993 | 0.113 [0.064, 0.161] SE 0.025 *p* < .001 *q* < .001 | 0.016 [−0.037, 0.069] SE 0.027 *p* = .554 *q* = .814 |
| Age 9 Teacher | 2,132 | 1,265 | −0.040 [−0.102, 0.022] SE 0.032 *p* = .202 *q* = .519 | 0.062 [0.007, 0.118] SE 0.028 *p* = .027 *q* = .106 | 0.028 [−0.025, 0.081] SE 0.027 *p* = .299 *q* = .673 |
| Age 9 Self | 2,730 | 1,576 | 0.000 [−0.056, 0.056] SE 0.029 *p* = .993 *q* = .993 | 0.056 [0.007, 0.104] SE 0.025 *p* = .025 *q* = .106 | 0.008 [−0.040, 0.057] SE 0.025 *p* = .737 *q* = .915 |
| Age 12 Parent | 5,174 | 2,918 | 0.004 [−0.035, 0.043] SE 0.020 *p* = .855 *q* = .993 | 0.055 [0.021, 0.089] SE 0.017 *p* = .002 *q* = .036 | 0.003 [−0.032, 0.037] SE 0.018 *p* = .879 *q* = .952 |
| Age 12 Teacher | 3,779 | 2,325 | 0.004 [−0.041, 0.049] SE 0.023 *p* = .866 *q* = .993 | −0.011 [−0.050, 0.028] SE 0.020 *p* = .581 *q* = .709 | −0.031 [−0.068, 0.006] SE 0.019 *p* = .096 *q* = .384 |
| Age 16 Parent | 4,588 | 2,612 | 0.035 [−0.005, 0.075] SE 0.020 *p* = .084 *q* = .298 | 0.055 [0.015, 0.095] SE 0.021 *p* = .008 *q* = .044 | −0.004 [−0.044, 0.037] SE 0.020 *p* = .862 *q* = .952 |
| Age 16 Self | 4,608 | 2,627 | −0.029 [−0.071, 0.013] SE 0.021 *p* = .181 *q* = .483 | 0.026 [−0.010, 0.062] SE 0.018 *p* = .162 *q* = .315 | 0.014 [−0.025, 0.054] SE 0.020 *p* = .470 *q* = .752 |
| Age 26 Self | 1,737 | 1,335 | 0.041 [−0.024, 0.107] SE 0.033 *p* = .212 *q* = .526 | 0.038 [−0.018, 0.094] SE 0.029 *p* = .187 *q* = .345 | −0.013 [−0.069, 0.044] SE 0.029 *p* = .659 *q* = .879 |

**Notes:** Values are standardised regression coefficients (β), with 95% confidence intervals (CIs) in brackets, followed by standard errors (SE) and two-sided *p* values. N, number of offspring; Families, number of family clusters. Child, maternal and paternal PGS were entered simultaneously; child PGS coefficients index direct genetic effects and parental PGS coefficients index indirect genetic effects. SEs are family-clustered robust estimates; 95% CIs and *p* values use family-cluster degrees of freedom. False discovery rate (FDR)-adjusted *p* values (*q*; Benjamini–Hochberg) were computed separately for each estimate type across all PGS and outcomes in this table. Models adjust for sex, genotyping chip and the first ten genetic principal components; age-specific models also adjust for age at assessment.

**Supplementary Table S6. Trio-PGS analyses of ASD traits** *(continued)*

#### Cognitive PGS · Demange et al. (2021)

| Outcome (age, reporter) | N (offspring) | Families (clusters) | Child PGS (β, 95% CI, SE, *p*, *q*) | Maternal PGS (β, 95% CI, SE, *p*, *q*) | Paternal PGS (β, 95% CI, SE, *p*, *q*) |
| --- | --- | --- | --- | --- | --- |
| Age 8 Parent | 5,444 | 3,048 | −0.002 [−0.041, 0.037] SE 0.020 *p* = .913 *q* = .993 | −0.030 [−0.063, 0.003] SE 0.017 *p* = .076 *q* = .195 | −0.045 [−0.078, −0.012] SE 0.017 *p* = .008 *q* = .144 |
| Age 9 Parent | 2,838 | 1,602 | −0.025 [−0.080, 0.029] SE 0.028 *p* = .363 *q* = .657 | −0.021 [−0.070, 0.028] SE 0.025 *p* = .398 *q* = .541 | −0.048 [−0.098, 0.003] SE 0.026 *p* = .063 *q* = .356 |
| Age 9 Teacher | 2,132 | 1,265 | 0.002 [−0.056, 0.060] SE 0.030 *p* = .952 *q* = .993 | −0.019 [−0.074, 0.037] SE 0.028 *p* = .514 *q* = .661 | −0.022 [−0.072, 0.027] SE 0.025 *p* = .379 *q* = .752 |
| Age 9 Self | 2,730 | 1,576 | 0.053 [−0.001, 0.107] SE 0.027 *p* = .054 *q* = .259 | 0.011 [−0.038, 0.060] SE 0.025 *p* = .665 *q* = .772 | −0.045 [−0.094, 0.003] SE 0.025 *p* = .069 *q* = .356 |
| Age 12 Parent | 5,174 | 2,918 | −0.010 [−0.049, 0.029] SE 0.020 *p* = .611 *q* = .830 | −0.048 [−0.083, −0.013] SE 0.018 *p* = .007 *q* = .042 | −0.038 [−0.073, −0.003] SE 0.018 *p* = .034 *q* = .306 |
| Age 12 Teacher | 3,779 | 2,325 | 0.031 [−0.010, 0.072] SE 0.021 *p* = .138 *q* = .397 | 0.002 [−0.035, 0.038] SE 0.019 *p* = .933 *q* = .933 | 0.007 [−0.031, 0.044] SE 0.019 *p* = .725 *q* = .915 |
| Age 16 Parent | 4,588 | 2,612 | 0.017 [−0.023, 0.057] SE 0.021 *p* = .408 *q* = .657 | −0.020 [−0.059, 0.019] SE 0.020 *p* = .311 *q* = .472 | −0.046 [−0.085, −0.006] SE 0.020 *p* = .023 *q* = .237 |
| Age 16 Self | 4,608 | 2,627 | 0.074 [0.031, 0.116] SE 0.022 *p* < .001 *q* = .008 | −0.007 [−0.043, 0.029] SE 0.018 *p* = .709 *q* = .801 | 0.008 [−0.030, 0.046] SE 0.019 *p* = .677 *q* = .886 |
| Age 26 Self | 1,737 | 1,335 | −0.003 [−0.067, 0.060] SE 0.032 *p* = .919 *q* = .993 | −0.043 [−0.097, 0.011] SE 0.028 *p* = .122 *q* = .251 | 0.006 [−0.057, 0.069] SE 0.032 *p* = .857 *q* = .952 |

**Notes:** Values are standardised regression coefficients (β), with 95% confidence intervals (CIs) in brackets, followed by standard errors (SE) and two-sided *p* values. N, number of offspring; Families, number of family clusters. Child, maternal and paternal PGS were entered simultaneously; child PGS coefficients index direct genetic effects and parental PGS coefficients index indirect genetic effects. SEs are family-clustered robust estimates; 95% CIs and *p* values use family-cluster degrees of freedom. False discovery rate (FDR)-adjusted *p* values (*q*; Benjamini–Hochberg) were computed separately for each estimate type across all PGS and outcomes in this table. Models adjust for sex, genotyping chip and the first ten genetic principal components; age-specific models also adjust for age at assessment.

**Supplementary Table S6. Trio-PGS analyses of ASD traits** *(continued)*

#### Educational attainment PGS · Okbay et al. (2022)

| Outcome (age, reporter) | N (offspring) | Families (clusters) | Child PGS (β, 95% CI, SE, *p*, *q*) | Maternal PGS (β, 95% CI, SE, *p*, *q*) | Paternal PGS (β, 95% CI, SE, *p*, *q*) |
| --- | --- | --- | --- | --- | --- |
| Age 8 Parent | 5,444 | 3,048 | 0.022 [−0.017, 0.061] SE 0.020 *p* = .273 *q* = .612 | −0.081 [−0.116, −0.046] SE 0.018 *p* < .001 *q* < .001 | −0.086 [−0.120, −0.053] SE 0.017 *p* < .001 *q* < .001 |
| Age 9 Parent | 2,838 | 1,602 | −0.036 [−0.092, 0.021] SE 0.029 *p* = .219 *q* = .526 | −0.060 [−0.113, −0.007] SE 0.027 *p* = .026 *q* = .106 | −0.047 [−0.100, 0.006] SE 0.027 *p* = .083 *q* = .356 |
| Age 9 Teacher | 2,132 | 1,265 | 0.027 [−0.035, 0.088] SE 0.031 *p* = .392 *q* = .657 | −0.031 [−0.085, 0.024] SE 0.028 *p* = .268 *q* = .429 | −0.001 [−0.062, 0.059] SE 0.031 *p* = .965 *q* = .990 |
| Age 9 Self | 2,730 | 1,576 | 0.016 [−0.043, 0.074] SE 0.030 *p* = .606 *q* = .830 | −0.013 [−0.064, 0.038] SE 0.026 *p* = .620 *q* = .744 | −0.018 [−0.068, 0.031] SE 0.025 *p* = .472 *q* = .752 |
| Age 12 Parent | 5,174 | 2,918 | −0.001 [−0.042, 0.041] SE 0.021 *p* = .978 *q* = .993 | −0.092 [−0.128, −0.057] SE 0.018 *p* < .001 *q* < .001 | −0.046 [−0.082, −0.011] SE 0.018 *p* = .010 *q* = .144 |
| Age 12 Teacher | 3,779 | 2,325 | 0.016 [−0.032, 0.063] SE 0.024 *p* = .517 *q* = .730 | −0.030 [−0.069, 0.010] SE 0.020 *p* = .137 *q* = .274 | 0.016 [−0.024, 0.055] SE 0.020 *p* = .439 *q* = .752 |
| Age 16 Parent | 4,588 | 2,612 | −0.001 [−0.044, 0.042] SE 0.022 *p* = .967 *q* = .993 | −0.045 [−0.085, −0.004] SE 0.021 *p* = .031 *q* = .106 | −0.042 [−0.083, −0.001] SE 0.021 *p* = .045 *q* = .356 |
| Age 16 Self | 4,608 | 2,627 | 0.075 [0.031, 0.119] SE 0.023 *p* < .001 *q* = .009 | −0.023 [−0.063, 0.018] SE 0.021 *p* = .279 *q* = .437 | −0.006 [−0.046, 0.034] SE 0.020 *p* = .784 *q* = .925 |
| Age 26 Self | 1,737 | 1,335 | −0.032 [−0.102, 0.038] SE 0.036 *p* = .368 *q* = .657 | −0.057 [−0.112, −0.003] SE 0.028 *p* = .038 *q* = .119 | 0.021 [−0.040, 0.082] SE 0.031 *p* = .501 *q* = .752 |

**Notes:** Values are standardised regression coefficients (β), with 95% confidence intervals (CIs) in brackets, followed by standard errors (SE) and two-sided *p* values. N, number of offspring; Families, number of family clusters. Child, maternal and paternal PGS were entered simultaneously; child PGS coefficients index direct genetic effects and parental PGS coefficients index indirect genetic effects. SEs are family-clustered robust estimates; 95% CIs and *p* values use family-cluster degrees of freedom. False discovery rate (FDR)-adjusted *p* values (*q*; Benjamini–Hochberg) were computed separately for each estimate type across all PGS and outcomes in this table. Models adjust for sex, genotyping chip and the first ten genetic principal components; age-specific models also adjust for age at assessment.

### Supplementary Table S7. Twin-comparison analyses of ADHD and ASD latent factors

#### ADHD PGS · Demontis et al. (2023)

| Outcome (trait, reporter) | N (offspring) | Families (clusters) | Population (β, 95% CI, SE, *p*, *q*) | Direct (β, 95% CI, SE, *p*, *q*) | Between (β, 95% CI, SE, *p*, *q*) | Indirect (β, 95% CI, SE, *p*, *q*) |
| --- | --- | --- | --- | --- | --- | --- |
| ADHD Common | 6,504 | 3,264 | 0.169 [0.146, 0.193] SE 0.012 *p* < .001 *q* < .001 | 0.160 [0.123, 0.196] SE 0.019 *p* < .001 *q* < .001 | 0.173 [0.143, 0.202] SE 0.015 *p* < .001 *q* < .001 | 0.009 [−0.027, 0.045] SE 0.018 *p* = .604 *q* = .744 |
| ADHD Parent | 6,452 | 3,227 | 0.160 [0.135, 0.184] SE 0.013 *p* < .001 *q* < .001 | 0.151 [0.115, 0.188] SE 0.019 *p* < .001 *q* < .001 | 0.163 [0.132, 0.194] SE 0.016 *p* < .001 *q* < .001 | 0.009 [−0.027, 0.045] SE 0.018 *p* = .620 *q* = .749 |
| ADHD Self | 4,639 | 2,567 | 0.088 [0.057, 0.119] SE 0.015 *p* < .001 *q* < .001 | 0.090 [0.031, 0.149] SE 0.030 *p* = .003 *q* = .010 | 0.088 [0.053, 0.123] SE 0.018 *p* < .001 *q* < .001 | −0.002 [−0.055, 0.051] SE 0.027 *p* = .941 *q* = .980 |
| ADHD Teacher | 4,590 | 2,362 | 0.175 [0.145, 0.204] SE 0.015 *p* < .001 *q* < .001 | 0.156 [0.110, 0.203] SE 0.024 *p* < .001 *q* < .001 | 0.180 [0.144, 0.216] SE 0.018 *p* < .001 *q* < .001 | 0.018 [−0.027, 0.062] SE 0.023 *p* = .423 *q* = .629 |
| ASD Common | 6,188 | 3,122 | 0.096 [0.070, 0.122] SE 0.013 *p* < .001 *q* < .001 | 0.025 [−0.013, 0.063] SE 0.020 *p* = .198 *q* = .316 | 0.119 [0.086, 0.151] SE 0.017 *p* < .001 *q* < .001 | 0.071 [0.033, 0.110] SE 0.020 *p* < .001 *q* = .005 |
| ASD Parent | 5,940 | 2,970 | 0.089 [0.062, 0.117] SE 0.014 *p* < .001 *q* < .001 | 0.015 [−0.023, 0.052] SE 0.019 *p* = .445 *q* = .593 | 0.113 [0.079, 0.148] SE 0.017 *p* < .001 *q* < .001 | 0.075 [0.036, 0.114] SE 0.020 *p* < .001 *q* = .003 |
| ASD Self | 5,234 | 2,724 | 0.062 [0.034, 0.090] SE 0.014 *p* < .001 *q* < .001 | 0.024 [−0.028, 0.074] SE 0.026 *p* = .363 *q* = .504 | 0.075 [0.040, 0.109] SE 0.017 *p* < .001 *q* < .001 | 0.039 [−0.009, 0.086] SE 0.024 *p* = .109 *q* = .386 |
| ASD Teacher | 4,583 | 2,413 | 0.070 [0.040, 0.102] SE 0.016 *p* < .001 *q* < .001 | 0.049 [−0.003, 0.099] SE 0.026 *p* = .064 *q* = .132 | 0.077 [0.041, 0.115] SE 0.019 *p* < .001 *q* < .001 | 0.022 [−0.024, 0.071] SE 0.024 *p* = .363 *q* = .629 |

**Notes:** Values are standardised regression coefficients (β), with 95% confidence intervals (CIs) in brackets, followed by standard errors (SE) and two-sided *p* values. N, number of offspring; Families, number of family clusters. Population, association of offspring PGS in the population model; Direct, within-family coefficient (child deviation from twin-pair mean PGS); Between, between-family coefficient (twin-pair mean PGS); Indirect, population minus within-family coefficient. SEs and percentile 95% CIs were obtained by bootstrapping with families as the resampling unit; *p* values were derived from z = β/SE using bootstrap SEs. False discovery rate (FDR)-adjusted *p* values (*q*; Benjamini–Hochberg) were computed separately for each estimate type across all PGS and outcomes in this table. Models adjust for sex, genotyping chip and the first ten genetic principal components.

**Supplementary Table S7. Twin-comparison analyses of ADHD and ASD latent factors** *(continued)*

#### ASD PGS · Grove et al. (2019)

| Outcome (trait, reporter) | N (offspring) | Families (clusters) | Population (β, 95% CI, SE, *p*, *q*) | Direct (β, 95% CI, SE, *p*, *q*) | Between (β, 95% CI, SE, *p*, *q*) | Indirect (β, 95% CI, SE, *p*, *q*) |
| --- | --- | --- | --- | --- | --- | --- |
| ADHD Common | 6,504 | 3,264 | 0.051 [0.026, 0.076] SE 0.013 *p* < .001 *q* < .001 | 0.051 [0.010, 0.090] SE 0.021 *p* = .013 *q* = .032 | 0.051 [0.021, 0.081] SE 0.016 *p* = .001 *q* = .002 | 0.000 [−0.039, 0.038] SE 0.020 *p* = .993 *q* = .993 |
| ADHD Parent | 6,452 | 3,227 | 0.049 [0.024, 0.074] SE 0.013 *p* < .001 *q* < .001 | 0.050 [0.011, 0.089] SE 0.020 *p* = .012 *q* = .030 | 0.049 [0.018, 0.080] SE 0.016 *p* = .002 *q* = .003 | −0.001 [−0.039, 0.035] SE 0.019 *p* = .949 *q* = .980 |
| ADHD Self | 4,639 | 2,567 | 0.045 [0.016, 0.073] SE 0.015 *p* = .003 *q* = .004 | 0.006 [−0.051, 0.063] SE 0.029 *p* = .836 *q* = .877 | 0.057 [0.023, 0.089] SE 0.017 *p* < .001 *q* = .001 | 0.039 [−0.013, 0.089] SE 0.026 *p* = .137 *q* = .415 |
| ADHD Teacher | 4,590 | 2,362 | 0.036 [0.006, 0.066] SE 0.015 *p* = .016 *q* = .021 | 0.021 [−0.029, 0.070] SE 0.025 *p* = .392 *q* = .533 | 0.041 [0.006, 0.076] SE 0.018 *p* = .023 *q* = .029 | 0.015 [−0.031, 0.062] SE 0.024 *p* = .536 *q* = .686 |
| ASD Common | 6,188 | 3,122 | 0.073 [0.047, 0.099] SE 0.013 *p* < .001 *q* < .001 | 0.107 [0.068, 0.146] SE 0.020 *p* < .001 *q* < .001 | 0.062 [0.030, 0.095] SE 0.017 *p* < .001 *q* < .001 | −0.033 [−0.071, 0.005] SE 0.019 *p* = .086 *q* = .386 |
| ASD Parent | 5,940 | 2,970 | 0.070 [0.042, 0.098] SE 0.014 *p* < .001 *q* < .001 | 0.097 [0.057, 0.138] SE 0.020 *p* < .001 *q* < .001 | 0.061 [0.027, 0.095] SE 0.017 *p* < .001 *q* < .001 | −0.027 [−0.065, 0.011] SE 0.019 *p* = .162 *q* = .432 |
| ASD Self | 5,234 | 2,724 | 0.069 [0.041, 0.096] SE 0.014 *p* < .001 *q* < .001 | 0.100 [0.053, 0.146] SE 0.024 *p* < .001 *q* < .001 | 0.058 [0.025, 0.090] SE 0.017 *p* < .001 *q* = .001 | −0.031 [−0.074, 0.010] SE 0.021 *p* = .149 *q* = .415 |
| ASD Teacher | 4,583 | 2,413 | 0.048 [0.016, 0.080] SE 0.016 *p* = .003 *q* = .004 | 0.068 [0.021, 0.118] SE 0.025 *p* = .006 *q* = .018 | 0.042 [0.004, 0.079] SE 0.019 *p* = .027 *q* = .033 | −0.020 [−0.066, 0.026] SE 0.023 *p* = .390 *q* = .629 |

**Notes:** Values are standardised regression coefficients (β), with 95% confidence intervals (CIs) in brackets, followed by standard errors (SE) and two-sided *p* values. N, number of offspring; Families, number of family clusters. Population, association of offspring PGS in the population model; Direct, within-family coefficient (child deviation from twin-pair mean PGS); Between, between-family coefficient (twin-pair mean PGS); Indirect, population minus within-family coefficient. SEs and percentile 95% CIs were obtained by bootstrapping with families as the resampling unit; *p* values were derived from z = β/SE using bootstrap SEs. False discovery rate (FDR)-adjusted *p* values (*q*; Benjamini–Hochberg) were computed separately for each estimate type across all PGS and outcomes in this table. Models adjust for sex, genotyping chip and the first ten genetic principal components.

**Supplementary Table S7. Twin-comparison analyses of ADHD and ASD latent factors** *(continued)*

#### Depression PGS · Howard et al. (2019)

| Outcome (trait, reporter) | N (offspring) | Families (clusters) | Population (β, 95% CI, SE, *p*, *q*) | Direct (β, 95% CI, SE, *p*, *q*) | Between (β, 95% CI, SE, *p*, *q*) | Indirect (β, 95% CI, SE, *p*, *q*) |
| --- | --- | --- | --- | --- | --- | --- |
| ADHD Common | 6,504 | 3,264 | 0.085 [0.059, 0.111] SE 0.013 *p* < .001 *q* < .001 | 0.051 [0.012, 0.090] SE 0.020 *p* = .011 *q* = .028 | 0.096 [0.065, 0.128] SE 0.016 *p* < .001 *q* < .001 | 0.034 [−0.004, 0.073] SE 0.019 *p* = .077 *q* = .386 |
| ADHD Parent | 6,452 | 3,227 | 0.079 [0.053, 0.104] SE 0.013 *p* < .001 *q* < .001 | 0.045 [0.006, 0.084] SE 0.020 *p* = .024 *q* = .052 | 0.089 [0.057, 0.120] SE 0.016 *p* < .001 *q* < .001 | 0.034 [−0.005, 0.072] SE 0.020 *p* = .087 *q* = .386 |
| ADHD Self | 4,639 | 2,567 | 0.086 [0.057, 0.116] SE 0.015 *p* < .001 *q* < .001 | 0.101 [0.045, 0.158] SE 0.029 *p* < .001 *q* = .002 | 0.083 [0.048, 0.117] SE 0.018 *p* < .001 *q* < .001 | −0.015 [−0.069, 0.037] SE 0.027 *p* = .583 *q* = .732 |
| ADHD Teacher | 4,590 | 2,362 | 0.047 [0.017, 0.078] SE 0.015 *p* = .002 *q* = .003 | 0.008 [−0.042, 0.058] SE 0.025 *p* = .752 *q* = .840 | 0.058 [0.023, 0.096] SE 0.019 *p* = .002 *q* = .003 | 0.039 [−0.008, 0.086] SE 0.024 *p* = .109 *q* = .386 |
| ASD Common | 6,188 | 3,122 | 0.071 [0.045, 0.097] SE 0.013 *p* < .001 *q* < .001 | 0.055 [0.017, 0.094] SE 0.019 *p* = .005 *q* = .014 | 0.076 [0.044, 0.108] SE 0.016 *p* < .001 *q* < .001 | 0.016 [−0.023, 0.056] SE 0.020 *p* = .418 *q* = .629 |
| ASD Parent | 5,940 | 2,970 | 0.066 [0.040, 0.093] SE 0.014 *p* < .001 *q* < .001 | 0.050 [0.011, 0.089] SE 0.020 *p* = .011 *q* = .028 | 0.072 [0.039, 0.105] SE 0.017 *p* < .001 *q* < .001 | 0.016 [−0.022, 0.057] SE 0.020 *p* = .409 *q* = .629 |
| ASD Self | 5,234 | 2,724 | 0.098 [0.068, 0.126] SE 0.015 *p* < .001 *q* < .001 | 0.071 [0.022, 0.117] SE 0.024 *p* = .004 *q* = .012 | 0.106 [0.070, 0.139] SE 0.018 *p* < .001 *q* < .001 | 0.027 [−0.017, 0.072] SE 0.023 *p* = .233 *q* = .446 |
| ASD Teacher | 4,583 | 2,413 | 0.015 [−0.015, 0.044] SE 0.015 *p* = .317 *q* = .344 | 0.044 [−0.006, 0.093] SE 0.025 *p* = .076 *q* = .152 | 0.006 [−0.029, 0.042] SE 0.018 *p* = .739 *q* = .763 | −0.030 [−0.078, 0.019] SE 0.024 *p* = .225 *q* = .446 |

**Notes:** Values are standardised regression coefficients (β), with 95% confidence intervals (CIs) in brackets, followed by standard errors (SE) and two-sided *p* values. N, number of offspring; Families, number of family clusters. Population, association of offspring PGS in the population model; Direct, within-family coefficient (child deviation from twin-pair mean PGS); Between, between-family coefficient (twin-pair mean PGS); Indirect, population minus within-family coefficient. SEs and percentile 95% CIs were obtained by bootstrapping with families as the resampling unit; *p* values were derived from z = β/SE using bootstrap SEs. False discovery rate (FDR)-adjusted *p* values (*q*; Benjamini–Hochberg) were computed separately for each estimate type across all PGS and outcomes in this table. Models adjust for sex, genotyping chip and the first ten genetic principal components.

**Supplementary Table S7. Twin-comparison analyses of ADHD and ASD latent factors** *(continued)*

#### Externalising PGS · Karlsson Linnér et al. (2021)

| Outcome (trait, reporter) | N (offspring) | Families (clusters) | Population (β, 95% CI, SE, *p*, *q*) | Direct (β, 95% CI, SE, *p*, *q*) | Between (β, 95% CI, SE, *p*, *q*) | Indirect (β, 95% CI, SE, *p*, *q*) |
| --- | --- | --- | --- | --- | --- | --- |
| ADHD Common | 6,504 | 3,264 | 0.154 [0.128, 0.179] SE 0.013 *p* < .001 *q* < .001 | 0.145 [0.107, 0.184] SE 0.020 *p* < .001 *q* < .001 | 0.156 [0.125, 0.188] SE 0.016 *p* < .001 *q* < .001 | 0.009 [−0.029, 0.046] SE 0.019 *p* = .656 *q* = .778 |
| ADHD Parent | 6,452 | 3,227 | 0.144 [0.119, 0.171] SE 0.013 *p* < .001 *q* < .001 | 0.138 [0.100, 0.175] SE 0.019 *p* < .001 *q* < .001 | 0.146 [0.116, 0.179] SE 0.016 *p* < .001 *q* < .001 | 0.006 [−0.031, 0.044] SE 0.019 *p* = .757 *q* = .850 |
| ADHD Self | 4,639 | 2,567 | 0.103 [0.073, 0.133] SE 0.015 *p* < .001 *q* < .001 | 0.143 [0.085, 0.202] SE 0.030 *p* < .001 *q* < .001 | 0.092 [0.057, 0.127] SE 0.018 *p* < .001 *q* < .001 | −0.041 [−0.093, 0.013] SE 0.028 *p* = .144 *q* = .415 |
| ADHD Teacher | 4,590 | 2,362 | 0.122 [0.091, 0.152] SE 0.016 *p* < .001 *q* < .001 | 0.089 [0.039, 0.141] SE 0.026 *p* < .001 *q* = .003 | 0.131 [0.094, 0.167] SE 0.019 *p* < .001 *q* < .001 | 0.033 [−0.018, 0.082] SE 0.026 *p* = .197 *q* = .446 |
| ASD Common | 6,188 | 3,122 | 0.048 [0.020, 0.075] SE 0.014 *p* < .001 *q* = .001 | −0.001 [−0.040, 0.038] SE 0.020 *p* = .942 *q* = .963 | 0.063 [0.029, 0.097] SE 0.017 *p* < .001 *q* < .001 | 0.049 [0.010, 0.089] SE 0.020 *p* = .015 *q* = .107 |
| ASD Parent | 5,940 | 2,970 | 0.047 [0.018, 0.074] SE 0.015 *p* = .002 *q* = .003 | −0.006 [−0.043, 0.033] SE 0.019 *p* = .774 *q* = .840 | 0.063 [0.028, 0.098] SE 0.018 *p* < .001 *q* < .001 | 0.052 [0.013, 0.093] SE 0.020 *p* = .010 *q* = .083 |
| ASD Self | 5,234 | 2,724 | 0.031 [0.003, 0.059] SE 0.014 *p* = .027 *q* = .032 | 0.041 [−0.012, 0.091] SE 0.026 *p* = .117 *q* = .221 | 0.028 [−0.005, 0.062] SE 0.017 *p* = .100 *q* = .120 | −0.010 [−0.055, 0.037] SE 0.023 *p* = .680 *q* = .778 |
| ASD Teacher | 4,583 | 2,413 | 0.023 [−0.009, 0.056] SE 0.017 *p* = .176 *q* = .198 | 0.001 [−0.049, 0.050] SE 0.026 *p* = .963 *q* = .963 | 0.029 [−0.010, 0.068] SE 0.020 *p* = .147 *q* = .171 | 0.021 [−0.027, 0.069] SE 0.025 *p* = .380 *q* = .629 |

**Notes:** Values are standardised regression coefficients (β), with 95% confidence intervals (CIs) in brackets, followed by standard errors (SE) and two-sided *p* values. N, number of offspring; Families, number of family clusters. Population, association of offspring PGS in the population model; Direct, within-family coefficient (child deviation from twin-pair mean PGS); Between, between-family coefficient (twin-pair mean PGS); Indirect, population minus within-family coefficient. SEs and percentile 95% CIs were obtained by bootstrapping with families as the resampling unit; *p* values were derived from z = β/SE using bootstrap SEs. False discovery rate (FDR)-adjusted *p* values (*q*; Benjamini–Hochberg) were computed separately for each estimate type across all PGS and outcomes in this table. Models adjust for sex, genotyping chip and the first ten genetic principal components.

**Supplementary Table S7. Twin-comparison analyses of ADHD and ASD latent factors** *(continued)*

#### Anxiety PGS · Purves et al. (2020)

| Outcome (trait, reporter) | N (offspring) | Families (clusters) | Population (β, 95% CI, SE, *p*, *q*) | Direct (β, 95% CI, SE, *p*, *q*) | Between (β, 95% CI, SE, *p*, *q*) | Indirect (β, 95% CI, SE, *p*, *q*) |
| --- | --- | --- | --- | --- | --- | --- |
| ADHD Common | 6,504 | 3,264 | −0.043 [−0.068, −0.017] SE 0.013 *p* < .001 *q* = .002 | −0.029 [−0.067, 0.008] SE 0.019 *p* = .131 *q* = .233 | −0.048 [−0.079, −0.016] SE 0.016 *p* = .003 *q* = .004 | −0.014 [−0.049, 0.021] SE 0.018 *p* = .437 *q* = .635 |
| ADHD Parent | 6,452 | 3,227 | −0.040 [−0.065, −0.014] SE 0.013 *p* = .002 *q* = .003 | −0.027 [−0.065, 0.010] SE 0.019 *p* = .159 *q* = .266 | −0.045 [−0.075, −0.013] SE 0.016 *p* = .005 *q* = .007 | −0.013 [−0.048, 0.023] SE 0.018 *p* = .489 *q* = .652 |
| ADHD Self | 4,639 | 2,567 | −0.049 [−0.078, −0.020] SE 0.015 *p* = .001 *q* = .002 | −0.042 [−0.097, 0.013] SE 0.028 *p* = .128 *q* = .233 | −0.051 [−0.084, −0.017] SE 0.017 *p* = .003 *q* = .004 | −0.006 [−0.055, 0.042] SE 0.025 *p* = .793 *q* = .876 |
| ADHD Teacher | 4,590 | 2,362 | −0.004 [−0.033, 0.025] SE 0.015 *p* = .790 *q* = .829 | 0.009 [−0.037, 0.055] SE 0.023 *p* = .688 *q* = .800 | −0.009 [−0.045, 0.027] SE 0.018 *p* = .617 *q* = .647 | −0.014 [−0.056, 0.030] SE 0.022 *p* = .529 *q* = .686 |
| ASD Common | 6,188 | 3,122 | −0.036 [−0.064, −0.009] SE 0.014 *p* = .010 *q* = .013 | −0.012 [−0.049, 0.024] SE 0.019 *p* = .515 *q* = .659 | −0.045 [−0.078, −0.010] SE 0.017 *p* = .008 *q* = .011 | −0.024 [−0.060, 0.013] SE 0.018 *p* = .195 *q* = .446 |
| ASD Parent | 5,940 | 2,970 | −0.033 [−0.059, −0.004] SE 0.014 *p* = .018 *q* = .023 | −0.011 [−0.047, 0.026] SE 0.019 *p* = .552 *q* = .676 | −0.041 [−0.075, −0.006] SE 0.018 *p* = .023 *q* = .029 | −0.022 [−0.058, 0.014] SE 0.018 *p* = .234 *q* = .446 |
| ASD Self | 5,234 | 2,724 | −0.051 [−0.079, −0.022] SE 0.015 *p* < .001 *q* = .001 | −0.034 [−0.081, 0.012] SE 0.024 *p* = .162 *q* = .266 | −0.056 [−0.093, −0.022] SE 0.018 *p* = .002 *q* = .003 | −0.017 [−0.061, 0.027] SE 0.023 *p* = .459 *q* = .642 |
| ASD Teacher | 4,583 | 2,413 | −0.022 [−0.054, 0.008] SE 0.016 *p* = .169 *q* = .193 | −0.006 [−0.054, 0.042] SE 0.024 *p* = .803 *q* = .857 | −0.028 [−0.067, 0.009] SE 0.020 *p* = .162 *q* = .185 | −0.016 [−0.061, 0.029] SE 0.023 *p* = .469 *q* = .642 |

**Notes:** Values are standardised regression coefficients (β), with 95% confidence intervals (CIs) in brackets, followed by standard errors (SE) and two-sided *p* values. N, number of offspring; Families, number of family clusters. Population, association of offspring PGS in the population model; Direct, within-family coefficient (child deviation from twin-pair mean PGS); Between, between-family coefficient (twin-pair mean PGS); Indirect, population minus within-family coefficient. SEs and percentile 95% CIs were obtained by bootstrapping with families as the resampling unit; *p* values were derived from z = β/SE using bootstrap SEs. False discovery rate (FDR)-adjusted *p* values (*q*; Benjamini–Hochberg) were computed separately for each estimate type across all PGS and outcomes in this table. Models adjust for sex, genotyping chip and the first ten genetic principal components.

**Supplementary Table S7. Twin-comparison analyses of ADHD and ASD latent factors** *(continued)*

#### Neuroticism PGS · Nagel et al. (2018)

| Outcome (trait, reporter) | N (offspring) | Families (clusters) | Population (β, 95% CI, SE, *p*, *q*) | Direct (β, 95% CI, SE, *p*, *q*) | Between (β, 95% CI, SE, *p*, *q*) | Indirect (β, 95% CI, SE, *p*, *q*) |
| --- | --- | --- | --- | --- | --- | --- |
| ADHD Common | 6,504 | 3,264 | 0.041 [0.016, 0.066] SE 0.013 *p* = .002 *q* = .003 | 0.006 [−0.032, 0.045] SE 0.020 *p* = .749 *q* = .840 | 0.053 [0.023, 0.084] SE 0.016 *p* < .001 *q* = .002 | 0.035 [−0.001, 0.073] SE 0.019 *p* = .067 *q* = .386 |
| ADHD Parent | 6,452 | 3,227 | 0.040 [0.015, 0.064] SE 0.013 *p* = .002 *q* = .003 | 0.006 [−0.031, 0.043] SE 0.019 *p* = .766 *q* = .840 | 0.052 [0.021, 0.082] SE 0.016 *p* = .001 *q* = .002 | 0.034 [−0.003, 0.072] SE 0.019 *p* = .070 *q* = .386 |
| ADHD Self | 4,639 | 2,567 | 0.039 [0.010, 0.067] SE 0.015 *p* = .009 *q* = .012 | 0.016 [−0.036, 0.070] SE 0.027 *p* = .552 *q* = .676 | 0.045 [0.011, 0.079] SE 0.017 *p* = .008 *q* = .011 | 0.022 [−0.029, 0.071] SE 0.025 *p* = .379 *q* = .629 |
| ADHD Teacher | 4,590 | 2,362 | 0.001 [−0.029, 0.030] SE 0.015 *p* = .947 *q* = .965 | −0.030 [−0.079, 0.019] SE 0.025 *p* = .229 *q* = .353 | 0.012 [−0.025, 0.047] SE 0.018 *p* = .505 *q* = .548 | 0.031 [−0.014, 0.074] SE 0.023 *p* = .173 *q* = .442 |
| ASD Common | 6,188 | 3,122 | 0.053 [0.027, 0.078] SE 0.013 *p* < .001 *q* < .001 | 0.022 [−0.016, 0.059] SE 0.019 *p* = .254 *q* = .369 | 0.064 [0.031, 0.096] SE 0.017 *p* < .001 *q* < .001 | 0.031 [−0.005, 0.069] SE 0.019 *p* = .101 *q* = .386 |
| ASD Parent | 5,940 | 2,970 | 0.055 [0.027, 0.081] SE 0.014 *p* < .001 *q* < .001 | 0.023 [−0.015, 0.061] SE 0.019 *p* = .232 *q* = .353 | 0.065 [0.030, 0.097] SE 0.017 *p* < .001 *q* < .001 | 0.032 [−0.008, 0.068] SE 0.019 *p* = .102 *q* = .386 |
| ASD Self | 5,234 | 2,724 | 0.047 [0.020, 0.076] SE 0.014 *p* < .001 *q* = .001 | 0.011 [−0.037, 0.060] SE 0.024 *p* = .645 *q* = .764 | 0.059 [0.026, 0.094] SE 0.017 *p* < .001 *q* < .001 | 0.036 [−0.009, 0.080] SE 0.023 *p* = .114 *q* = .386 |
| ASD Teacher | 4,583 | 2,413 | −0.004 [−0.034, 0.026] SE 0.015 *p* = .790 *q* = .829 | 0.001 [−0.045, 0.051] SE 0.025 *p* = .955 *q* = .963 | −0.006 [−0.042, 0.032] SE 0.019 *p* = .752 *q* = .764 | −0.006 [−0.054, 0.038] SE 0.024 *p* = .812 *q* = .880 |

**Notes:** Values are standardised regression coefficients (β), with 95% confidence intervals (CIs) in brackets, followed by standard errors (SE) and two-sided *p* values. N, number of offspring; Families, number of family clusters. Population, association of offspring PGS in the population model; Direct, within-family coefficient (child deviation from twin-pair mean PGS); Between, between-family coefficient (twin-pair mean PGS); Indirect, population minus within-family coefficient. SEs and percentile 95% CIs were obtained by bootstrapping with families as the resampling unit; *p* values were derived from z = β/SE using bootstrap SEs. False discovery rate (FDR)-adjusted *p* values (*q*; Benjamini–Hochberg) were computed separately for each estimate type across all PGS and outcomes in this table. Models adjust for sex, genotyping chip and the first ten genetic principal components.

**Supplementary Table S7. Twin-comparison analyses of ADHD and ASD latent factors** *(continued)*

#### Cognitive PGS · Demange et al. (2021)

| Outcome (trait, reporter) | N (offspring) | Families (clusters) | Population (β, 95% CI, SE, *p*, *q*) | Direct (β, 95% CI, SE, *p*, *q*) | Between (β, 95% CI, SE, *p*, *q*) | Indirect (β, 95% CI, SE, *p*, *q*) |
| --- | --- | --- | --- | --- | --- | --- |
| ADHD Common | 6,504 | 3,264 | −0.113 [−0.139, −0.087] SE 0.013 *p* < .001 *q* < .001 | −0.114 [−0.152, −0.076] SE 0.019 *p* < .001 *q* < .001 | −0.113 [−0.145, −0.081] SE 0.016 *p* < .001 *q* < .001 | 0.001 [−0.035, 0.040] SE 0.019 *p* = .965 *q* = .980 |
| ADHD Parent | 6,452 | 3,227 | −0.111 [−0.137, −0.086] SE 0.013 *p* < .001 *q* < .001 | −0.116 [−0.153, −0.078] SE 0.019 *p* < .001 *q* < .001 | −0.110 [−0.141, −0.078] SE 0.016 *p* < .001 *q* < .001 | 0.004 [−0.032, 0.042] SE 0.019 *p* = .825 *q* = .880 |
| ADHD Self | 4,639 | 2,567 | −0.001 [−0.031, 0.031] SE 0.016 *p* = .950 *q* = .965 | −0.019 [−0.077, 0.039] SE 0.030 *p* = .510 *q* = .659 | 0.005 [−0.029, 0.041] SE 0.018 *p* = .781 *q* = .781 | 0.019 [−0.032, 0.070] SE 0.026 *p* = .472 *q* = .642 |
| ADHD Teacher | 4,590 | 2,362 | −0.114 [−0.143, −0.085] SE 0.015 *p* < .001 *q* < .001 | −0.087 [−0.133, −0.041] SE 0.024 *p* < .001 *q* < .001 | −0.122 [−0.158, −0.086] SE 0.018 *p* < .001 *q* < .001 | −0.027 [−0.071, 0.018] SE 0.023 *p* = .237 *q* = .446 |
| ASD Common | 6,188 | 3,122 | −0.051 [−0.078, −0.024] SE 0.014 *p* < .001 *q* < .001 | 0.020 [−0.022, 0.062] SE 0.021 *p* = .350 *q* = .498 | −0.074 [−0.108, −0.041] SE 0.017 *p* < .001 *q* < .001 | −0.071 [−0.110, −0.031] SE 0.021 *p* < .001 *q* = .006 |
| ASD Parent | 5,940 | 2,970 | −0.062 [−0.090, −0.036] SE 0.014 *p* < .001 *q* < .001 | 0.012 [−0.030, 0.054] SE 0.021 *p* = .560 *q* = .676 | −0.087 [−0.121, −0.054] SE 0.017 *p* < .001 *q* < .001 | −0.075 [−0.115, −0.034] SE 0.021 *p* < .001 *q* = .005 |
| ASD Self | 5,234 | 2,724 | 0.025 [−0.003, 0.053] SE 0.014 *p* = .074 *q* = .088 | 0.052 [0.004, 0.098] SE 0.024 *p* = .031 *q* = .066 | 0.017 [−0.018, 0.050] SE 0.017 *p* = .317 *q* = .356 | −0.027 [−0.070, 0.017] SE 0.022 *p* = .228 *q* = .446 |
| ASD Teacher | 4,583 | 2,413 | 0.019 [−0.011, 0.049] SE 0.015 *p* = .205 *q* = .227 | 0.029 [−0.021, 0.077] SE 0.025 *p* = .247 *q* = .368 | 0.016 [−0.021, 0.053] SE 0.019 *p* = .400 *q* = .441 | −0.010 [−0.057, 0.037] SE 0.024 *p* = .681 *q* = .778 |

**Notes:** Values are standardised regression coefficients (β), with 95% confidence intervals (CIs) in brackets, followed by standard errors (SE) and two-sided *p* values. N, number of offspring; Families, number of family clusters. Population, association of offspring PGS in the population model; Direct, within-family coefficient (child deviation from twin-pair mean PGS); Between, between-family coefficient (twin-pair mean PGS); Indirect, population minus within-family coefficient. SEs and percentile 95% CIs were obtained by bootstrapping with families as the resampling unit; *p* values were derived from z = β/SE using bootstrap SEs. False discovery rate (FDR)-adjusted *p* values (*q*; Benjamini–Hochberg) were computed separately for each estimate type across all PGS and outcomes in this table. Models adjust for sex, genotyping chip and the first ten genetic principal components.

**Supplementary Table S7. Twin-comparison analyses of ADHD and ASD latent factors** *(continued)*

#### Educational attainment PGS · Okbay et al. (2022)

| Outcome (trait, reporter) | N (offspring) | Families (clusters) | Population (β, 95% CI, SE, *p*, *q*) | Direct (β, 95% CI, SE, *p*, *q*) | Between (β, 95% CI, SE, *p*, *q*) | Indirect (β, 95% CI, SE, *p*, *q*) |
| --- | --- | --- | --- | --- | --- | --- |
| ADHD Common | 6,504 | 3,264 | −0.149 [−0.174, −0.123] SE 0.013 *p* < .001 *q* < .001 | −0.127 [−0.169, −0.085] SE 0.022 *p* < .001 *q* < .001 | −0.154 [−0.184, −0.124] SE 0.015 *p* < .001 *q* < .001 | −0.021 [−0.061, 0.018] SE 0.021 *p* = .301 *q* = .551 |
| ADHD Parent | 6,452 | 3,227 | −0.143 [−0.170, −0.117] SE 0.013 *p* < .001 *q* < .001 | −0.126 [−0.169, −0.083] SE 0.022 *p* < .001 *q* < .001 | −0.148 [−0.178, −0.117] SE 0.016 *p* < .001 *q* < .001 | −0.018 [−0.060, 0.024] SE 0.021 *p* = .403 *q* = .629 |
| ADHD Self | 4,639 | 2,567 | −0.038 [−0.069, −0.006] SE 0.016 *p* = .018 *q* = .022 | −0.077 [−0.143, −0.013] SE 0.033 *p* = .020 *q* = .046 | −0.028 [−0.064, 0.006] SE 0.018 *p* = .120 *q* = .142 | 0.040 [−0.020, 0.098] SE 0.031 *p* = .194 *q* = .446 |
| ADHD Teacher | 4,590 | 2,362 | −0.142 [−0.171, −0.113] SE 0.015 *p* < .001 *q* < .001 | −0.111 [−0.162, −0.058] SE 0.026 *p* < .001 *q* < .001 | −0.150 [−0.184, −0.116] SE 0.017 *p* < .001 *q* < .001 | −0.031 [−0.081, 0.018] SE 0.025 *p* = .207 *q* = .446 |
| ASD Common | 6,188 | 3,122 | −0.095 [−0.123, −0.068] SE 0.014 *p* < .001 *q* < .001 | 0.065 [0.021, 0.107] SE 0.022 *p* = .004 *q* = .012 | −0.135 [−0.168, −0.102] SE 0.017 *p* < .001 *q* < .001 | −0.160 [−0.204, −0.116] SE 0.022 *p* < .001 *q* < .001 |
| ASD Parent | 5,940 | 2,970 | −0.102 [−0.130, −0.075] SE 0.014 *p* < .001 *q* < .001 | 0.059 [0.017, 0.103] SE 0.022 *p* = .007 *q* = .018 | −0.144 [−0.176, −0.111] SE 0.017 *p* < .001 *q* < .001 | −0.162 [−0.204, −0.119] SE 0.022 *p* < .001 *q* < .001 |
| ASD Self | 5,234 | 2,724 | −0.024 [−0.053, 0.004] SE 0.015 *p* = .110 *q* = .128 | 0.044 [−0.010, 0.097] SE 0.027 *p* = .108 *q* = .209 | −0.040 [−0.075, −0.007] SE 0.017 *p* = .019 *q* = .024 | −0.068 [−0.119, −0.017] SE 0.026 *p* = .010 *q* = .083 |
| ASD Teacher | 4,583 | 2,413 | 0.000 [−0.034, 0.032] SE 0.017 *p* = 1.000 *q* = 1.000 | 0.041 [−0.012, 0.097] SE 0.028 *p* = .142 *q* = .245 | −0.010 [−0.050, 0.028] SE 0.020 *p* = .617 *q* = .647 | −0.042 [−0.097, 0.012] SE 0.028 *p* = .134 *q* = .415 |

**Notes:** Values are standardised regression coefficients (β), with 95% confidence intervals (CIs) in brackets, followed by standard errors (SE) and two-sided *p* values. N, number of offspring; Families, number of family clusters. Population, association of offspring PGS in the population model; Direct, within-family coefficient (child deviation from twin-pair mean PGS); Between, between-family coefficient (twin-pair mean PGS); Indirect, population minus within-family coefficient. SEs and percentile 95% CIs were obtained by bootstrapping with families as the resampling unit; *p* values were derived from z = β/SE using bootstrap SEs. False discovery rate (FDR)-adjusted *p* values (*q*; Benjamini–Hochberg) were computed separately for each estimate type across all PGS and outcomes in this table. Models adjust for sex, genotyping chip and the first ten genetic principal components.

### Supplementary Table S8. Trio-PGS analyses of ADHD and ASD latent factors

#### ADHD PGS · Demontis et al. (2023)

| Outcome (trait, reporter) | N (offspring) | Families (clusters) | Child PGS (β, 95% CI, SE, *p*, *q*) | Maternal PGS (β, 95% CI, SE, *p*, *q*) | Paternal PGS (β, 95% CI, SE, *p*, *q*) |
| --- | --- | --- | --- | --- | --- |
| ADHD Common | 7,259 | 4,025 | 0.145 [0.113, 0.177] SE 0.016 *p* < .001 *q* < .001 | 0.021 [−0.008, 0.049] SE 0.014 *p* = .154 *q* = .379 | 0.019 [−0.012, 0.049] SE 0.015 *p* = .228 *q* = .486 |
| ADHD Parent | 6,440 | 3,221 | 0.141 [0.108, 0.174] SE 0.017 *p* < .001 *q* < .001 | 0.026 [−0.006, 0.058] SE 0.016 *p* = .106 *q* = .285 | 0.005 [−0.028, 0.038] SE 0.017 *p* = .763 *q* = .917 |
| ADHD Self | 4,631 | 2,563 | 0.068 [0.025, 0.111] SE 0.022 *p* = .002 *q* = .005 | 0.022 [−0.013, 0.057] SE 0.018 *p* = .214 *q* = .472 | 0.011 [−0.025, 0.046] SE 0.018 *p* = .548 *q* = .818 |
| ADHD Teacher | 4,580 | 2,357 | 0.156 [0.115, 0.196] SE 0.021 *p* < .001 *q* < .001 | −0.005 [−0.041, 0.031] SE 0.018 *p* = .777 *q* = .975 | 0.035 [−0.004, 0.074] SE 0.020 *p* = .077 *q* = .329 |
| ASD Common | 6,934 | 3,874 | 0.052 [0.019, 0.086] SE 0.017 *p* = .002 *q* = .005 | 0.054 [0.025, 0.084] SE 0.015 *p* < .001 *q* = .003 | 0.024 [−0.008, 0.056] SE 0.016 *p* = .146 *q* = .425 |
| ASD Parent | 5,930 | 2,965 | 0.042 [0.006, 0.078] SE 0.018 *p* = .023 *q* = .059 | 0.062 [0.028, 0.096] SE 0.017 *p* < .001 *q* = .003 | 0.016 [−0.021, 0.052] SE 0.018 *p* = .394 *q* = .700 |
| ASD Self | 5,224 | 2,719 | 0.029 [−0.012, 0.071] SE 0.021 *p* = .166 *q* = .280 | 0.047 [0.012, 0.081] SE 0.018 *p* = .009 *q* = .040 | 0.008 [−0.030, 0.045] SE 0.019 *p* = .688 *q* = .885 |
| ASD Teacher | 4,575 | 2,409 | 0.078 [0.035, 0.121] SE 0.022 *p* < .001 *q* = .001 | −0.012 [−0.047, 0.024] SE 0.018 *p* = .527 *q* = .823 | −0.001 [−0.040, 0.037] SE 0.020 *p* = .948 *q* = .970 |

**Notes:** Values are standardised regression coefficients (β), with 95% confidence intervals (CIs) in brackets, followed by standard errors (SE) and two-sided *p* values. N, number of offspring; Families, number of family clusters. Child, maternal and paternal PGS were entered simultaneously; child PGS coefficients index direct genetic effects and parental PGS coefficients index indirect genetic effects. SEs are family-clustered robust estimates; 95% CIs and *p* values use family-cluster degrees of freedom. False discovery rate (FDR)-adjusted *p* values (*q*; Benjamini–Hochberg) were computed separately for each estimate type across all PGS and outcomes in this table. Models adjust for sex, genotyping chip and the first ten genetic principal components.

**Supplementary Table S8. Trio-PGS analyses of ADHD and ASD latent factors** *(continued)*

#### ASD PGS · Grove et al. (2019)

| Outcome (trait, reporter) | N (offspring) | Families (clusters) | Child PGS (β, 95% CI, SE, *p*, *q*) | Maternal PGS (β, 95% CI, SE, *p*, *q*) | Paternal PGS (β, 95% CI, SE, *p*, *q*) |
| --- | --- | --- | --- | --- | --- |
| ADHD Common | 7,259 | 4,025 | 0.061 [0.029, 0.092] SE 0.016 *p* < .001 *q* < .001 | 0.000 [−0.029, 0.029] SE 0.015 *p* = .998 *q* = .998 | −0.020 [−0.050, 0.010] SE 0.015 *p* = .194 *q* = .467 |
| ADHD Parent | 6,440 | 3,221 | 0.060 [0.026, 0.094] SE 0.017 *p* < .001 *q* = .002 | 0.013 [−0.019, 0.045] SE 0.016 *p* = .427 *q* = .757 | −0.030 [−0.064, 0.003] SE 0.017 *p* = .077 *q* = .329 |
| ADHD Self | 4,631 | 2,563 | 0.025 [−0.017, 0.068] SE 0.022 *p* = .236 *q* = .336 | 0.029 [−0.006, 0.064] SE 0.018 *p* = .107 *q* = .285 | 0.005 [−0.033, 0.042] SE 0.019 *p* = .812 *q* = .917 |
| ADHD Teacher | 4,580 | 2,357 | 0.021 [−0.020, 0.062] SE 0.021 *p* = .319 *q* = .425 | −0.001 [−0.039, 0.036] SE 0.019 *p* = .944 *q* = .978 | 0.025 [−0.013, 0.064] SE 0.020 *p* = .193 *q* = .467 |
| ASD Common | 6,934 | 3,874 | 0.102 [0.068, 0.136] SE 0.017 *p* < .001 *q* < .001 | −0.005 [−0.035, 0.024] SE 0.015 *p* = .722 *q* = .943 | −0.039 [−0.070, −0.008] SE 0.016 *p* = .013 *q* = .166 |
| ASD Parent | 5,930 | 2,965 | 0.091 [0.054, 0.128] SE 0.019 *p* < .001 *q* < .001 | 0.002 [−0.033, 0.036] SE 0.018 *p* = .930 *q* = .978 | −0.034 [−0.069, 0.000] SE 0.018 *p* = .053 *q* = .299 |
| ASD Self | 5,224 | 2,719 | 0.078 [0.039, 0.116] SE 0.020 *p* < .001 *q* < .001 | 0.002 [−0.035, 0.040] SE 0.019 *p* = .900 *q* = .978 | −0.017 [−0.054, 0.020] SE 0.019 *p* = .366 *q* = .689 |
| ASD Teacher | 4,575 | 2,409 | 0.069 [0.030, 0.108] SE 0.020 *p* < .001 *q* = .002 | −0.023 [−0.061, 0.014] SE 0.019 *p* = .224 *q* = .478 | −0.011 [−0.048, 0.026] SE 0.019 *p* = .570 *q* = .818 |

**Notes:** Values are standardised regression coefficients (β), with 95% confidence intervals (CIs) in brackets, followed by standard errors (SE) and two-sided *p* values. N, number of offspring; Families, number of family clusters. Child, maternal and paternal PGS were entered simultaneously; child PGS coefficients index direct genetic effects and parental PGS coefficients index indirect genetic effects. SEs are family-clustered robust estimates; 95% CIs and *p* values use family-cluster degrees of freedom. False discovery rate (FDR)-adjusted *p* values (*q*; Benjamini–Hochberg) were computed separately for each estimate type across all PGS and outcomes in this table. Models adjust for sex, genotyping chip and the first ten genetic principal components.

**Supplementary Table S8. Trio-PGS analyses of ADHD and ASD latent factors** *(continued)*

#### Depression PGS · Howard et al. (2019)

| Outcome (trait, reporter) | N (offspring) | Families (clusters) | Child PGS (β, 95% CI, SE, *p*, *q*) | Maternal PGS (β, 95% CI, SE, *p*, *q*) | Paternal PGS (β, 95% CI, SE, *p*, *q*) |
| --- | --- | --- | --- | --- | --- |
| ADHD Common | 7,259 | 4,025 | 0.038 [0.004, 0.071] SE 0.017 *p* = .030 *q* = .071 | 0.046 [0.017, 0.074] SE 0.015 *p* = .002 *q* = .010 | 0.023 [−0.009, 0.054] SE 0.016 *p* = .160 *q* = .445 |
| ADHD Parent | 6,440 | 3,221 | 0.039 [0.003, 0.075] SE 0.018 *p* = .034 *q* = .078 | 0.050 [0.019, 0.082] SE 0.016 *p* = .002 *q* = .010 | 0.014 [−0.021, 0.048] SE 0.017 *p* = .436 *q* = .715 |
| ADHD Self | 4,631 | 2,563 | 0.079 [0.037, 0.121] SE 0.022 *p* < .001 *q* < .001 | 0.010 [−0.025, 0.046] SE 0.018 *p* = .561 *q* = .825 | 0.004 [−0.033, 0.040] SE 0.019 *p* = .850 *q* = .924 |
| ADHD Teacher | 4,580 | 2,357 | 0.020 [−0.022, 0.061] SE 0.021 *p* = .352 *q* = .460 | 0.013 [−0.026, 0.051] SE 0.020 *p* = .521 *q* = .823 | 0.030 [−0.010, 0.070] SE 0.020 *p* = .137 *q* = .418 |
| ASD Common | 6,934 | 3,874 | 0.032 [−0.002, 0.066] SE 0.017 *p* = .064 *q* = .132 | 0.061 [0.029, 0.092] SE 0.016 *p* < .001 *q* = .002 | 0.006 [−0.026, 0.039] SE 0.017 *p* = .706 *q* = .885 |
| ASD Parent | 5,930 | 2,965 | 0.029 [−0.008, 0.065] SE 0.019 *p* = .123 *q* = .219 | 0.068 [0.033, 0.103] SE 0.018 *p* < .001 *q* = .002 | −0.004 [−0.040, 0.032] SE 0.018 *p* = .813 *q* = .917 |
| ASD Self | 5,224 | 2,719 | 0.067 [0.027, 0.107] SE 0.020 *p* < .001 *q* = .003 | 0.034 [−0.003, 0.070] SE 0.019 *p* = .074 *q* = .237 | 0.018 [−0.022, 0.057] SE 0.020 *p* = .382 *q* = .699 |
| ASD Teacher | 4,575 | 2,409 | 0.018 [−0.024, 0.059] SE 0.021 *p* = .406 *q* = .520 | −0.011 [−0.051, 0.028] SE 0.020 *p* = .567 *q* = .825 | 0.007 [−0.032, 0.046] SE 0.020 *p* = .719 *q* = .885 |

**Notes:** Values are standardised regression coefficients (β), with 95% confidence intervals (CIs) in brackets, followed by standard errors (SE) and two-sided *p* values. N, number of offspring; Families, number of family clusters. Child, maternal and paternal PGS were entered simultaneously; child PGS coefficients index direct genetic effects and parental PGS coefficients index indirect genetic effects. SEs are family-clustered robust estimates; 95% CIs and *p* values use family-cluster degrees of freedom. False discovery rate (FDR)-adjusted *p* values (*q*; Benjamini–Hochberg) were computed separately for each estimate type across all PGS and outcomes in this table. Models adjust for sex, genotyping chip and the first ten genetic principal components.

**Supplementary Table S8. Trio-PGS analyses of ADHD and ASD latent factors** *(continued)*

#### Externalising PGS · Karlsson Linnér et al. (2021)

| Outcome (trait, reporter) | N (offspring) | Families (clusters) | Child PGS (β, 95% CI, SE, *p*, *q*) | Maternal PGS (β, 95% CI, SE, *p*, *q*) | Paternal PGS (β, 95% CI, SE, *p*, *q*) |
| --- | --- | --- | --- | --- | --- |
| ADHD Common | 7,259 | 4,025 | 0.142 [0.109, 0.174] SE 0.017 *p* < .001 *q* < .001 | 0.003 [−0.026, 0.031] SE 0.015 *p* = .860 *q* = .978 | 0.006 [−0.024, 0.036] SE 0.015 *p* = .695 *q* = .885 |
| ADHD Parent | 6,440 | 3,221 | 0.142 [0.107, 0.177] SE 0.018 *p* < .001 *q* < .001 | 0.000 [−0.032, 0.032] SE 0.016 *p* = .994 *q* = .998 | 0.003 [−0.030, 0.036] SE 0.017 *p* = .866 *q* = .924 |
| ADHD Self | 4,631 | 2,563 | 0.104 [0.061, 0.147] SE 0.022 *p* < .001 *q* < .001 | −0.005 [−0.040, 0.031] SE 0.018 *p* = .804 *q* = .978 | 0.004 [−0.034, 0.041] SE 0.019 *p* = .852 *q* = .924 |
| ADHD Teacher | 4,580 | 2,357 | 0.081 [0.039, 0.124] SE 0.022 *p* < .001 *q* < .001 | 0.009 [−0.029, 0.048] SE 0.020 *p* = .628 *q* = .837 | 0.052 [0.013, 0.091] SE 0.020 *p* = .008 *q* = .128 |
| ASD Common | 6,934 | 3,874 | 0.026 [−0.008, 0.059] SE 0.017 *p* = .132 *q* = .228 | 0.035 [0.004, 0.065] SE 0.015 *p* = .024 *q* = .081 | −0.007 [−0.039, 0.026] SE 0.017 *p* = .691 *q* = .885 |
| ASD Parent | 5,930 | 2,965 | 0.029 [−0.008, 0.065] SE 0.019 *p* = .123 *q* = .219 | 0.044 [0.009, 0.078] SE 0.018 *p* = .013 *q* = .049 | −0.013 [−0.050, 0.023] SE 0.018 *p* = .465 *q* = .744 |
| ASD Self | 5,224 | 2,719 | 0.006 [−0.036, 0.048] SE 0.022 *p* = .781 *q* = .877 | 0.016 [−0.021, 0.053] SE 0.019 *p* = .400 *q* = .753 | 0.025 [−0.015, 0.066] SE 0.021 *p* = .216 *q* = .477 |
| ASD Teacher | 4,575 | 2,409 | 0.001 [−0.045, 0.046] SE 0.023 *p* = .982 *q* = .982 | −0.004 [−0.042, 0.034] SE 0.019 *p* = .837 *q* = .978 | 0.037 [−0.002, 0.077] SE 0.020 *p* = .063 *q* = .310 |

**Notes:** Values are standardised regression coefficients (β), with 95% confidence intervals (CIs) in brackets, followed by standard errors (SE) and two-sided *p* values. N, number of offspring; Families, number of family clusters. Child, maternal and paternal PGS were entered simultaneously; child PGS coefficients index direct genetic effects and parental PGS coefficients index indirect genetic effects. SEs are family-clustered robust estimates; 95% CIs and *p* values use family-cluster degrees of freedom. False discovery rate (FDR)-adjusted *p* values (*q*; Benjamini–Hochberg) were computed separately for each estimate type across all PGS and outcomes in this table. Models adjust for sex, genotyping chip and the first ten genetic principal components.

**Supplementary Table S8. Trio-PGS analyses of ADHD and ASD latent factors** *(continued)*

#### Anxiety PGS · Purves et al. (2020)

| Outcome (trait, reporter) | N (offspring) | Families (clusters) | Child PGS (β, 95% CI, SE, *p*, *q*) | Maternal PGS (β, 95% CI, SE, *p*, *q*) | Paternal PGS (β, 95% CI, SE, *p*, *q*) |
| --- | --- | --- | --- | --- | --- |
| ADHD Common | 7,259 | 4,025 | −0.023 [−0.056, 0.010] SE 0.017 *p* = .173 *q* = .283 | −0.017 [−0.046, 0.013] SE 0.015 *p* = .269 *q* = .555 | −0.020 [−0.050, 0.009] SE 0.015 *p* = .177 *q* = .467 |
| ADHD Parent | 6,440 | 3,221 | −0.014 [−0.049, 0.021] SE 0.018 *p* = .429 *q* = .528 | −0.021 [−0.054, 0.012] SE 0.017 *p* = .214 *q* = .472 | −0.021 [−0.053, 0.012] SE 0.016 *p* = .211 *q* = .477 |
| ADHD Self | 4,631 | 2,563 | −0.042 [−0.082, −0.001] SE 0.021 *p* = .042 *q* = .093 | 0.005 [−0.029, 0.040] SE 0.018 *p* = .769 *q* = .975 | −0.017 [−0.052, 0.018] SE 0.018 *p* = .354 *q* = .687 |
| ADHD Teacher | 4,580 | 2,357 | 0.016 [−0.023, 0.054] SE 0.020 *p* = .428 *q* = .528 | −0.002 [−0.041, 0.037] SE 0.020 *p* = .920 *q* = .978 | −0.031 [−0.068, 0.007] SE 0.019 *p* = .106 *q* = .406 |
| ASD Common | 6,934 | 3,874 | 0.009 [−0.024, 0.042] SE 0.017 *p* = .581 *q* = .689 | −0.055 [−0.085, −0.024] SE 0.016 *p* < .001 *q* = .004 | −0.034 [−0.065, −0.004] SE 0.016 *p* = .029 *q* = .186 |
| ASD Parent | 5,930 | 2,965 | 0.025 [−0.011, 0.060] SE 0.018 *p* = .177 *q* = .283 | −0.055 [−0.090, −0.020] SE 0.018 *p* = .002 *q* = .010 | −0.040 [−0.074, −0.006] SE 0.017 *p* = .021 *q* = .184 |
| ASD Self | 5,224 | 2,719 | −0.027 [−0.066, 0.013] SE 0.020 *p* = .192 *q* = .300 | −0.009 [−0.045, 0.027] SE 0.018 *p* = .618 *q* = .837 | −0.030 [−0.068, 0.008] SE 0.019 *p* = .121 *q* = .406 |
| ASD Teacher | 4,575 | 2,409 | −0.008 [−0.049, 0.033] SE 0.021 *p* = .717 *q* = .819 | 0.001 [−0.038, 0.041] SE 0.020 *p* = .947 *q* = .978 | −0.025 [−0.063, 0.013] SE 0.019 *p* = .197 *q* = .467 |

**Notes:** Values are standardised regression coefficients (β), with 95% confidence intervals (CIs) in brackets, followed by standard errors (SE) and two-sided *p* values. N, number of offspring; Families, number of family clusters. Child, maternal and paternal PGS were entered simultaneously; child PGS coefficients index direct genetic effects and parental PGS coefficients index indirect genetic effects. SEs are family-clustered robust estimates; 95% CIs and *p* values use family-cluster degrees of freedom. False discovery rate (FDR)-adjusted *p* values (*q*; Benjamini–Hochberg) were computed separately for each estimate type across all PGS and outcomes in this table. Models adjust for sex, genotyping chip and the first ten genetic principal components.

**Supplementary Table S8. Trio-PGS analyses of ADHD and ASD latent factors** *(continued)*

#### Neuroticism PGS · Nagel et al. (2018)

| Outcome (trait, reporter) | N (offspring) | Families (clusters) | Child PGS (β, 95% CI, SE, *p*, *q*) | Maternal PGS (β, 95% CI, SE, *p*, *q*) | Paternal PGS (β, 95% CI, SE, *p*, *q*) |
| --- | --- | --- | --- | --- | --- |
| ADHD Common | 7,259 | 4,025 | 0.021 [−0.011, 0.053] SE 0.016 *p* = .206 *q* = .313 | 0.045 [0.016, 0.075] SE 0.015 *p* = .002 *q* = .010 | −0.006 [−0.036, 0.023] SE 0.015 *p* = .676 *q* = .885 |
| ADHD Parent | 6,440 | 3,221 | 0.020 [−0.014, 0.054] SE 0.017 *p* = .245 *q* = .341 | 0.043 [0.010, 0.075] SE 0.017 *p* = .010 *q* = .040 | −0.010 [−0.043, 0.023] SE 0.017 *p* = .561 *q* = .818 |
| ADHD Self | 4,631 | 2,563 | 0.022 [−0.017, 0.062] SE 0.020 *p* = .273 *q* = .372 | 0.029 [−0.005, 0.063] SE 0.017 *p* = .093 *q* = .271 | −0.001 [−0.038, 0.036] SE 0.019 *p* = .970 *q* = .970 |
| ADHD Teacher | 4,580 | 2,357 | −0.027 [−0.069, 0.015] SE 0.021 *p* = .210 *q* = .313 | 0.015 [−0.024, 0.054] SE 0.020 *p* = .444 *q* = .757 | 0.030 [−0.007, 0.068] SE 0.019 *p* = .116 *q* = .406 |
| ASD Common | 6,934 | 3,874 | 0.026 [−0.007, 0.058] SE 0.016 *p* = .119 *q* = .219 | 0.074 [0.043, 0.105] SE 0.016 *p* < .001 *q* < .001 | −0.013 [−0.044, 0.018] SE 0.016 *p* = .418 *q* = .715 |
| ASD Parent | 5,930 | 2,965 | 0.021 [−0.014, 0.056] SE 0.018 *p* = .234 *q* = .336 | 0.077 [0.042, 0.112] SE 0.018 *p* < .001 *q* < .001 | −0.021 [−0.056, 0.014] SE 0.018 *p* = .240 *q* = .495 |
| ASD Self | 5,224 | 2,719 | 0.038 [−0.002, 0.078] SE 0.020 *p* = .062 *q* = .132 | 0.020 [−0.018, 0.057] SE 0.019 *p* = .305 *q* = .592 | −0.005 [−0.043, 0.034] SE 0.020 *p* = .816 *q* = .917 |
| ASD Teacher | 4,575 | 2,409 | −0.002 [−0.043, 0.040] SE 0.021 *p* = .936 *q* = .982 | 0.015 [−0.024, 0.054] SE 0.020 *p* = .457 *q* = .757 | −0.017 [−0.053, 0.019] SE 0.018 *p* = .354 *q* = .687 |

**Notes:** Values are standardised regression coefficients (β), with 95% confidence intervals (CIs) in brackets, followed by standard errors (SE) and two-sided *p* values. N, number of offspring; Families, number of family clusters. Child, maternal and paternal PGS were entered simultaneously; child PGS coefficients index direct genetic effects and parental PGS coefficients index indirect genetic effects. SEs are family-clustered robust estimates; 95% CIs and *p* values use family-cluster degrees of freedom. False discovery rate (FDR)-adjusted *p* values (*q*; Benjamini–Hochberg) were computed separately for each estimate type across all PGS and outcomes in this table. Models adjust for sex, genotyping chip and the first ten genetic principal components.

**Supplementary Table S8. Trio-PGS analyses of ADHD and ASD latent factors** *(continued)*

#### Cognitive PGS · Demange et al. (2021)

| Outcome (trait, reporter) | N (offspring) | Families (clusters) | Child PGS (β, 95% CI, SE, *p*, *q*) | Maternal PGS (β, 95% CI, SE, *p*, *q*) | Paternal PGS (β, 95% CI, SE, *p*, *q*) |
| --- | --- | --- | --- | --- | --- |
| ADHD Common | 7,259 | 4,025 | −0.096 [−0.128, −0.064] SE 0.016 *p* < .001 *q* < .001 | 0.003 [−0.026, 0.031] SE 0.015 *p* = .852 *q* = .978 | −0.024 [−0.054, 0.005] SE 0.015 *p* = .109 *q* = .406 |
| ADHD Parent | 6,440 | 3,221 | −0.100 [−0.134, −0.066] SE 0.017 *p* < .001 *q* < .001 | 0.009 [−0.024, 0.041] SE 0.016 *p* = .605 *q* = .837 | −0.025 [−0.058, 0.007] SE 0.017 *p* = .127 *q* = .406 |
| ADHD Self | 4,631 | 2,563 | −0.008 [−0.049, 0.034] SE 0.021 *p* = .716 *q* = .819 | 0.013 [−0.021, 0.047] SE 0.017 *p* = .441 *q* = .757 | −0.001 [−0.037, 0.035] SE 0.018 *p* = .949 *q* = .970 |
| ADHD Teacher | 4,580 | 2,357 | −0.106 [−0.145, −0.067] SE 0.020 *p* < .001 *q* < .001 | −0.013 [−0.049, 0.022] SE 0.018 *p* = .461 *q* = .757 | 0.001 [−0.036, 0.038] SE 0.019 *p* = .958 *q* = .970 |
| ASD Common | 6,934 | 3,874 | 0.002 [−0.032, 0.037] SE 0.017 *p* = .889 *q* = .981 | −0.040 [−0.071, −0.010] SE 0.016 *p* = .010 *q* = .040 | −0.044 [−0.074, −0.013] SE 0.016 *p* = .005 *q* = .107 |
| ASD Parent | 5,930 | 2,965 | −0.013 [−0.050, 0.023] SE 0.019 *p* = .475 *q* = .574 | −0.041 [−0.076, −0.006] SE 0.018 *p* = .022 *q* = .078 | −0.039 [−0.074, −0.005] SE 0.018 *p* = .026 *q* = .185 |
| ASD Self | 5,224 | 2,719 | 0.033 [−0.006, 0.073] SE 0.020 *p* = .099 *q* = .196 | −0.010 [−0.046, 0.026] SE 0.018 *p* = .596 *q* = .837 | −0.005 [−0.044, 0.035] SE 0.020 *p* = .817 *q* = .917 |
| ASD Teacher | 4,575 | 2,409 | 0.033 [−0.006, 0.071] SE 0.020 *p* = .101 *q* = .196 | −0.011 [−0.047, 0.025] SE 0.018 *p* = .545 *q* = .825 | −0.010 [−0.047, 0.026] SE 0.019 *p* = .575 *q* = .818 |

**Notes:** Values are standardised regression coefficients (β), with 95% confidence intervals (CIs) in brackets, followed by standard errors (SE) and two-sided *p* values. N, number of offspring; Families, number of family clusters. Child, maternal and paternal PGS were entered simultaneously; child PGS coefficients index direct genetic effects and parental PGS coefficients index indirect genetic effects. SEs are family-clustered robust estimates; 95% CIs and *p* values use family-cluster degrees of freedom. False discovery rate (FDR)-adjusted *p* values (*q*; Benjamini–Hochberg) were computed separately for each estimate type across all PGS and outcomes in this table. Models adjust for sex, genotyping chip and the first ten genetic principal components.

**Supplementary Table S8. Trio-PGS analyses of ADHD and ASD latent factors** *(continued)*

#### Educational attainment PGS · Okbay et al. (2022)

| Outcome (trait, reporter) | N (offspring) | Families (clusters) | Child PGS (β, 95% CI, SE, *p*, *q*) | Maternal PGS (β, 95% CI, SE, *p*, *q*) | Paternal PGS (β, 95% CI, SE, *p*, *q*) |
| --- | --- | --- | --- | --- | --- |
| ADHD Common | 7,259 | 4,025 | −0.120 [−0.154, −0.085] SE 0.018 *p* < .001 *q* < .001 | −0.002 [−0.033, 0.029] SE 0.016 *p* = .897 *q* = .978 | −0.038 [−0.068, −0.007] SE 0.016 *p* = .016 *q* = .171 |
| ADHD Parent | 6,440 | 3,221 | −0.118 [−0.155, −0.081] SE 0.019 *p* < .001 *q* < .001 | 0.003 [−0.031, 0.037] SE 0.017 *p* = .876 *q* = .978 | −0.039 [−0.072, −0.005] SE 0.017 *p* = .023 *q* = .184 |
| ADHD Self | 4,631 | 2,563 | −0.052 [−0.098, −0.006] SE 0.023 *p* = .027 *q* = .066 | 0.033 [−0.005, 0.071] SE 0.019 *p* = .092 *q* = .271 | −0.011 [−0.049, 0.026] SE 0.019 *p* = .555 *q* = .818 |
| ADHD Teacher | 4,580 | 2,357 | −0.133 [−0.177, −0.090] SE 0.022 *p* < .001 *q* < .001 | 0.027 [−0.014, 0.068] SE 0.021 *p* = .200 *q* = .472 | −0.039 [−0.079, 0.001] SE 0.020 *p* = .056 *q* = .299 |
| ASD Common | 6,934 | 3,874 | −0.001 [−0.036, 0.035] SE 0.018 *p* = .972 *q* = .982 | −0.084 [−0.116, −0.053] SE 0.016 *p* < .001 *q* < .001 | −0.055 [−0.087, −0.024] SE 0.016 *p* < .001 *q* = .039 |
| ASD Parent | 5,930 | 2,965 | −0.001 [−0.039, 0.037] SE 0.019 *p* = .952 *q* = .982 | −0.097 [−0.133, −0.062] SE 0.018 *p* < .001 *q* < .001 | −0.054 [−0.089, −0.019] SE 0.018 *p* = .002 *q* = .064 |
| ASD Self | 5,224 | 2,719 | 0.001 [−0.040, 0.042] SE 0.021 *p* = .963 *q* = .982 | −0.029 [−0.069, 0.010] SE 0.020 *p* = .144 *q* = .369 | −0.009 [−0.050, 0.031] SE 0.021 *p* = .649 *q* = .885 |
| ASD Teacher | 4,575 | 2,409 | 0.003 [−0.042, 0.047] SE 0.023 *p* = .906 *q* = .982 | −0.021 [−0.060, 0.019] SE 0.020 *p* = .305 *q* = .592 | 0.016 [−0.024, 0.057] SE 0.021 *p* = .426 *q* = .715 |

**Notes:** Values are standardised regression coefficients (β), with 95% confidence intervals (CIs) in brackets, followed by standard errors (SE) and two-sided *p* values. N, number of offspring; Families, number of family clusters. Child, maternal and paternal PGS were entered simultaneously; child PGS coefficients index direct genetic effects and parental PGS coefficients index indirect genetic effects. SEs are family-clustered robust estimates; 95% CIs and *p* values use family-cluster degrees of freedom. False discovery rate (FDR)-adjusted *p* values (*q*; Benjamini–Hochberg) were computed separately for each estimate type across all PGS and outcomes in this table. Models adjust for sex, genotyping chip and the first ten genetic principal components.
